# Robust real-time estimation of pathogen transmission dynamics from wastewater

**DOI:** 10.1101/2025.10.23.25338640

**Authors:** Adrian Lison, Rachel McLeod, Jana S. Huisman, James D. Munday, Christoph Ort, Timothy R. Julian, Tanja Stadler

## Abstract

Wastewater monitoring has proven effective for tracking SARS-CoV-2 transmission during the COVID-19 pandemic. However, estimating transmission parameters for other pathogens remains difficult due to lower concentrations in sewage, uncertain shedding kinetics, and limited clinical validation data. Here we present a Bayesian semi-mechanistic wastewater model, called EpiSewer, which jointly accounts for uncertainty in infection dynamics, pathogen shedding, and measurement noise, including outliers and non-detects. This framework enables direct inference of transmission dynamics from raw concentration and flow data, eliminating the need for preprocessing such as smoothing, imputation, or outlier removal. It also provides short-term concentration forecasts for out-of-sample validation. We assessed EpiSewer across three seasons of multi-pathogen wastewater surveillance at 6–14 treatment plants in Switzerland, estimating the effective reproduction number (*R*_*t*_) for SARS-CoV-2, influenza A virus (IAV), and respiratory syncytial virus (RSV) in real time. *R*_*t*_ estimates were consistent and robust to measurement noise, even with IAV and RSV concentrations 10–50 times lower than SARS-CoV-2. Fourteen-day concentration forecasts were well-calibrated, with minimal bias during both epidemic growth and decline. Under reduced sampling frequencies, EpiSewer maintained unbiased forecasts while accurately reflecting increased uncertainty. Our approach enables robust inference of transmission dynamics for lower-abundance pathogens with limited clinical surveillance, using only a few wastewater samples per week.

## 1 Introduction

Wastewater-based infectious disease surveillance has gained significant traction during the COVID-19 pandemic as an effective tool for monitoring community-wide transmission dynamics [1]. When infected individuals shed genetic material of a pathogen into a municipal sewage system, the pathogen concentration in wastewater samples, determined using polymerase chain reaction (PCR) techniques, provides a signal of the trend in infections over time [2, 3]. The successful application of wastewater surveillance to SARS-CoV-2 has catalyzed the development of assays for other pathogens, including influenza A and B virus (IAV/IBV) [4, 5], respiratory syncytial virus (RSV) [6, 7], monkeypox virus [8], and human adenovirus [9]. Recent advances, such as digital PCR multiplexing, now allow the simultaneous detection of multiple pathogens in a single assay [10, 11], enabling cost-effective, multi-target surveillance through shared sampling and laboratory processes [12].

As wastewater surveillance programs expand to include a broader range of pathogens, wastewater-based estimates of transmission parameters can provide valuable insights into the spread of infectious diseases, supporting public health policy and health sector preparedness. For example, estimates of the effective reproduction number *R*_*t*_, indicating the average number of secondary infections caused per infected individual [13], can quantify trends and regional differences in the circulation of common viral infections and inform healthcare demand planning [14, 15]. Similarly, estimates of *R*_*t*_ and the epidemic growth rate can be used to identify exponential growth trends during outbreaks and support the implementation and evaluation of public health measures [16–18]. These applications of wastewater monitoring are especially promising for emerging and non-notifiable diseases, where clinical surveillance is often sparse or non-existent [19].

To date, wastewater-based *R*_*t*_ and growth rates have been primarily estimated for SARS-CoV-2 [20–27], which is found in wastewater at high concentrations and for which clinical surveillance data were widely available for model calibration and validation during the COVID-19 pandemic. Increasingly, other respiratory viruses such as IAV, IBV, and RSV, are being monitored in sewage across several countries [4–7, 10]. However, routine estimation of transmission dynamics from wastewater remains challenging due to several factors.

First, for many pathogens, measured gene concentrations are substantially lower than those of SARS-CoV-2, reducing the signal-to-noise ratio in wastewater data [7]. Specifically, PCR-based gene concentration measurements are subject to concentration-dependent measurement noise and non-detects [28, 29], which can frequently occur for pathogens detected at lower concentrations. These statistical properties are seldom reflected in epidemiological models, which typically assume – explicitly or implicitly – that wastewater measurements have a similar error structure as case counts [3, 21] or follow generic distributions such as the normal, log-normal or gamma distribution [25–27]. This also means that non-detects, i. e. zero measurements, must be imputed or removed from the data [26].

In addition, wastewater time series often contain gaps from unsampled days, typically due to cost constraints in monitoring programs [30, 31]. To interpolate and smooth these data, non-parametric methods, such as moving averages [20], LOESS [21], or smoothing splines [23], are commonly applied. These methods rely on the calibration of hyperparameters such as window sizes, weights, or knot positions, which can substantially affect the inferred transmission dynamics. Calibration is particularly challenging for pathogens with limited or no supporting clinical data, as the variability of wastewater measurements depends on numerous factors, including the pathogen’s shedding kinetics, sampling frequency, and laboratory methods [32]. Moreover, uncertainty from non-parametric smoothing is generally difficult to propagate to the subsequent modeling.

Finally, transmission parameter estimates are influenced by the specific shedding kinetics of each pathogen. Due to inherent delays between infection and shedding, wastewater concentrations lag behind transmission dynamics in the community [33, 34]. This lag in signal imposes a natural limit on the precision with which *R*_*t*_ or growth rates can be estimated close to the present, leading to overconfident real-time estimates if not accounted for [20, 21]. However, the shedding profiles of most pathogens remain poorly characterized [35, 36], and shedding profile uncertainty is typically neglected when estimating transmission parameters from wastewater [24, 27].

Collectively, these factors reduce the reliability of wastewater-based estimation of transmission dynamics. Moreover, because wastewater models are rarely evaluated in real time – partly due to the lack of ground truth for *R*_*t*_ or growth rates – errors in model specification can go unnoticed, for example when surveillance programs reduce sampling frequency to cut costs [30].

To address these challenges, we developed EpiSewer, a Bayesian semi-mechanistic wastewater model that explicitly represents the underlying infection, shedding, and measurement processes in wastewater surveillance. This enables routine transmission monitoring of multiple pathogens by estimating *R*_*t*_ and growth rates from raw, noisy concentration measurement data without prior smoothing, imputation, or outlier detection. Through epidemiologically interpretable parameters, the model can be adapted to different pathogens, taking into account limited knowledge of shedding characteristics through uncertain priors. Moreover, our generative modeling approach naturally provides probabilistic short-term forecasts of concentration measurements, enabling performance evaluation in near real time during ongoing surveillance.

We have used EpiSewer during three years of multi-pathogen wastewater monitoring in Switzerland to produce real-time estimates of *R*_*t*_, informing Swiss public health officials via a public dashboard at https://wise.ethz.ch with data from currently 10 wastewater treatment plants (WWTPs). Here we demonstrate the capabilities of our approach in quantifying the transmission dynamics of SARS-CoV-2, influenza A virus (IAV), and respiratory syncytial virus (RSV) across the last three seasonal waves. We evaluate the real-time performance of wastewater-based *R*_*t*_ estimates and pathogen concentration forecasts and test the model’s ability to handle non-daily sampling and outliers in the measurement data. Our findings highlight the strength of generative modeling to robustly infer transmission dynamics of pathogens from sparse and noisy wastewater data.

## 2 Results

### Wastewater model

EpiSewer is a Bayesian model describing the time evolution of pathogen concentrations in wastewater (Figure 1). In this model, new infections are generated through a time-discrete, stochastic renewal process with a smoothly time-varying effective reproduction number *R*_*t*_ (Figure 1A–B). After infection, individuals shed genetic material of the pathogen into the wastewater over a certain number of days according to a shedding profile (Figure 1C). The relative intensity of shedding on each day after infection follows a discretized, parametric distribution with uncertain mean and variance, and the total shedding load can vary across individuals. We further assume that genetic material is well mixed in sewage and diluted in proportion to the daily measured wastewater flow within the catchment (Figure 1D). Samples taken at a WWTP are assumed to be representative of the daily wastewater concentration, with the exception of occasional high outlier concentrations, e. g. due to incomplete mixing, which are modeled using an extreme value distribution (Figure 1E). Finally, pathogen concentrations in the sample are quantified using PCR, where measurements are subject to concentration-dependent noise and false-negative non-detects, i. e. a measured concentration of zero despite the presence of target nucleic acid in the sample (Figure 1F). We here model a digital PCR approach, in which sample extracts are independently amplified and tested across a large number of chambers or droplets. However, other techniques, such as quantitative PCR, can be incorporated in a similar manner.

**Fig 1.**
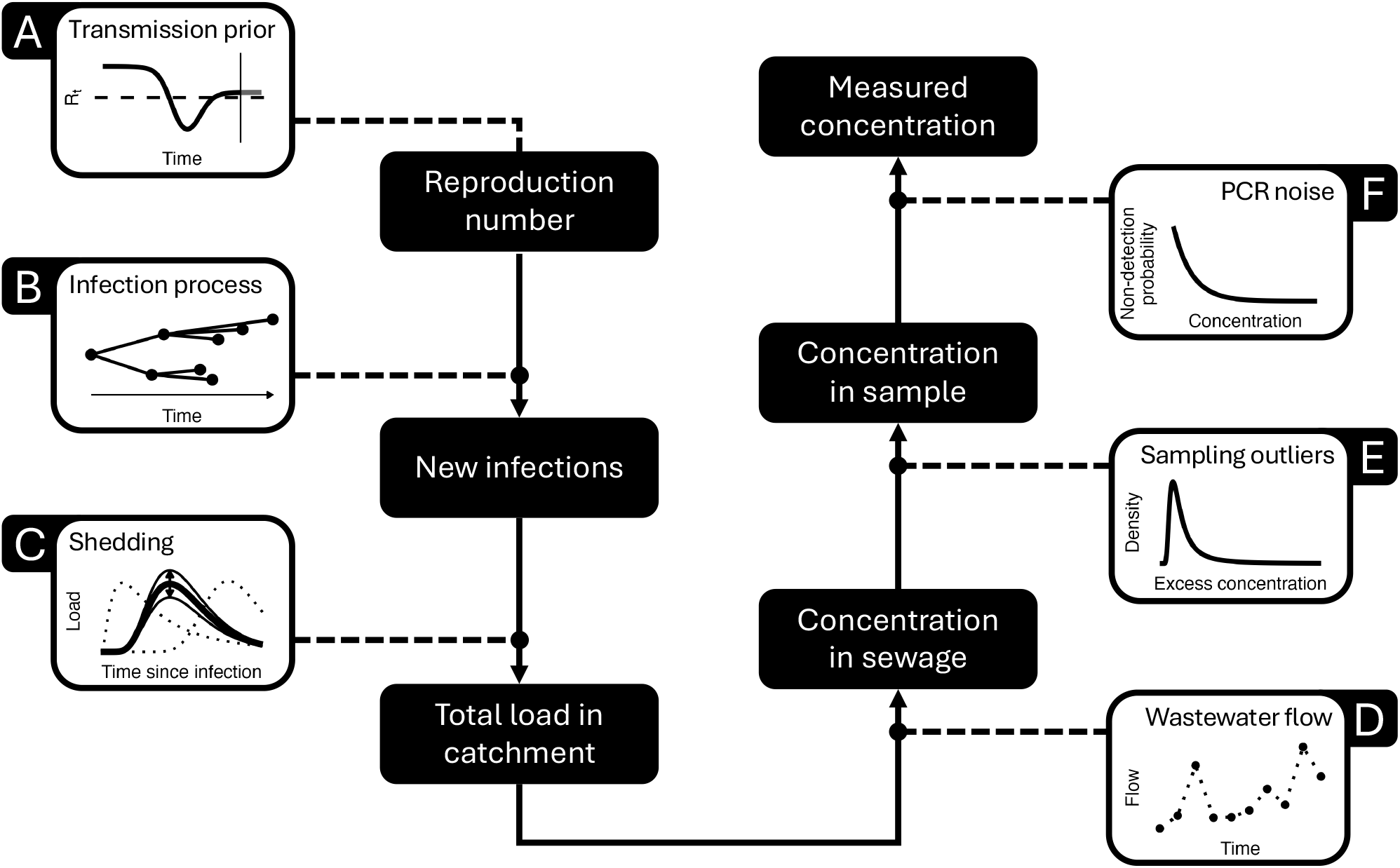
Visual summary of the EpiSewer model. A) Population-level transmission dynamics are modeled via smooth changes in the effective reproduction number *R*_*t*_. B) New infections are generated through an age-dependent branching process, represented by a stochastic renewal model. C) Infected individuals shed genetic material into the wastewater, modeled via a shedding load distribution with parameter uncertainty and individual-level variation in shedding intensity. D) The concentration of genetic material in the sewage depends on the dilution by wastewater flow within the catchment area. E) Concentrations in wastewater samples are assumed to be representative of sewage concentrations, with occasional outliers following an extreme value distribution. F) PCR-based concentration measurements are modeled with concentration-dependent measurement noise and non-detects. We here assume a digital PCR approach.

Using Markov Chain Monte Carlo sampling, EpiSewer is fitted directly to a time series of measured pathogen concentrations to produce Bayesian estimates of the effective reproduction number *R*_*t*_, epidemic growth rate *r*_*t*_, and other parameters of interest, e. g. infection numbers and loads. We also obtain a posterior predictive distribution of measurements on both sampled and unsampled days, allowing inspection of the model fit and out-of-sample validation.

### Handling of non-detects and outliers in concentration data

We fitted the model to concentration measurements of SARS-CoV-2, IAV, and RSV from a multi-year wastewater surveillance program in Switzerland that comprised 6 WWTPs during the winter season 2022/23, 14 WWTPs during the winter season 2023/24, and 10 WWTPs during the winter season 2024/25 [5, 37]. For each WWTP, pathogen concentrations were quantified using digital droplet PCR from an average of 5 samples per week until 2025, then from 4 samples per week. Figure 2A-C shows concentrations measured during the winter season 2023/24 in Switzerland, which featured clear seasonal waves of all three pathogens, for the catchments of Zurich (471000 persons), Lugano (124000 persons), and Chur (55000 persons). Results for further catchments and the winter seasons 2022/23 and 2024/25 are shown in Supporting Information F.

**Fig 2.**
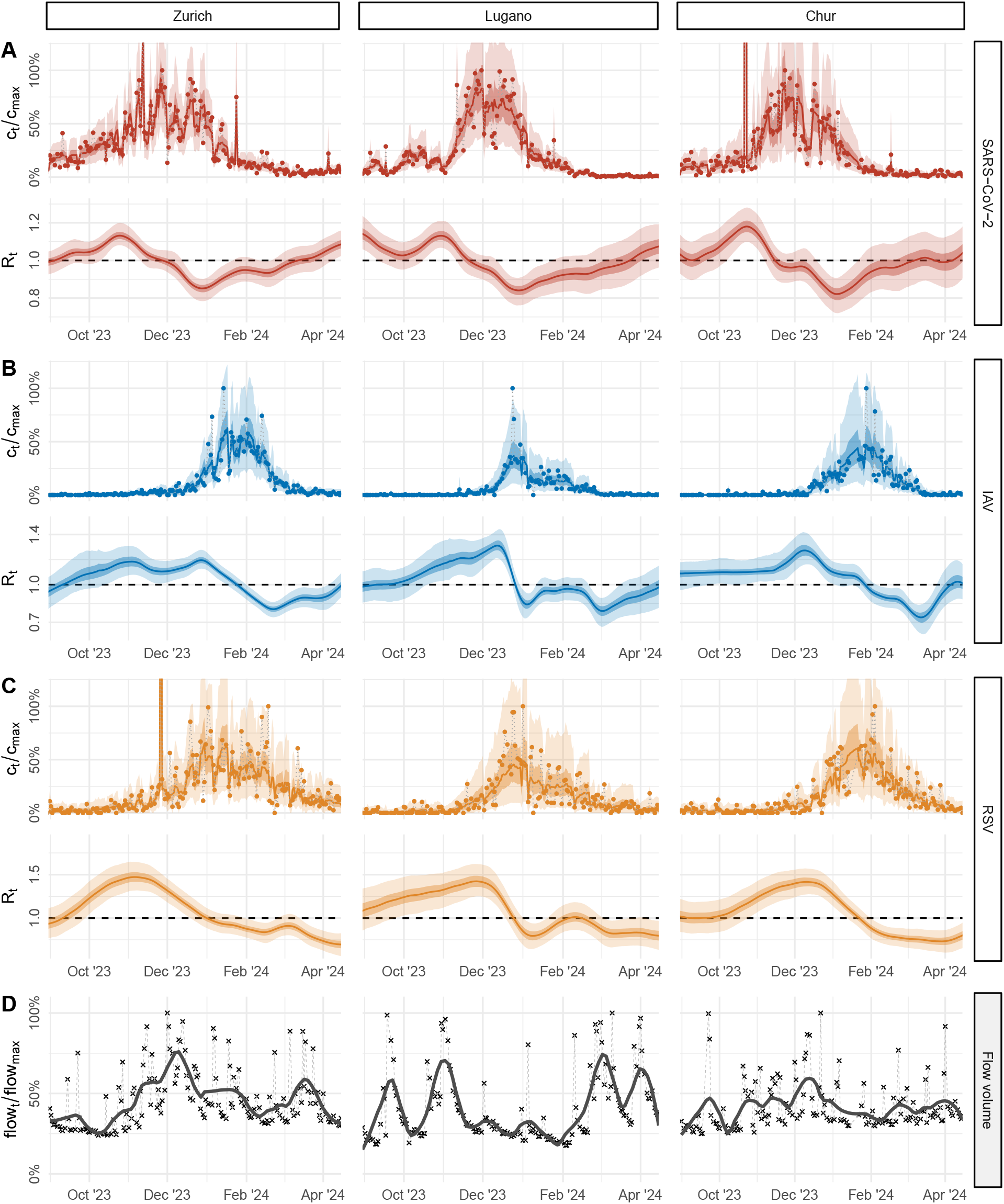
Viral transmission dynamics in Switzerland quantified from wastewater at treatment plants in Zurich, Chur, and Lugano during the 2023/24 winter season. (A-C) Relative pathogen concentrations (top) and estimates of the effective reproduction number *R*_*t*_ (bottom) for SARS-CoV-2 (red), influenza A virus (IAV; blue), and respiratory syncytial virus (RSV; orange) based on longitudinal samples from a large (Zurich, 471000 persons), medium-sized (Lugano, 124000 persons), and small (Chur, 55000 persons) wastewater catchment in Switzerland during the winter season 2023/24. Shown are measured pathogen concentrations (dots), the posterior median (lines), and 50% and 95% credible intervals (dark and light bands) of predicted concentrations and estimated *R*_*t*_, respectively. Concentrations were normalized by the maximum observed non-outlier concentration (*c*_max_). (D) Relative daily flow volumes at the sampled WWTP of each catchment. Lines show a smooth trend estimated using LOESS with a 4-week window. Flows were normalized by the maximum observed flow (flow_max_).

For each pathogen, gene concentrations were similar across catchments. However, absolute concentrations of IAV and RSV were considerably lower than those of SARS-CoV-2. Across the 14 WWTPs sampled, the maximum measured concentration (excluding outliers) ranged between 914–2973 gc*/*mL for SARS-CoV-2, but only 43–302 gc*/*mL for IAV and 18–104 gc*/*mL for RSV. The lower concentrations resulted in a large percentage of non-detects at the start of the winter season (46–83 % of IAV and 19–42 % of RSV measurements before December 31, 2023). Using our dPCR-specific measurement model, we were able to include these observations without imputation or censoring.

We also observed occasional extreme spikes in measured concentrations across all monitored seasons. By modeling excess sample concentrations via an extreme value distribution, EpiSewer can automatically identify individual outlier observations. For example, during the winter season 2023/24, we encountered outliers for SARS-CoV-2 in Chur (Oct 22, 2023) and for RSV in Zurich (Nov 26, 2023), falling into the 99.89% and 99.87% quantiles of the extreme value distribution (Figure 2). This allowed us to fit our model without prior outlier detection. Indeed, excluding detected outliers before fitting the model produced practically identical *R*_*t*_ estimates, indicating that they had no significant impact on the estimated transmission dynamics (Supporting Information E.2). Moreover, as shown for RSV in Zurich in Figure 3A, real-time *R*_*t*_ estimates remained robust even when the most recent observation was an outlier.

**Fig 3.**
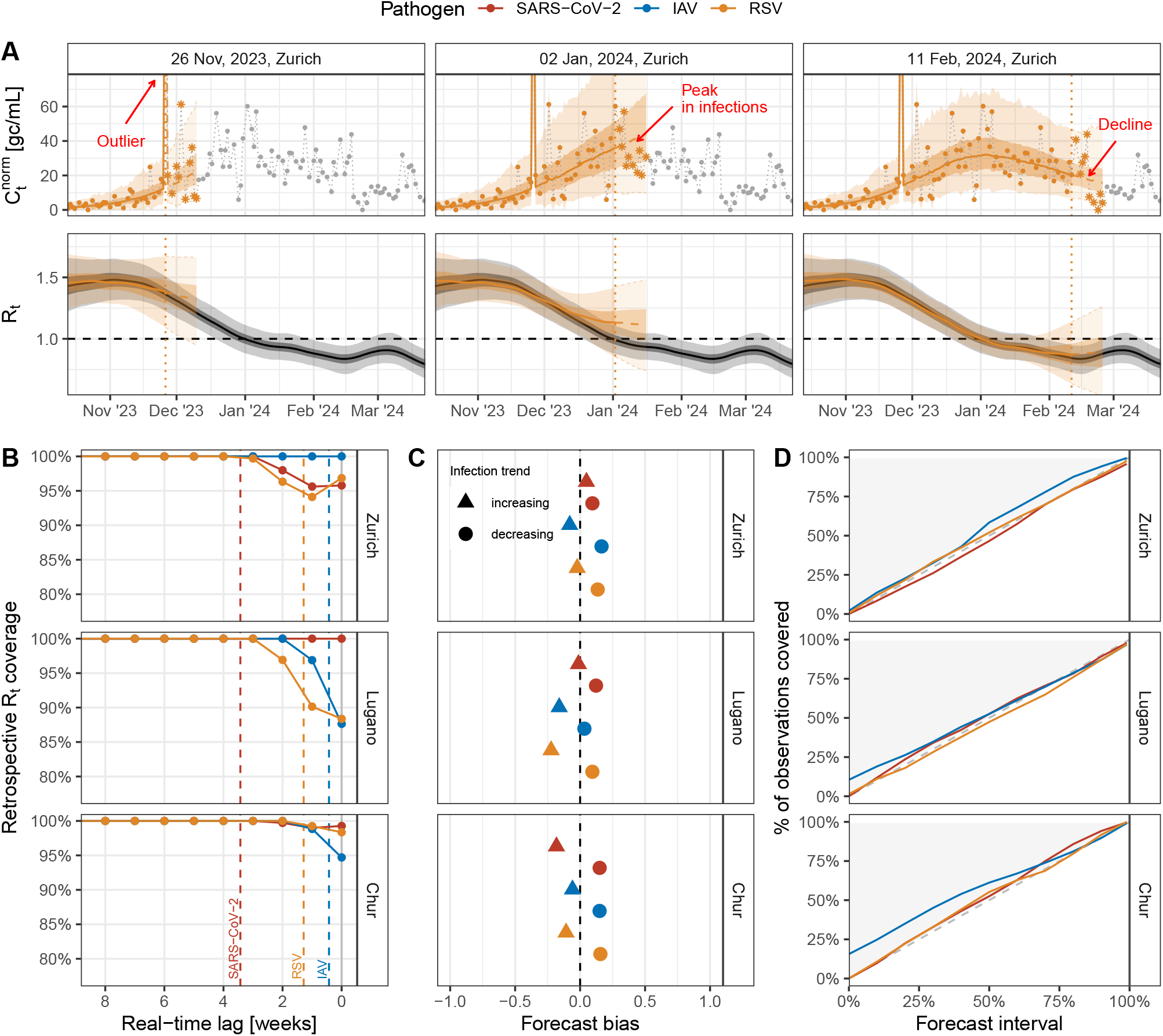
Real-time performance of wastewater-based *R*_*t*_ estimates and concentration forecasts. We evaluated real-time *R*_*t*_ estimates and 1–14-day-ahead forecasts of concentration measurements for the catchments of Zurich, Lugano, and Chur, Switzerland, during the 2023/24 seasonal wave of SARS-CoV-2, IAV, and RSV. (A) Example real-time estimates for RSV in Zurich on three estimation dates (vertical dotted lines): immediately after a high outlier measurement, at the peak of the seasonal wave, and during the declining phase of infections. Top panels show forecasts and observed concentrations (stars), normalized to median flow. Bottom panels show real-time estimates and 14-day-ahead projections of *R*_*t*_, along with retrospective *R*_*t*_ (in gray). Lines indicate posterior medians; shaded bands represent 50% (dark) and 95% (light) credible intervals. (B) Consistency of real-time *R*_*t*_ (estimated on each day with new data) with retrospective *R*_*t*_ (estimated at the end of the seasonal wave). For each real-time *R*_*t*_ estimate, we computed the retrospective coverage, i. e. the percentage of days on which the 95% credible interval (CrI) contained the retrospective median *R*_*t*_. Dots show the coverage stratified by lags of 0–8 weeks from the date of estimation. Vertical dashed lines show the 90% quantile of the shedding load distribution of each pathogen. (C) Bias of concentration measurement forecasts, stratified by increasing vs. decreasing infection trends. Scores range from −1 (maximum underprediction) to 1 (maximum overprediction); 0 indicates no bias. (D) Calibration of concentration measurement forecasts. The diagonal line indicates perfect calibration, where predictive intervals match their nominal coverage.

### Estimation of transmission dynamics

Based on measured wastewater concentrations, we estimated the effective reproduction number *R*_*t*_ over time for each pathogen and catchment (Figure 2A-C and Supporting Information F). Pronounced seasonal waves were observed for all pathogens in all catchments and seasons, except for SARS-CoV-2 during the 2024/25 season (Fig S21). Maximum *R*_*t*_ values were generally higher for RSV (median estimate 1.42 – 1.47 across catchments in 2023/24) than for SARS-CoV-2 (1.13 – 1.18) and IAV (1.19 – 1.31). This difference likely reflects the longer generation time assumed for RSV in our analysis, rather than faster epidemic growth. Indeed, the epidemic growth rate *r*_*t*_ estimated by EpiSewer was similar across pathogens (Figures S27–S29). While the timing of transmission dynamics varied by pathogen, it was largely synchronized across catchments. In 2023/24, we estimated that the peak in infections (indicated by the median *R*_*t*_ falling below 1) occurred earlier for SARS-CoV-2 (Nov 7 – Dec 2 across catchments), followed by RSV (Dec 13 – Jan 31), and IAV (Dec 14 – Feb 09). Due to pathogen-specific shedding delays, the estimated infection peaks generally preceded peaks in wastewater concentrations.

To model pathogen loads in wastewater based on observed concentration measurements, EpiSewer incorporates the daily wastewater flow at each WWTP, which varies over time due to rainfall and other environmental factors. For instance, during the peak of the 2023/24 SARS-CoV-2 wave, flow volumes were above average at the Zurich WWTP but below average at the Lugano WWTP (Figure 2D). When not using flow data and assuming a constant flow rate instead, the SARS-CoV-2 wave is underestimated in Zurich and overestimated in Lugano (Figures S9–S10). This highlights the importance of normalizing wastewater concentrations to correct for catchment-specific temporal differences in the dilution of genetic material in sewage.

### Real-time performance

We fitted EpiSewer on each day with new measurement data, obtaining real-time estimates of *R*_*t*_ and short-term forecasts of concentration measurements (see Movies S1–S3 for real-time animations of model predictions). Figure 3 summarizes the model’s real-time performance across the catchments of Zurich, Lugano, and Chur during the 2023/24 winter season. Real-time *R*_*t*_ estimates showed greater uncertainty near the present but were generally consistent with retrospective estimates computed at the end of each wave (Figure 3A). For estimates obtained in the same week as the latest observation, the 95% credible intervals of *R*_*t*_ captured the retrospective median *R*_*t*_ on 96–100 % of dates for SARS-CoV-2, 88–100 % for IAV, and 88–98 % for RSV (Figure 3B). With a one-week lag, retrospective coverage exceeded 90% for all pathogens and reached 100% after a three-week lag. The mean absolute error (MAE) of same-week median *R*_*t*_ estimates was 0.06–0.08 for SARS-CoV-2, 0.05–0.07 for IAV, and 0.10–0.14 for RSV, and fell below 0.05 for all pathogens after a three-week lag (Figures S24-S26).

By projecting the recent long-term trend in *R*_*t*_ forward, we generated probabilistic forecasts of concentration measurements 1–14 days ahead. At each estimation date, we computed the forecast bias, which compares predicted quantiles to observations and is optimal when the predicted median matches the observed concentration. We also assessed forecast calibration by comparing the percentage of observations that fall within a given forecast interval with the interval’s nominal coverage. Example forecasts for RSV in Zurich are shown in Figure 3A. Forecasts were generally well calibrated and showed little bias, indicating accurate prediction of short-term trends (Figure 3C–D). During periods of increasing infections, forecast bias was consistently above –22%, corresponding to an average underprediction by the 61% quantile (with 50% indicating an unbiased forecast).

During decreasing infections, forecast bias was consistently below 17%, corresponding to an average overprediction by the 41.5% quantile. Calibration was generally good across all pathogens, with forecast intervals capturing the expected proportion of observations. However, we observed a tendency towards over-coverage at narrow forecast intervals for IAV at smaller catchments. Comparable *R*_*t*_ consistency and forecast performance were achieved in the 2022/23 and 2024/25 winter seasons (Figures S22-S23). In the 2022/23 season, forecasts for IAV and RSV were overdispersed at the catchments of Lugano and Chur, as measurements were only available from November 1, 2022 onward.

### Robustness under sparse sampling

Due to logistical and cost constraints, many monitoring programs are not able to sustain daily sampling and analysis of wastewater, resulting in gaps in the measurement data. In the model, missing measurements are omitted from the likelihood function, eliminating the need for imputation while naturally accounting for the associated uncertainty. To evaluate the robustness of *R*_*t*_ estimates at lower sampling frequencies, we subsampled the wastewater data (5 sampled days per week on average) to 3 days per week, 1 day per week, and 1 day per 2 weeks. Figure 4A shows the resulting *R*_*t*_ estimates for the catchment of Zurich during the winter season 2023/24. As sampling frequency decreased, *R*_*t*_ estimates became more uncertain but preserved the overall trajectory (Figure 4A). Across pathogens, the mean absolute error (MAE) of retrospective median *R*_*t*_ estimated from subsampled data was 0.01–0.04 for 3 days per week, 0.01–0.07 for 1 day per week, and 0.01–0.10 for 1 day per 2 weeks (Figure S18). Estimates remained insensitive to the specific weekdays sampled at a frequency of 3 days per week, but became weekday-dependent at lower frequencies (Figure S19). This was particularly observed at the onset of the IAV and RSV seasons, where a high proportion of non-detects lead to a loss of signal for *R*_*t*_ under subsampling (Figure 4A). The bias of real-time concentration forecasts from subsampled data remained small, however, overdispersion increased when sampling only 1 day per week or less (Figure 4B–C).

**Fig 4.**
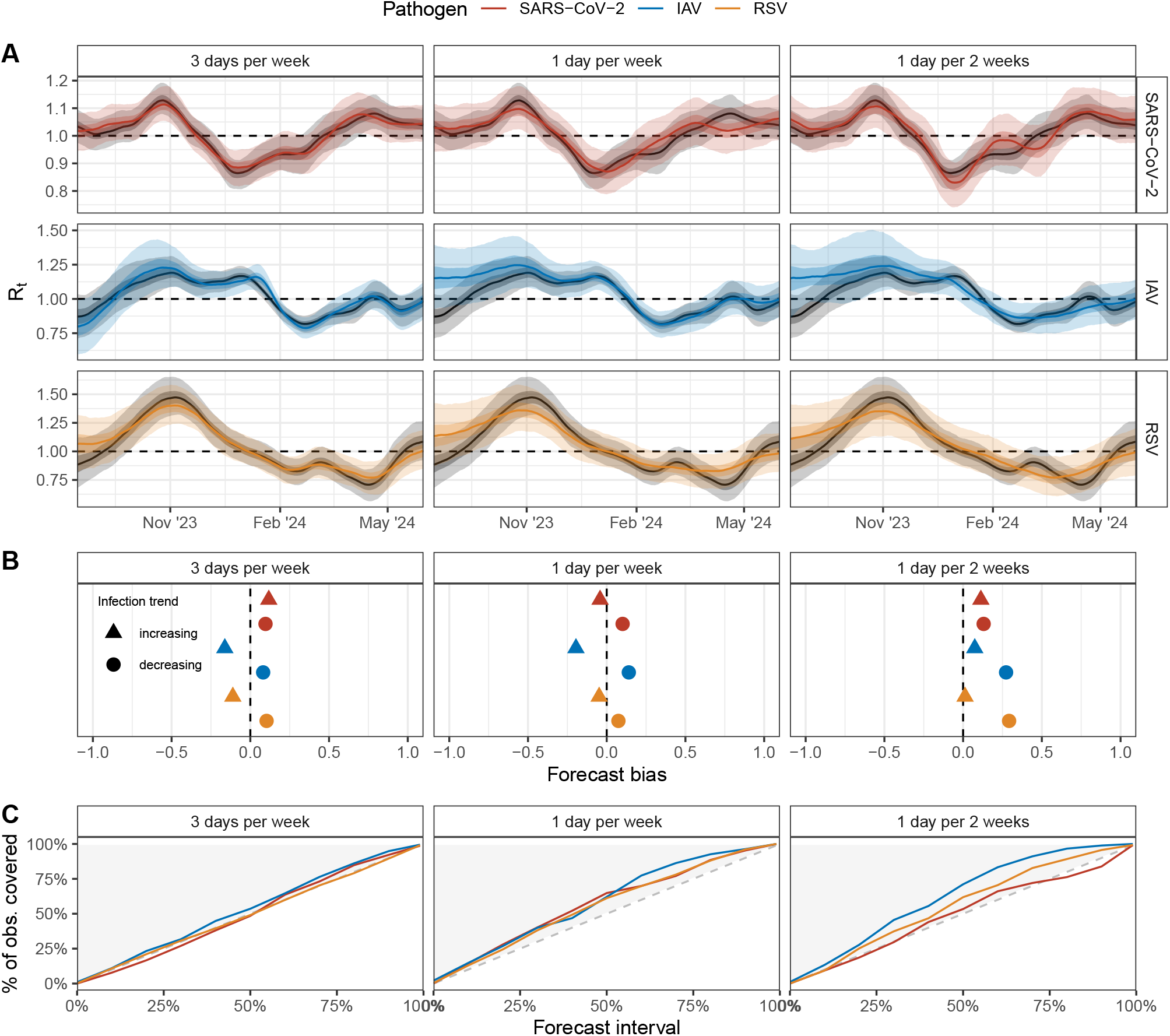
Effects of reduced sampling on *R*_*t*_ estimates and concentration forecasts. Concentration measurements of SARS-CoV-2, IAV, and RSV (catchment of Zurich, Switzerland, winter season 2023/24) were subsampled at different frequencies to simulate reduced sampling. (A) Retrospective *R*_*t*_ estimates on May 31, 2024 based on samples on 3 days per week (Mon/Tue, Wed/Thu, and Fri), 1 day per week (every Fri), and 1 day per 2 weeks (every other Fri). Lines show posterior medians; bands indicate 50% (dark) and 95% (light) credible intervals. The baseline using all available data (5 days/week) is shown in gray. (B) Bias of 1–14-day ahead concentration forecasts from models fitted on subsampled data, stratified by increasing vs. decreasing infection trends. Scores range from −1 (maximum underprediction) to 1 (maximum overprediction); 0 indicates no bias. (C) Calibration of 1–14-day ahead forecasts from models fitted on subsampled data. The diagonal line indicates perfect calibration, where predictive intervals match their nominal coverage.

## 3 Discussion

In this study, we developed EpiSewer, a Bayesian model for real-time quantification of pathogen transmission dynamics from wastewater. By explicitly modeling the infection, shedding, and measurement processes that underlie wastewater data, we could parameterize the model using interpretable assumptions about PCR-based measurement error and the pathogen’s generation time and shedding profile, which can be informed by independent epidemiological evidence. Unlike other methods that rely on preprocessing steps such as smoothing, imputation, or outlier detection to analyze wastewater data [26], EpiSewer avoids these steps entirely, reducing dependence on difficult-to-calibrate hyperparameters. This makes our approach well-suited for monitoring a broad range of pathogens through multi-target environmental surveillance [12], even if historical wastewater and clinical data are limited. As evidence on shedding kinetics remains sparse for many infectious diseases [31,35], we incorporate uncertainty in the shedding profile using a prior distribution over both the mean and coefficient of variation of the shedding distribution. We also modeled individual-level variation in shedding intensity, but found this to have minimal influence on inferred transmission dynamics, likely because WWTP-level concentrations alone do not provide sufficient information to distinguish individual variability from sampling and measurement noise.

For the pathogens analyzed in this study, real-time *R*_*t*_ estimates from EpiSewer were generally consistent with retrospective estimates, providing timely information about epidemic growth. This even applied to most same-week estimates, although these were subject to substantial uncertainty, stemming from delays in the transmission signal from wastewater. These inherent delays, resulting from biological lags between infection and pathogen shedding [33], occur independently of sample logistics or lab processing and are comparable to reporting delays in clinical data, which similarly obscure recent changes in transmission [38]. Our wastewater model captured the uncertainty arising from delayed shedding through wider posterior distributions for *R*_*t*_ near the present. However, point estimates, such as the posterior median, should be interpreted with caution, as they only reliably reflected transmission changes after a lag of 2–3 weeks. As such, a consolidation period may be necessary before using real-time estimates in public health decision-making. Importantly, because our *R*_*t*_ estimates are aligned to infection time rather than observation, the lags described here are not directly comparable to those between wastewater and clinical case data [2].

A major challenge in evaluating wastewater-based transmission models is the absence of ground truth for key epidemiological parameters such as *R*_*t*_. Previous studies have validated wastewater-based *R*_*t*_ estimates against case-based estimates derived from clinical data, but this approach is prone to biases in case ascertainment and reporting, and is sensitive to the method used to estimate *R*_*t*_ from cases [3, 20, 24, 39]. In this study, we used forecast-based metrics to evaluate the real-time performance of EpiSewer. This approach leverages the close link between real-time transmission dynamics and forecasting of observations in the generative model. Short-term concentration forecasts from EpiSewer showed little bias 1–14 days ahead and were generally well-calibrated, although we observed a slight tendency toward overdispersion in smaller catchments and for pathogens with low detectable concentrations, likely due to reduced signal-to-noise ratios when measurements are low and include many non-detects. These results indicate that the model reliably estimated the current growth trend in infections, which, beyond *R*_*t*_ estimation, is the basis for downstream tasks such as nowcasting and forecasting clinical cases or hospitalizations [27]. Forecast-based evaluation also enables continuous model assessment during real-time surveillance. For instance, by monitoring the accuracy of flow-normalized concentration forecasts over time, violated model assumptions about the shedding or measurement of a pathogen could be swiftly detected. Unlike assessments that rely on predicted clinical data [24, 26], this approach requires no auxiliary observations and is particularly suited for non-notifiable diseases that lack clinical reporting.

Due to resource and cost constraints, many wastewater monitoring programs have limited sampling frequency [30]. To evaluate how EpiSewer performs under such conditions, we tested its robustness by subsampling our wastewater data. Due to the model’s generative structure, increased measurement uncertainty from reduced sampling was inherently reflected in the *R*_*t*_ estimates, without requiring parameter adjustments. For the pathogens and sites analyzed, we found that duplicate measurements of 24-hour composite samples on three days per week were sufficient to produce accurate *R*_*t*_ estimates with little additional uncertainty. At lower sampling frequencies, e. g. once per week, EpiSewer still captured overall transmission dynamics but produced more uncertain estimates and, at very low concentrations, became sensitive to the specific weekdays sampled. These findings suggest that sampling frequency should ideally be tailored to the monitoring objective. For instance, identifying the onset of a seasonal wave likely requires three or more samples per week, whereas tracking overall *R*_*t*_ during peak transmission may tolerate slightly lower frequencies.

We note several limitations of our model. First, to account for the varying dilution of genetic material in wastewater, EpiSewer requires flow data from the monitored sampling site and so is limited to programs using influent wastewater. If flow data are unavailable, a constant flow can be assumed; however, this makes it impossible to account for seasonal fluctuations, e. g. due to rainfall. Alternative normalization approaches based on fecal shedding markers also exist [40,41]. These could be modeled similarly to flow data and and are also applicable to solids-based wastewater monitoring. Second, when modeling individual-level variation in shedding, we accounted only for differences in the absolute amount of genetic material shed, not for variation in the timing of shedding across individuals, as there is currently limited evidence to parameterize such models [33, 42]. However, for larger catchment populations, we expect the dominant source of uncertainty in wastewater loads to stem from uncertainty in the average shedding profile and from variation in the total amount shed, which we explicitly model in our approach. Third, we used a realistic error model tailored to concentration measurements from digital PCR, which is widely used in wastewater surveillance due to its reliable quantification of absolute target concentrations [43]. Other studies, however, often rely on quantitative PCR, which can be modeled in EpiSewer using simpler likelihood functions such as the log-normal distribution. Quantitative PCR has similar error characteristics as digital PCR, including the occurrence of non-detects [29], which EpiSewer also supports through a hurdle model likelihood. Finally, we modeled the infection process using a stochastic renewal model that does not account for population structure or differences in population size. However, the population contributing to transmission may differ from the wastewater catchment population, and the latter may fluctuate due to commuting and other mobility patterns [31, 44]. Incorporating such differences, e. g. using mobility data and information on contact patterns, may be important when modeling low-prevalence pathogens or catchments with substantial population variability.

We implemented the EpiSewer model as a user-friendly and configurable open-source R package to support its use across different surveillance settings [45]. As part of our ongoing multi-pathogen wastewater surveillance in Switzerland, we will continue to publish up-to-date reproduction number estimates on our public dashboard (https://wise.ethz.ch) for Swiss health authorities and the general public. As wastewater monitoring expands to new pathogens of interest, semi-mechanistic modeling of wastewater data, as demonstrated in this work, allows robust estimation of transmission dynamics in real time and provides the necessary interpretability to check for violated epidemiological assumptions and other forms of model misspecification. This will enable more detailed analysis of wastewater data to complement traditional public health surveillance and provide insights into the transmission of clinically under-monitored and emerging infectious diseases.

## 4 Methods

### Wastewater data

In November 2022, Eawag, Swiss Federal Institute of Aquatic Science and Technology, launched the combined monitoring of SARS-CoV-2, influenza A and B virus (IAV and IBV) and respiratory syncytial virus (RSV) in Swiss wastewater [5, 37]. Autosampler-based, 24-hour volume-proportional composite samples of raw influent wastewater were collected on average 5 days per week from 6 municipal WWTPs across Switzerland and analyzed using a droplet-based, four-plex digital PCR (dPCR) assay for SARS-CoV-2 (N2 gene), influenza A virus (M gene), influenza B virus (M gene), and RSV (N gene), as previously described in Nadeau et al. [5]. Target concentrations in gene copies per mL were measured using two technical replicates per sample and summarized by the arithmetic mean. In July 2023, an improved six-plex dPCR assay with two additional targets (N1 gene of SARS-CoV-2, M gene of Murine Hepatitis Virus for extraction efficiency control) was introduced, and monitoring was expanded to a total of fourteen WWTPs [37]. The catchment areas of the treatment plants vary in size from 16500 to 471000 inhabitants and together cover 27% of the Swiss population. Since January 2025, the monitoring was downsized to ten WWTPs. Further details on the treatment plants, analyzed samples, extraction, and preprocessing are provided in Supporting Information A. In our analysis, we used the N2 gene as a target for SARS-CoV-2, which was measured by both the four-plex and the six-plex assay. Furthermore, since IBV did not consistently show epidemic waves during the monitored seasons, we only included results for IAV. To account for the varying dilution of wastewater due to rainfall and other factors, we used the daily flow measured at each treatment plant in our analysis.

### Model definition

In the following, we define the EpiSewer model, a Bayesian generative model of wastewater data that can be used to estimate the effective reproduction number *R*_*t*_ and other measures of transmission. Our model applies to a time series of concentration measurements for a specific sampling site and target gene. We use a unified time index *t* ∈ *{*1, 2,…, *T}* for all time-dependent variables, where *t* = 1 denotes the first day of the modeled infection trajectory and *t* = *T* the day of the last wastewater measurement. Due to delays in shedding by infected individuals, we also model infections before the first wastewater measurement, i. e. wastewater measurements start only on day *t* = 1 + *S*, with shedding of genetic material up to *S* days after infection.

To model daily measured pathogen concentrations *c*_*t*_, we use a dPCR-specific likelihood from Lison et al. [46] that accounts for concentration-dependent measurement noise and false negatives. Specifically, we let *λ*_*t*_ be the expected target concentration in the wastewater sample collected on day *t*. Using *p*_zero_(*λ*_*t*_), the probability of false non-detection from dPCR, we define a hurdle model likelihood for dPCR measurements (Supporting Information B), i. e.

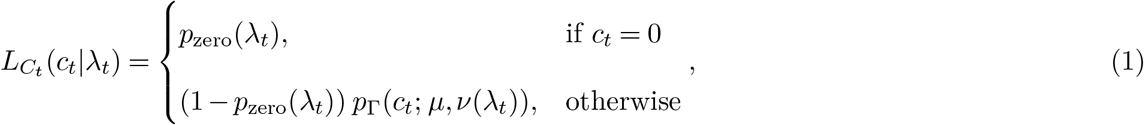

where *p*_*Γ*_ is the probability density of non-zero concentrations, modeled as Gamma distributed with mean 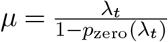 and coefficient of variation *ν*(*λ*_*t*_). This likelihood accounts for the partitioning statistics of dPCR [28] and allows direct fitting to concentration measurements, including non-detects. We note that *p*_zero_(*λ*_*t*_) and *ν*(*λ*_*t*_) depend on lab-specific parameters such as the pre-PCR coefficient of variation and the volume and number of partitions in the dPCR assay, which we jointly estimate from the measurement data (Supporting Information C.1).

Assuming a sewer residence time of less than one day and uniform mixing of genetic material in the wastewater [3], we model the expected concentration *λ*_*t*_ in a 24-hour composite sample as

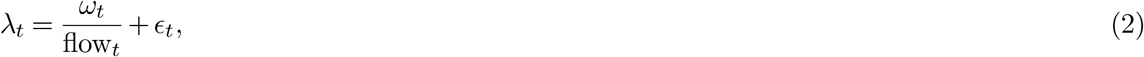

where *ω*_*t*_ is the daily expected detectable load shed in the catchment, flow_*t*_ is the measured daily wastewater flow volume at the sampling site, and *ϵ*_*t*_ *>* 0 represents positive outlier concentrations that violate the uniform mixing assumption. We use *ϵ*_*t*_ to model occasional spikes in concentrations as regularly found in wastewater data [47]. As a prior for *ϵ*_*t*_, we use a Type II extreme value distribution with parameters chosen such that 99% of the probability mass is below a concentration that would be observable from one infection in the catchment, which matches historically observed outlier frequencies in our data (Supporting Information C.4). This heavily right-tailed prior can capture extreme spikes in concentrations while having a negligible influence on modeled concentrations on most days.

The expected load shed in the catchment on day *t*, denoted *ω*_*t*_, depends on the numbers of individuals infected on previous days and their intensity of shedding *s* days after infection. We model *ω*_*t*_ via a convolution

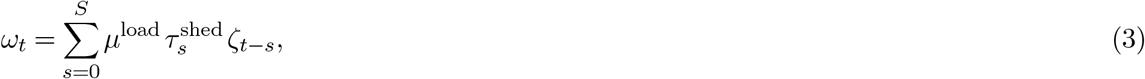

where *µ*^load^ is the average detectable total shedding load per person, 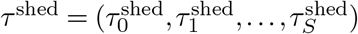 is a discrete distribution of the shedding load over time, and 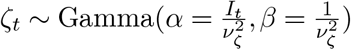 is the number of infected individuals *I*_*t*_, weighted by their overall shedding intensity. Here, the coefficient of variation *ν*_*ζ*_ describes how the strength of shedding varies across individuals, with an intensity *<* 1 indicating below-average shedding and an intensity *>* 1 indicating above-average shedding. We obtain the shedding load distribution by placing priors on the mean *µ*^shed^ and coefficient of variation *ν*^shed^ of a positive continuous distribution (e. g. gamma or log-normal), and transforming it into a discrete distribution *τ* ^shed^ within the model (Supporting Information C.2). This approach has been previously used to model reporting delay distributions [48, 49] and here allows us to account for uncertain shedding profiles.

To model the number of individuals infected on day *t*, we use a stochastic renewal model similar to earlier work [38, 50]. This defines the expected number of infections *ι*_*t*_ through the renewal equation

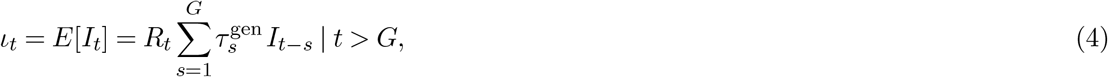

where *R*_*t*_ is the instantaneous effective reproduction number on day *t* [13] and 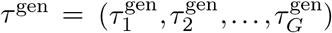 is a discrete generation time distribution with maximum generation time *G*. The realized number of infections *I*_*t*_ is modeled as Poisson distributed, i. e.

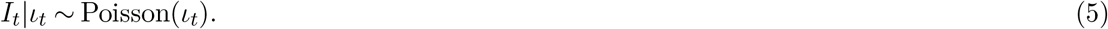

At the start of the modeled time period, when the renewal model cannot be applied, we model a seeding phase [51] in which the expected number of infections follows an exponential growth process with initial value *ι*_1_ and a time-varying growth rate *r*_*t*_, i. e.

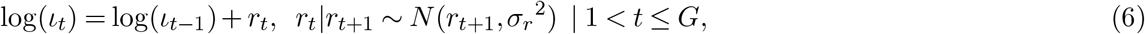

where *r*_*t*_ follows a backward-in-time random walk with initial value *r*_*G*+1_ and standard deviation *σ*_*r*_. We set *r*_*G*+1_ to be consistent with the reproduction number *R*_*t*_ at the end of the seeding phase by solving the renewal equation 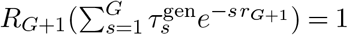 [52]. If there are many non-detects at the beginning of the measured time series, we extend the seeding phase until the first date with three consecutive detects. From the estimated number of infections, we also compute *r*_*t*_ for *t > G* using an infectiousness-based estimator by Parag et al. [53] (Supporting Information C.5).

### Priors and epidemiological assumptions

We informed model parameters related to infection and shedding based on findings from the epidemiological literature. For the generation time distribution, we obtained parametric distributions for the serial interval for SARS-CoV-2 [54], human influenza [55], and RSV [56, 57] and discretized them with a daily resolution. For the shedding load distribution, we used uncertainty information from estimates in the literature to define appropriate priors for the mean and coefficient of variation of the distribution. For SARS-CoV-2, we used a Gamma distribution with a mean of 8.0–17.7 days based on an observational study combining wastewater and case surveillance data [34]. For IAV and RSV, we used viral load measurements from nasal wash or swab samples as a proxy for shedding into wastewater. Specifically, we used a Gamma distribution with a mean of 2.1–2.9 days for IAV, based on a meta-analysis of volunteer challenge studies [5, 58]. For RSV, we used a Gamma distribution with a mean of 4.4–9.2 days based on a household study by Otomaru et al. [59]. Details on the discretization of the shedding load distribution in our model and the priors used for each pathogen are provided in Supporting Information C.2.

We assumed a pathogen-and catchment-specific total detectable load per infection, i. e. the absolute number of gene copies shed by an infected individual and detectable from wastewater samples [3], which also depends on the degradation of genetic material during in-sewer transport and storage [60] and extraction efficiency in the lab. In our main analysis, we calibrated this parameter to catchment-specific infection numbers estimated using publicly available data from Sentinella reporting and participatory surveillance in Switzerland (Supporting Information C.3). However, we obtained practically identical estimates for most catchments using a simpler calibration, in which the initial load detected at the start of each season was equated to an incidence of 0.01% of the catchment population (Figures S13–S15). This is because the load per infection serves as a constant scaling factor in the model, influencing estimated trends only when incidence is low enough for stochastic infection and shedding dynamics to dominate (Figure S16).

To account for heterogeneity in shedding [33, 35, 42, 61], we modeled individual-level variability in shedding loads. Due to the lack of quantitative estimates in the literature, we assumed a coefficient of variation of 1, implying that the top 25% of individuals contribute 60% of the total load, and the bottom 25% less than 4%, consistent with observed SARS-CoV-2 shedding variability [33]. Sensitivity analysis showed that transmission estimates are largely insensitive to this assumption (Supplementary Figure S17).

To regularize the estimated *R*_*t*_ trajectory, we decomposed the reproduction number into a long-term trend (*α*_*t*_) and a short-term deviation (*δ*_*t*_), each modeled with an approximate Gaussian process [49, 62] using long- and short-term kernels, respectively, i. e.

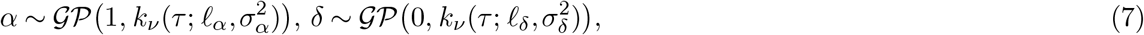

with Matérn kernel *k*_*ν*_ and smoothness parameter 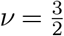. For the long-term trend, we used truncated normal priors on the length scale *𝓁*_*α*_ (mean 12 weeks, SD 1 week) and kernel magnitude *σ*_*α*_ (mean 0.4, SD 0.1). For the short-term deviation, we placed truncated normal priors on *𝓁*_*δ*_ (mean 3 weeks, SD 0.5 weeks) and *σ*_*δ*_ (mean 0.2, SD 0.05). To ensure *R*_*t*_ *>* 0, we computed the reproduction number as *g*(*R*_*t*_) = *α*_*t*_ + *δ*_*t*_, where 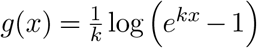 is an inverse softplus link with sharpness parameter *k* = 4, yielding an approximately linear mapping for realistic *R*_*t*_ values. The smoothing of *R*_*t*_ is described in more detail in Supporting Information C.5. For the laboratory parameters of our dPCR-specific measurement model, we used broad priors reflecting typical dPCR instruments and protocols (Supporting Information C.1).

### Estimation

Our model defines a likelihood for observed concentration measurements *c* given *R*_*t*_, measured flow, and all other epidemiological parameters. We use Markov Chain Monte Carlo to draw samples from the posterior distribution of *R*_*t*_, i. e.

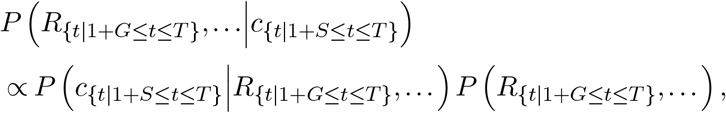

where *P R*_*{t*|1+*G*≤*t*≤*T }*_,… is our prior for *R*_*t*_ and the other parameters. For this, we implemented our model in the probabilistic programming language “stan” [63]. We fitted the model by running 4 chains, each with 1000 warm-up and 2500 sampling iterations, of the No-U-Turn Sampler (NUTS) in cmdstan version 2.34.1 through the cmdstanr interface [64]. We assessed convergence and effective sample sizes using standard Bayesian diagnostics [65,66]. Further details on the model implementation, sampling, and diagnostic checks can be found in Supporting Information D. The model is integrated into the open-source R package “EpiSewer” [45], which facilitates model specification, fitting, and visualization of results.

### Forecasting

To produce short-term forecasts of concentration measurements, we projected transmission dynamics forward by evaluating the Gaussian processes for the long-term trend and short-term deviations in *R*_*t*_ up to 14 days into the future. Due to the different length scales of the long-term and short-term component, forecasts naturally tend towards the recent long-term trend. For each posterior sample from the fitted model, we then propagated the renewal equation forward using the forecasted *R*_*t*_ trajectory and other sampled parameters, applying the shedding and measurement components to simulate dPCR measurements on the forecasted dates. Unknown future flow volumes were imputed with the median of historical values. By combining simulated measurements from all posterior samples, we obtained a posterior predictive distribution for future concentration measurements.

### Real-time performance and subsampling study

We assessed the performance of real-time estimation during seasonal infection waves of SARS-CoV-2, IAV, and RSV. For each pathogen, we defined the start of the seasonal wave as the first week in the second half of the year with a 10% weekly growth rate and no consecutive non-detects before the peak. We defined the end of a seasonal wave as the first week three months after the peak with a decrease in weekly incidence of less than 10%. On each day of the wave with new measurements, we obtained real-time *R*_*t*_ estimates for the respective pathogen by fitting EpiSewer to all measurements up to that date, using data from 1 month before the start of the wave. For each date, we also produced real-time forecasts of the measured concentrations for the next 14 days. We note that in the winter season 2022/23, measurements for Lugano, Chur, Sensetal, and Altenrhein were only available from November 1, 2022 onward. Posterior *R*_*t*_ and forecast distributions were summarized using the median and the 1%, 3%, …, 97%, and 99% credible intervals. Real-time estimates were compared to a retrospective estimate based on all measurements up to August 1 of the following year (June 1 for 2025). For this, we calculated the percentage of days on which the retrospective *R*_*t*_ estimate was covered by the 95% credible interval of the real-time estimate, stratified by lag. To evaluate the performance of short-term concentration forecasts, we computed scores for forecast calibration [67], i. e. the percentage of observations within nominal forecast intervals, and forecast bias [68], i. e.

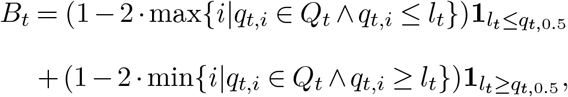

where *l*_*t*_ = *c*_*t*_ *×* flow_*t*_ is the flow-normalized concentration measurement and *Q*_*t*_ is the set of lower and upper quantiles of the credible intervals of the forecast distribution for day *t*. Calibration curves and bias were averaged across forecasts and estimation dates. To mimic sparse sampling, we subsampled SARS-CoV-2, IAV, and RSV measurement time series from the Zurich catchment during the winter season 2023/24 to lower frequencies, i. e. 3 days per week (Mon/Tue, Wed/Thu, and Fri), 1 day per week (every Fri), and 1 day per 2 weeks (every other Fri). The original time series had a frequency of 5 days per week, with measurements missing only on Mon and Wed in one week and on Tue and Thu the following week. When subsampling, we replaced days with missing measurements in the original series by the next day when possible. To evaluate the real-time performance of EpiSewer on subsampled data, we measured the calibration and bias of short-term concentration forecasts as described above. To examine the impact of the specific weekdays sampled, we computed retrospective *R*_*t*_ estimates for all possible weekday combinations of each subsampling frequency.

## Acknowledgments

This work features a secondary analysis of open wastewater data provided by Eawag, Swiss Federal Institute of Aquatic Science and Technology. We kindly thank the members of the Wastewater Monitoring Laboratory, including Lea Caduff, Sheena Conforti, Seju Kang, Jolinda de Korne-Elenbaas, Charles Gan, Melissa Pitton, Linda Schneider, Nadja Widrig, Nadine Hürlimann, and Anna Wettlauffer, for their contributions to the development of digital PCR assays and their application in ongoing wastewater surveillance. We thank the Swiss Federal Office of Public Health FOPH for supporting the wastewater surveillance by Eawag. We thank Sam Abbott, Kaitlyn Johnson, Dylan Morris, and Damon Bayer for insightful discussions about the modeling of shedding loads and more. We thank Samuel Brand for his input on seeding of renewal models.

## Declarations

### Author Contributions

**Conceptualization:** A.L., J.S.H.

**Data curation:** A.L., R.E.M., and J.D.M.

**Formal Analysis:** A.L.

**Funding acquisition:** C.O., T.R.J., and T.S.

**Investigation:** A.L.

**Methodology:** A.L.

**Project administration:** A.L.

**Resources:** T.S.

**Software:** A.L.

**Supervision:** T.J.

**Validation:** A.L.

**Visualization:** A.L.

**Writing – original draft:** A.L.

**Writing – review & editing:** A.L., R.E.M., J.S.H., J.D.M., C.O., T.R.J., and T.S.

### Ethics approval

Ethics approval was not required for this study.

### Data availability

All data and analysis scripts are publicly available at https://github.com/adrian-lison/EpiSewer-study. The EpiSewer model is available as an open-source R package at https://github.com/adrian-lison/EpiSewer.

