## Supporting information for "Robust real-time estimation of pathogen transmission dynamics from wastewater"

### Supporting information for: Robust real-time estimation of transmission dynamics from wastewater

**F Additional results** **29**

**References** **47**

### A Wastewater data

We used data from three seasons of wastewater surveillance conducted by the Swiss Federal Institute of Aquatic Science and Technology (EAWAG). During the respiratory virus season 2022/23, samples were collected since November 2022 from 6 wastewater treatment plants and analyzed with a frequency of 5 days per week, i. e. all weekdays except for Wednesdays and Fridays (Figure S1). During the season 2023/24, samples were collected from 14 wastewater treatment plants and analyzed with a frequency of 5 days per week, using a biweekly sampling scheme that varied between batch groups (Figure S2). During the season 2024/2025, samples were collected from 10 wastewater treatment plants and analyzed with a frequency of 4 days per week, again using a biweekly sampling scheme that varied between batch groups (Figure S3). The population of the associated catchment of each wastewater treatment plant is given in Figures S1 to S3, respectively.

The specific extraction and quantification protocol used has been described earlier in Huisman et al. [1] and Nadeau et al. [2]. Briefly, vacuum filtration was used to extract viral RNA from a 40 mL aliquot of each sample. The resulting elution of 80 µL was diluted by a factor of 1:3. A 25 µL reaction mix with 5 µL of the template and 20 µL of reagents was created. Target concentrations were then quantified by running a droplet-based, multiplex digital PCR assay using the Naica Crystal Digital PCR System (Stilla® Technologies). Sapphire Chips with a maximum number of 30000 droplets and an average droplet volume of 0.519 nL were used. Each sample was run in duplicate, and we used the arithmetic mean of the replicate measurements for analysis.

During the respiratory virus season 2022/23, a fourplex digital PCR assay with the targets SARS-CoV-2 (N2 gene), Influenza A (M gene), Influenza B (M gene), and RSV (N gene) was used. For the seasons 2023/24 and 2024/2025, a similar, six-plex assay with two additional targets (N1 gene of SARS-CoV-2, M gene of Murine Hepatitis Virus for extraction efficiency control) was used. Here, we focused our analysis on the targets SARS-CoV-2 (N2 gene), Influenza A (M gene), and RSV (N gene), which were consistently found across seasons.

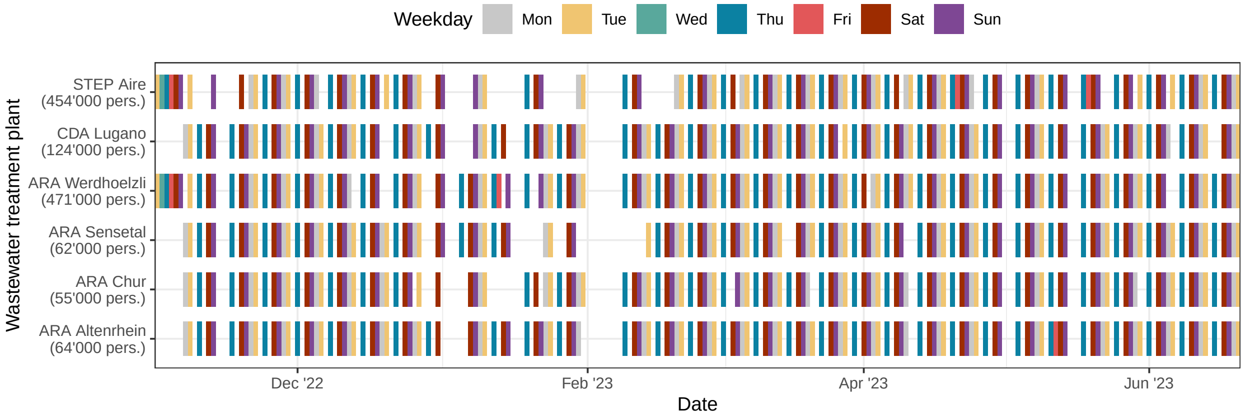

**Fig S1. Overview of analyzed samples from 6 wastewater treatment plants during the 2022/23 respiratory virus season in Switzerland.** As a rule, 5 samples per week (all weekdays except for Wednesdays and Fridays) from each treatment plant were analyzed. For each sample, the concentration of SARS-CoV-2 (N2 gene), Influenza A and B virus (M gene), and Respiratory syncytial virus (N gene) was quantified in duplicate using a fourplex digital PCR assay. Only samples which passed quality control are shown.

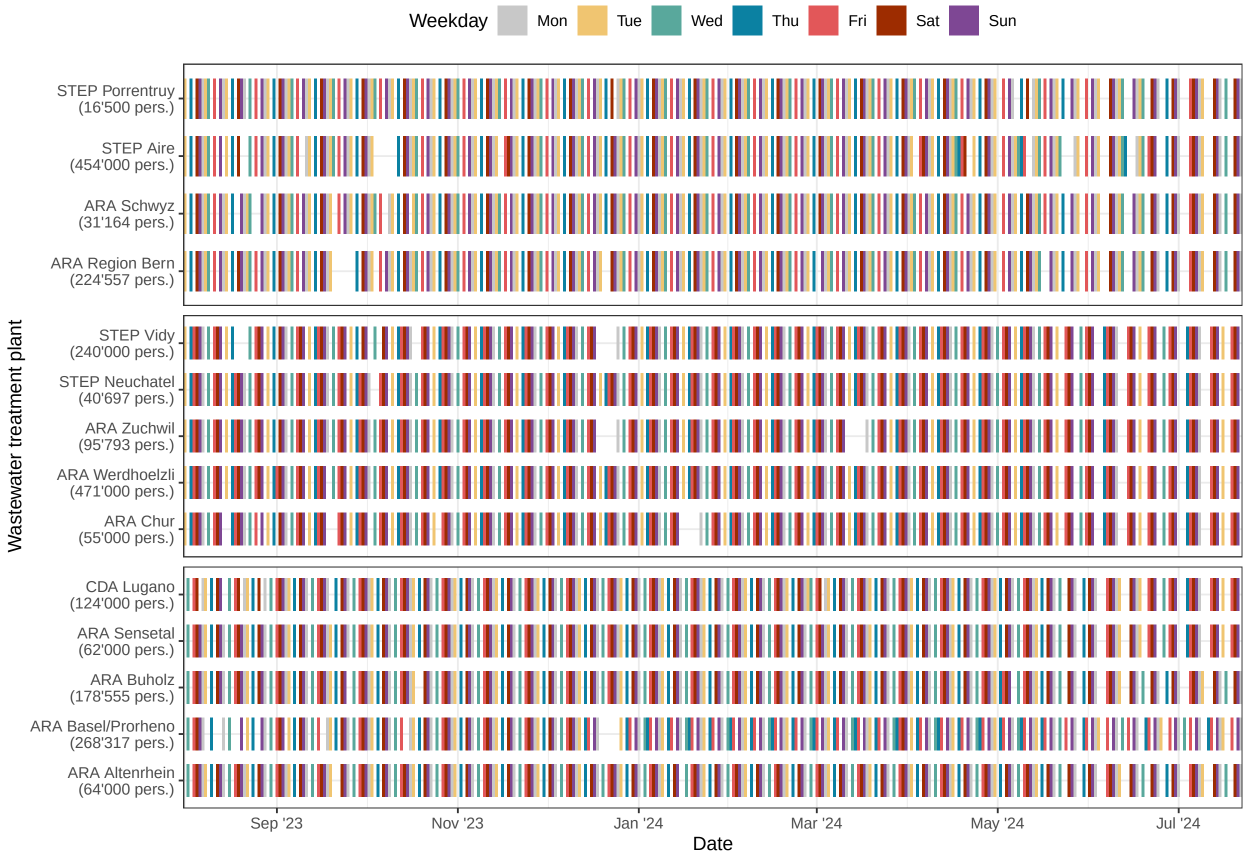

**Fig S2. Overview of analyzed samples from 14 wastewater treatment plants during the 2023/24 respiratory virus season in Switzerland.** As a rule, 5 samples per week from each treatment plant were analyzed. The specific weekdays analyzed varied between batch groups and alternated on a biweekly basis. For each sample, the concentration of SARS-CoV-2 (N1 and N2 genes), Influenza A and B virus (M gene), Respiratory syncytial virus (N gene), and Murine Hepatitis Virus (M gene, internal control) was quantified in duplicate using a six-plex digital PCR assay. Only samples which passed quality control are shown.

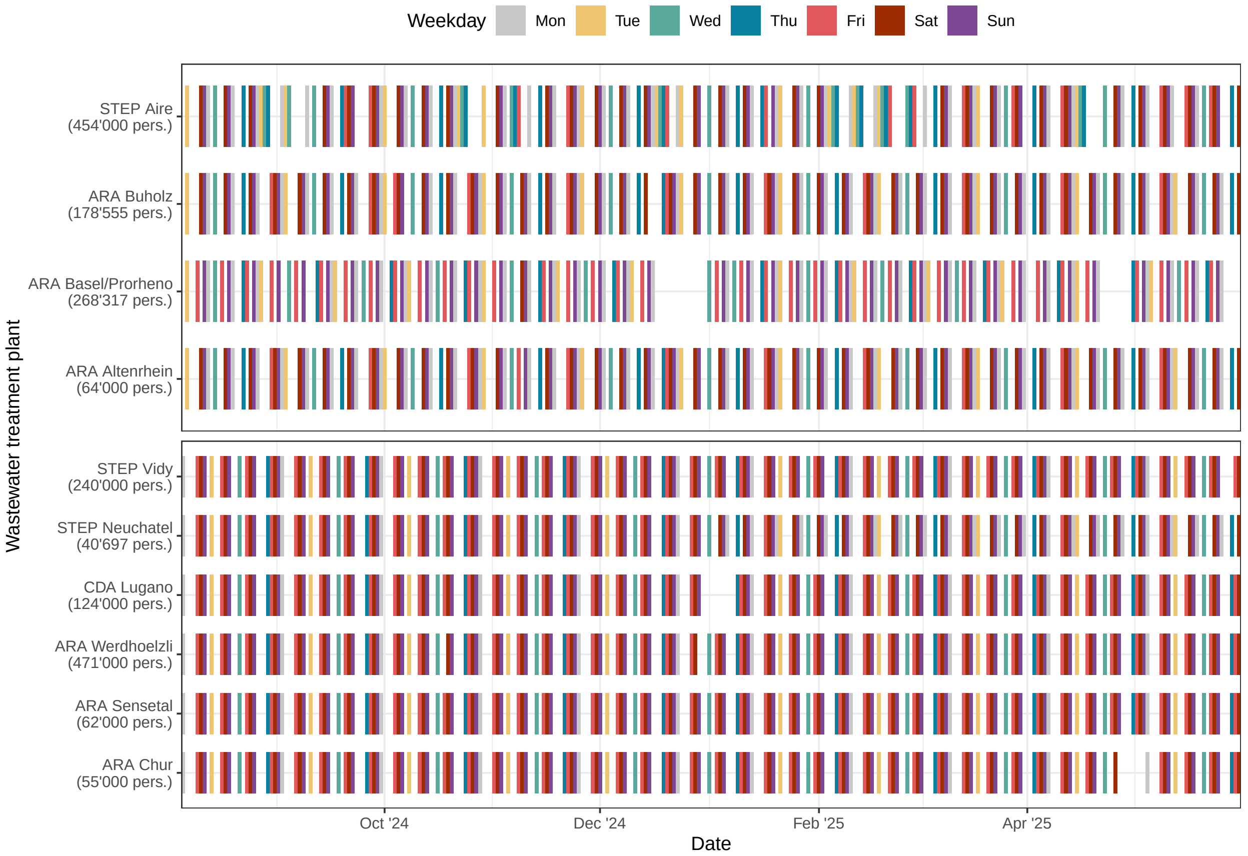

**Fig S3. Overview of analyzed samples from 10 wastewater treatment plants during the 2024/25 respiratory virus season in Switzerland.** As a rule, 4 samples per week from each treatment plant were analyzed. The specific weekdays analyzed varied between batch groups and alternated on a biweekly basis. For each sample, the concentration of SARS-CoV-2 (N1 and N2 genes), Influenza A and B virus (M gene), Respiratory syncytial virus (N gene), and Murine Hepatitis Virus (M gene, internal control) was quantified in duplicate using a six-plex digital PCR assay. Only samples which passed quality control are shown.

#### B Modeling measurements from digital PCR

We model the conditional distribution of concentration measurements from a digital PCR reaction with $m$ independent partitions and a conversion factor $c$. The factor $c$ describes the expected number of gene copies per partition from one concentration unit of the original sample and depends both on the partition volume and on the relative difference in concentration between the original sample and the reaction mix, e. g. due to dilution and addition of reagents. We also assume that given $\lambda_t$, the expected concentration in our model, the concentration in the PCR reaction mix is a random variable $\Lambda_t^{\text{pre}}$ with mean $\lambda_t$ and a pre-PCR coefficient of variation $\nu_{\text{pre}}$. The coefficient $\nu_{\text{pre}}$ describes all unexplained variation in target concentrations before the PCR quantification, including noise from wastewater sampling and from extraction and preprocessing.

##### B.1 Probability of non-detection

From the Poisson count statistics of positive partitions in digital PCR, it can be shown [3] that under the above assumptions, the probability of non-detection, i. e. a zero measurement, is

$$p_{\text{zero}}(\lambda_t) = \mathbb{E}[\exp(-\Lambda_t^{\text{pre}} cmn)], \tag{1}$$

where $n$ is the number of technical replicates of the sample. The expected value $\mathbb{E}[\exp(-\Lambda_t^{\text{pre}} cmn)]$ can be obtained from the moment-generating function (MGF) of $\Lambda_t^{\text{pre}}$. Here we assume that the pre-PCR variation is Log-Normal distributed, and use an approximation of the MGF of the Log-Normal distribution by Asmussen et al. [4], i. e.

$$\mathbb{E}[\exp(-\Lambda_t^{\text{pre}} cmn)] \approx \frac{1}{\sqrt{1+w}} \exp\left(-\frac{(w^2+2w)}{2\log(1+\nu_{\text{pre}}^2)}\right), \tag{2}$$

where $w$ is an evaluation of the Lambert $W$ function [5] with

$$w = W\left(-\lambda_t cmn \frac{\log(1+\nu_{\text{pre}}^2)}{\sqrt{1+\nu_{\text{pre}}^2}}\right). \tag{3}$$

As described in the main text, the probability of non-detection $p_{\text{zero}}(\lambda_t)$ is used in the hurdle model as the likelihood of zero measurements, and to adjust the distribution of non-zero measurements for the conditioning on detection.

##### B.2 Distribution of non-zero measurements

We characterize the distribution of non-zero measurements $\hat{\Lambda}_t$ by the mean and relative variance conditional on detection. As we assume an unbiased assay, the conditional mean of non-zero measurements is

$$\mathbb{E}\left[\hat{\Lambda}_t \middle| \hat{\Lambda}_t > 0\right] = \frac{\mathbb{E}\left[\hat{\Lambda}_t\right]}{P(\hat{\Lambda}_t > 0)} = \frac{\mathbb{E}\left[\hat{\Lambda}_t\right]}{1 - p_{\text{zero}}(\lambda_t)} = \frac{\lambda_t}{1 - p_{\text{zero}}(\lambda_t)}. \tag{4}$$

Moreover, the unconditional variance of concentrations averaged from $n$ technical replicates is

$$\text{Var}[\hat{\Lambda}_t] = \lambda_t^2 \nu_{\text{pre}}^2 + \frac{1}{c^2 mn} \left(\mathbb{E}[\exp(\Lambda_t^{\text{pre}} c)] - 1\right), \tag{5}$$

where $E[\exp(\Lambda_t^{\text{pre}} c)]$ can again be obtained from the moment-generating function (MGF) of $\Lambda_t^{\text{pre}}$ [3]. For this, we again use the approximation of the Log-Normal MGF from Eq. (2), but now let

$$w = W\left(\lambda_t \, c \frac{\log(1 + \nu_{\text{pre}}^2)}{\sqrt{1 + \nu_{\text{pre}}^2}}\right). \tag{6}$$

Using the unconditional variance from Eq. (5), the conditional relative variance of non-zero measurements is [3]

$$\nu^2(\lambda_t) = \frac{\text{Var}[\hat{\Lambda}_t]}{\lambda_t^2}(1 - p_{\text{zero}}(\lambda_t)) - p_{\text{zero}}(\lambda_t). \tag{7}$$

Finally, to model the conditional distribution of non-zero measurements via a Gamma distribution, we obtain the relevant parameters from the conditional mean and relative variance, i. e. with shape $\alpha$ and rate $\beta$ given by

$$\alpha = \frac{\mathbb{E}\left[\hat{\Lambda}_t \middle| \hat{\Lambda}_t > 0\right]^2}{\text{Var}\left[\hat{\Lambda}_t \middle| \hat{\Lambda}_t > 0\right]} = \frac{1}{\nu^2(\lambda_t)}, \quad \beta = \frac{\mathbb{E}\left[\hat{\Lambda}_t \middle| \hat{\Lambda}_t > 0\right]}{\text{Var}\left[\hat{\Lambda}_t \middle| \hat{\Lambda}_t > 0\right]} = \frac{1 - p_{\text{zero}}(\lambda_t)}{\lambda_t \nu^2(\lambda_t)}, \tag{8}$$

which corresponds to a mean of $\mu = \frac{\lambda_t}{1 - p_{\text{zero}}(\lambda_t)}$ and coefficient of variation of $\nu = \nu(\lambda_t)$.

### C Priors and epidemiological assumptions

#### C.1 Digital PCR parameters

The dPCR-specific observation model described in Supplement B assumes a number of partitions $m$ and a conversion factor $c$. Although these parameters were known to us for our specific laboratory protocol, we used the generic priors described in Lison et al. [3] to mimic settings in which detailed laboratory information might not be available. We model the number of partitions $m_t$ for a given sample on day $t$ via the average number of partitions $\mu_m$ and the coefficient of partition number variation $\nu_m$ under approximately log-normal distributed partition loss as described in Lison et al. [3]. We also used a broad prior for the pre-PCR coefficient of variation $\nu_{\text{pre}}$. An overview of the priors chosen is given in Table S1.

| Parameter | Description | Prior | Details |
|---|---|---|---|
| $\mu_m$ | average number of partitions | $\text{Normal}^{+}(\mu = 20000, \sigma = 5000)$ | accommodates dPCR systems with between 10000 and 30000 partitions |
| $\nu_m$ | coefficient of partition number variation | $\text{Normal}^{+}(\mu = 0, \sigma = 0.05)$ | less than 10% variation in the number of partitions |
| $c$ | concentration conversion factor | $\text{Normal}^{+}\left(\mu = 1 \times 10^{-5}, \sigma = 4 \times 10^{-5}\right)$ | $c \ll 100 \times 10^{-6} = 1 \times 10^{-4}$ |
| $\nu_{\text{pre}}$ | pre-PCR coefficient of variation | $\text{Normal}^{+}(\mu = 0, \sigma = 1)$ | weakly informative prior, less than 200% variation |

**Table S1. Overview of priors for the dPCR-specific model.** Shown are priors for the parameters of the digital PCR model described in Supplement B.

#### C.2 Generation time and shedding load distributions

Table S2 provides an overview of the generation time and shedding load distributions assumed for SARS-CoV-2, Influenza A, and RSV. In our discrete-time renewal model, the generation time has a minimum of 1 day. Thus, to represent the assumed, Log-Normal distributed generation time of SARS-CoV-2, we discretized the probability mass between 0 and 2 to a generation time of 1 day. For the Gamma distributed generation time of IAV and RSV, we used the shifted Gamma distribution as proposed in Cori et al. [6], which corresponds to a Gamma random variate shifted by one day. In contrast, the shedding load distributions for SARS-CoV-2, IAV, and RSV were discretized as $0, 1, 2, \ldots$ days after infection, since shedding can also start on the day of infection in our model.

The maximum generation time $G$ and shedding delay $S$ were chosen heuristically to cover at least 99.5% of the probability mass of the provided prior.

To model uncertainty of the shedding load distribution, we placed priors on the mean $\mu^{\text{shed}}$ and coefficient of variation $\nu^{\text{shed}}$ of the distribution. To construct the priors, we extracted parameter estimates with uncertainty information from the literature. For SARS-CoV-2, we used results from an observational study by Cavany et al. [7], where the shedding load distribution was inferred using a Bayesian model of wastewater and case surveillance data. We used two sets of normal priors to account for the range of possible parameters found by the authors in a sensitivity analysis of the testing delay. These priors represent the posterior distributions of parameters for an assumed testing delay of 0 and 5 days, respectively. We then modeled $\mu^{\text{shed}}$ and $\nu^{\text{shed}}$ as weighted means of the parameters for a delay of 0 and 5 days, with the weights $\boldsymbol{\theta}$ given by a Dirichlet prior. Here, we used a symmetric Dirichlet prior with concentration parameter $a = 1$, corresponding to a uniform distribution over the weight simplex. This means that we equally support parameters in between the two extremes found in the sensitivity analyses by Cavany et al. [7]. By choosing a smaller concentration parameter, this assumption can be modified towards a more isolated support of the extremes, which may be more appropriate in other cases. We also note that this approach generalizes to more than two sets of priors, and allows for differential weighting of results from different studies through the parameters of the Dirichlet prior. For IAV, we used a Gamma viral load distribution from Nadeau et al. [2] that was fitted to results from a meta-analysis of human volunteer challenge studies by Carrat et al. [8], based on nasal wash samples. We used the estimated parameters and associated standard errors of the model fitted by [2] to construct normal priors for $\mu^{\text{shed}}$ and $\nu^{\text{shed}}$. For RSV, we used results from a household study based on nasal swab samples by Otomaru et al. [9], and constructed normal priors for $\mu^{\text{shed}}$ and $\nu^{\text{shed}}$ based on the means and credible intervals of parameters of a Gamma distribution fitted by the authors. We note that when constructing priors for the mean and coefficient of variation of the Gamma distribution, we were not able to account for the correlation between the original shape and rate parameters estimated by the respective studies.

##### C.3 Total shedding load per infection

We obtained site-specific estimates for the average load per infection by relating a catchment's loads in the wastewater to the number of infections in that catchment. To estimate the number of infections

| Distribution | Pathogen | Assumption | Details |
|---|---|---|---|
| Generation time | SARS-CoV-2 | Log-Normal$(\mu = 3.0, \sigma = 1.5)$ | Park et al. [10], Omicron generation time, growth-rate corrected within-household estimate |
| | IAV | $\text{Gamma}^{\rightarrow}(\mu = 2.6, \sigma = 1.7)$ | te Beest et al. [11] |
| | RSV | $\text{Gamma}^{\rightarrow}(\mu = 7.5, \sigma = 2.1)$ | Vink et al. [12], serial interval (originally normal distributed), data from Crowcroft et al. [13] |
| Shedding load | SARS-CoV-2 | $\text{Gamma}(\mu = \boldsymbol{\theta} \cdot \boldsymbol{\mu}^{\text{shed}}, \nu = \boldsymbol{\theta} \cdot \boldsymbol{\nu}^{\text{shed}})$<br>$\boldsymbol{\mu}^{\text{shed}} \sim N\left((9.16, 15.69)^T, \text{diag}(0.83^2, 0.79^2)\right)$<br>$\boldsymbol{\nu}^{\text{shed}} \sim N\left((0.87, 0.53)^T, \text{diag}(0.06^2, 0.04^2)\right)$<br>$\boldsymbol{\theta} \sim \text{Dirichlet}(1, 1)$ | Cavany et al. [7], observational study, range of sensitivity analysis assuming a testing delay of 0–5 days |
| | IAV | $\text{Gamma}(\mu = \mu^{\text{shed}}, \nu = \nu^{\text{shed}})$<br>$\mu^{\text{shed}} \sim N\left(2.50, 0.19^2\right)$<br>$\nu^{\text{shed}} \sim N\left(0.34, 0.01^2\right)$ | Nadeau et al. [2], parametric estimate using empirical distribution from Carrat et al. [8], nasal washing |
| | RSV | $\text{Gamma}(\mu = \mu^{\text{shed}}, \nu = \nu^{\text{shed}})$<br>$\mu^{\text{shed}} \sim N\left(6.76, 1.17^2\right)$<br>$\nu^{\text{shed}} \sim N\left(0.35, 0.05^2\right)$ | Otomaru et al. [9], nasal swabs |

**Table S2. Overview of assumed generation time and shedding load distributions.** Note that for easier comparison, we here parameterize the Log-Normal and Gamma distribution by their mean $\mu$ and standard deviation $\sigma$ or coefficient of variation $\nu$. $\text{Gamma}^{\rightarrow}$ denotes the shifted Gamma distribution, as proposed in Cori et al. [6]. If $X$ is shifted Gamma distributed with some parameters, then $X - 1$ is Gamma distributed with identical parameters. $\boldsymbol{\theta}$, $\boldsymbol{\mu}^{\text{shed}}$, and $\boldsymbol{\nu}^{\text{shed}}$ denote vectors. Note that the generation time and shedding load distributions are discretized as $\tau^{\text{gen}} = (\tau_1^{\text{gen}}, \tau_2^{\text{gen}}, \ldots, \tau_G^{\text{gen}})$ and $\tau^{\text{shed}} = (\tau_0^{\text{shed}}, \tau_1^{\text{shed}}, \ldots, \tau_S^{\text{shed}})$ in our model.

in a catchment, we used data from the Swiss Sentinel Surveillance System (Sentinella). The data is publicly available from the Infectious Diseases Dashboard by the Swiss Federal Office of Public Health [14]. This system is based on voluntary contributions from approximately 170 enrolled general practitioners, internists and paediatricians, who report information on consultations of patients with acute respiratory infection (ARI, "acute onset of illness with cough, sore throat, shortness of breath or rhinitis AND of infectious origin, as judged by a physician") or influenza-like illness (ILI, "sudden onset of high fever (>38°C) and cough or sore throat") in medical practices and during home visits. Nasopharyngeal swabs are taken from a subset of consulted patients with acute respiratory infection, and tested at the National Reference Centre for Influenza (NRCI) for various respiratory pathogens, including SARS-CoV-2, Influenza A and B virus, and RSV. From the ratio of the number of positive tests for a given pathogen $j$ to the total number of tests, the weekly proportion $p_w^{\mathrm{j}}$ of consulted patients with ARI that are infected with the pathogen can be estimated. In addition, using the reported numbers of ARI consultations, the total number of consultations, reported age and sex distributions of consulted patients, and general population statistics in Switzerland, the Federal Office of Public Health (FOPH) projects a weekly incidence of ARI-related consultations $c_w^{\mathrm{ARI}}$ for the Swiss population. Finally, we used estimates based on syndromic reports by the participatory surveillance system "Grippenet Suisse" [15] for the incidence of influenza-like illness infections in Switzerland as a conservative proxy (subset of ARI infections) for the under-ascertainment of acute respiratory infections in Switzerland. For each flu season, we estimated the underreporting factor as $\rho = \frac{\sum_w I_w^{\mathrm{Grippenet}}}{\sum_w c_w^{\mathrm{ARI}}}$, where $I_w^{\mathrm{Grippenet}}$ is the estimated ILI incidence from Grippenet and $c_w^{\mathrm{ARI}}$ the number of ARI consultations in week $w$ of the season, respectively. Assuming that consultation is independent of the probability of infection with a pathogen, we then estimated the total number of infections with pathogen $j$ in week $w$ in the Swiss population as $I_w^j = p_w^{\mathrm{j}} \times c_w^{\mathrm{ARI}} \times \rho$. Using the population size $n_k$ of a catchment $k$ relative to the Swiss population size $n = 8\,770\,000$, we obtained a crude estimate of the weekly number of infections with the pathogen in that catchment, $I_{w,k}^j = I_w^j \times \frac{n_k}{n}$. For each week, we divided the sum of measured loads on all days $t \in T_w$ of the week by the weekly number of infections to obtain a weekly estimate of the load per infection $\hat{\mu}_w^{\mathrm{load}} = \frac{\sum_{t \in T_w} c_t \times \mathrm{flow}_t}{I_{w,k}^j}$. We found the estimated weekly load per infection to be relatively stable over time and during seasonal waves. For our model parameter $\mu^{\mathrm{load}}$, we used the median of weekly estimated average loads per infection from the first weeks of the flu season before December 1.

#### C.4 Outliers

The parameter $\epsilon_t$ represents spikes in sample concentrations that strongly deviate from the concentration predicted by our model under the assumption of uniform mixing. While there are various potential explanations for such spikes, including unrepresentative sampling and sewer-level effects [16], we here use a phenomenological approach. We parameterized $\epsilon_t$ using a generalized extreme value distribution, i. e.

$$\epsilon_t \sim \text{GEV}(k\mu^\epsilon, k\sigma^\epsilon, \xi^\epsilon). \tag{9}$$

with scaling factor $k$, location $\mu^\epsilon = 0$, scale $\sigma^\epsilon = 2 \times 10^{-8}$, and shape $\xi^\epsilon = 4$. This yields a Type II/Fréchet extreme value distribution with a heavy right tail. As a scaling factor, we chose $k = \frac{\mu^{\text{load}}}{\text{med(flow)}}$ to approximate the expected sample concentration corresponding to only 1 infection in the catchment. The resulting prior assigns less than 1% probability to spikes larger than 1 infection ($\approx 3$ per year), approximately 0.5% to spikes larger than 10 infections ($\approx$ 1–2 per year), and still over 0.1% to spikes larger than 1000 infections ($\approx 0.5$ per year). These probabilities approximately match the observed frequency of outliers in our historical data.

#### C.5 Transmission dynamics

In the following, we provide further details on the priors used for the reproduction number $R_t$ and the seeding phase of the infection trajectory. As described in the main text, we smoothed $R_t$ using two approximate Gaussian processes with long- and short-term kernels, i. e.

$$\alpha \sim \mathcal{GP}(1, k_\nu(\tau; \ell_\alpha, \sigma_\alpha^2)),\ \delta \sim \mathcal{GP}(0, k_\nu(\tau; \ell_\delta, \sigma_\delta^2)), \tag{10}$$

see Supplement D for details on the approximation. By decomposing $R_t$ into long- and short-term components, we obtain desirable behavior in real-time estimation, where transmission dynamics are only partially informed by wastewater data due to shedding delays. In the absence of signal, the short-term deviation component reverts to its mean of zero, causing $R_t$ to follow the long-term trend $\alpha_t$. Although $\alpha_t$ also reverts to its prior mean of $R_t = 1$ over long time scales, this effect is negligible for real-time estimation and short-term forecasts, as the long kernel length scale enforces strong correlation between consecutive $R_t$ values. Thus, in expectation, $R_t$ remains at its current level near the present, representing a minimally informed assumption of stable transmission dynamics when wastewater data provide no contrary evidence. Moreover, through the use of an inverse softplus link $g(R_t) = \alpha_t + \delta_t$, we obtain a prior for $R_t$ that is approximately symmetric with respect to changes in transmission. This is not given for other popular link functions such as the logarithmic function, which e. g. favors a decrease from $R_t = 2$ to $R_t = 1$ over an increase from $R_t = 1$ to $R_t = 2$. In contrast, the softplus function closely approximates the identity function for practically relevant values of $R_t$ when a high scaling factor ($k = 4$) is used.

For the seeding phase, during which $R_t$ cannot be calculated, we let the expected number of infections $\iota_t$ follow an exponential growth process, where $\iota_1$ is the initial number of infections. We used a broad prior for $\iota_1$, informed by a rough empirical estimate of the initial number of infections to ensure that the seeding phase is modeled on the right order of magnitude. Specifically, we used the supplied wastewater measurements and shedding assumptions to approximately back-calculate the initial number of infections and then constructed a truncated normal prior with parameters chosen such that the 5% and 95% of the distribution correspond approximately to 1/10 and 10 times the empirical estimate of the number of infections. The exponential growth rate $r_t$ during the seeding phase was modeled via a backward-in-time random walk with starting value $r_{G+1}$ and standard deviation $\sigma_r$, i. e.

$$r_t | r_{t+1} \sim N(r_{t+1}, {\sigma_r}^2) \mid 1 < t \leq G. \tag{11}$$

We obtain $r_{G+1}$ from the reproduction number $R_{G+1}$ using the relationship $R_{G+1}(\sum_{s=1}^{G} \tau_s^{\text{gen}} e^{-s r_{G+1}}) = 1$, which we solve numerically for $r_{G+1}$ via root-finding with the Newton–Raphson method. For $\sigma_r$, we used a $\text{Normal}^+(\mu = 0, \sigma = 0.01)$ prior, allowing the growth rate to vary by up to $\approx \pm 4$ percentage points per day. This approach models the initial $R_t$ values as a continuation of an exponential growth trend, while allowing us to express the expected growth rates during the seeding phase via a prior for the initial reproduction number $R_{G+1}$.

To compute the growth rate for $t > G$, we used the estimator

$$r_t = \frac{d \log \Lambda_{t+\bar{\tau}}}{dt}, \quad \Lambda_t = \sum_{s=1}^{G} \tau_s^{\text{gen}} \iota_{t-s}, \tag{12}$$

which corresponds to the logarithmic derivative of the total expected infectiousness $\Lambda_t$, shifted backward in time by the mean generation time $\bar{\tau}$. This estimator was proposed by Parag et al. [17] as a more stable alternative to the logarithmic derivative of new infections.

#### D Estimation

We implemented our Bayesian hierarchical model in the probabilistic programming language "stan" [18] to allow sampling from the posterior distribution of parameters via Hamiltonian Monte Carlo. As sampling of discrete parameters is not supported in stan, we used continuous approximations for the realized number of infections, where $I_t$ is modeled as Normal distributed with the mean and standard deviation of a Poisson variate [19–22], i. e.

$$I_t|\iota_t \sim \text{Normal}(\mu = \iota_t, \sigma = \sqrt{\iota_t}). \tag{13}$$

To improve sampling efficiency, we also approximated the distribution for $\zeta_t \sim \text{Gamma}(\alpha = \frac{I_t}{\nu_\zeta^2}, \beta = \frac{1}{\nu_\zeta^2})$ using a Normal distribution with the same mean and standard deviation when the shape parameter of the Gamma distribution was $\alpha \geq 30$.

To sample from the Gaussian processes used as smoothing priors for $R_t$, we implemented a basis function approximation using Laplace eigenfunctions described in Riutort-Mayol et al. [23, 24]. This approach was implemented in a similar fashion by Abbott et al. for estimating $R_t$ from case count data [25]. In our implementation, we used a boundary factor of $c = 3$, which is higher than the minimum recommended by Riutort-Mayol et al. to ensure accurate real-time estimates, i. e. also near the boundaries. Moreover, based on recommendations in Riutort-Mayol et al., we used $m = 3.42\,c\,\frac{S}{l}$ different basis functions, where $l$ is the length scale of the Gaussian process (we used the 5% quantile of the length scale prior for a conservative result), and $S$ is the maximum absolute value of the zero-centered input space, which is $\frac{n-1}{2}$, where $n$ is the number of $R_t$ time steps modeled.

We generally used non-centered parameterizations of normally distributed nuisance parameters such as random walk increments. Where possible, we reparameterized parameters on the logarithmic scale to improve numerical stability.

For sampling, we ran the No-U-Turn Sampler (NUTS) implemented in cmdstan version 2.34.1 [18] using 4 parallel chains with 1000 warm-up and 2500 sampling iterations each, a maximum tree depth of 15, an initial step size of 0.01, and an adaptation target acceptance statistic of 0.99. For each fitted model, we assessed several diagnostics of the NUTS sampler, specifically the number of divergent transitions [26] and the Bayesian fraction of missing information [27]. The majority of fitted models had no divergent transitions (Figure S4). We found divergent transitions for model fits on some days, in particular when fitting models during the off-season 2022/23, which included a two-week processing break due to the switch from a four-plex to a six-plex assay. However, we did not observe a noticeable difference between transmission dynamic estimates on days with and without divergent transitions. This suggests that small numbers of divergent transitions might be acceptable for our application [28], but if highly accurate estimates are important, we suggest running the model with a higher adaptation target acceptance statistic. The Bayesian fraction of missing information was consistently above the default threshold of 0.2 across model fits (Figure S5). We also checked the daily $R_t$ estimates via the Gelman-Rubin convergence diagnostic ($\hat{R} < 1.05$) [29] and for sufficient effective sample sizes (ESS $>$ 400) [30]. $\hat{R}$ values were below 1.05 for most model fits, although there were some outliers with higher $\hat{R}$ values on a small number of dates (Figure S6). Effective sample sizes were consistently above 400 for both the bulk and tail of the distribution, again with the exception of a small number of dates that had a reduced bulk effective sample size (Figures S7 and S8).

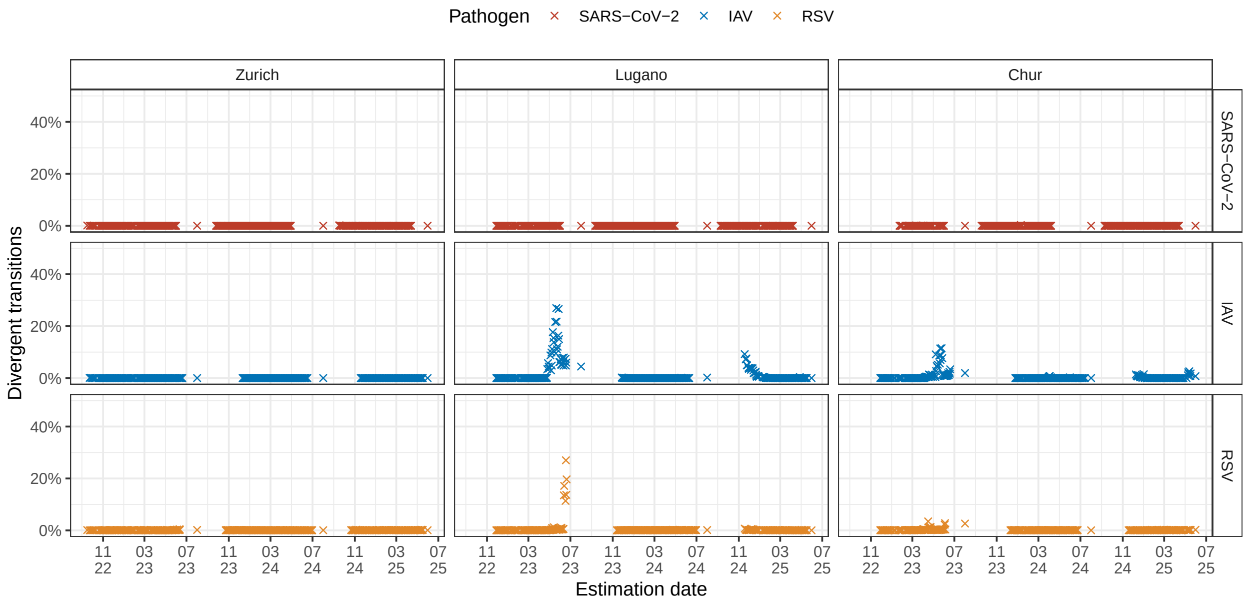

**Fig S4. Percentage of divergent transitions per fitted model.** Boxplots show the share of divergent transitions out of 10 000 sampling iterations for all real-time model fits for SARS-CoV-2, IAV, and RSV, across catchments and seasons.

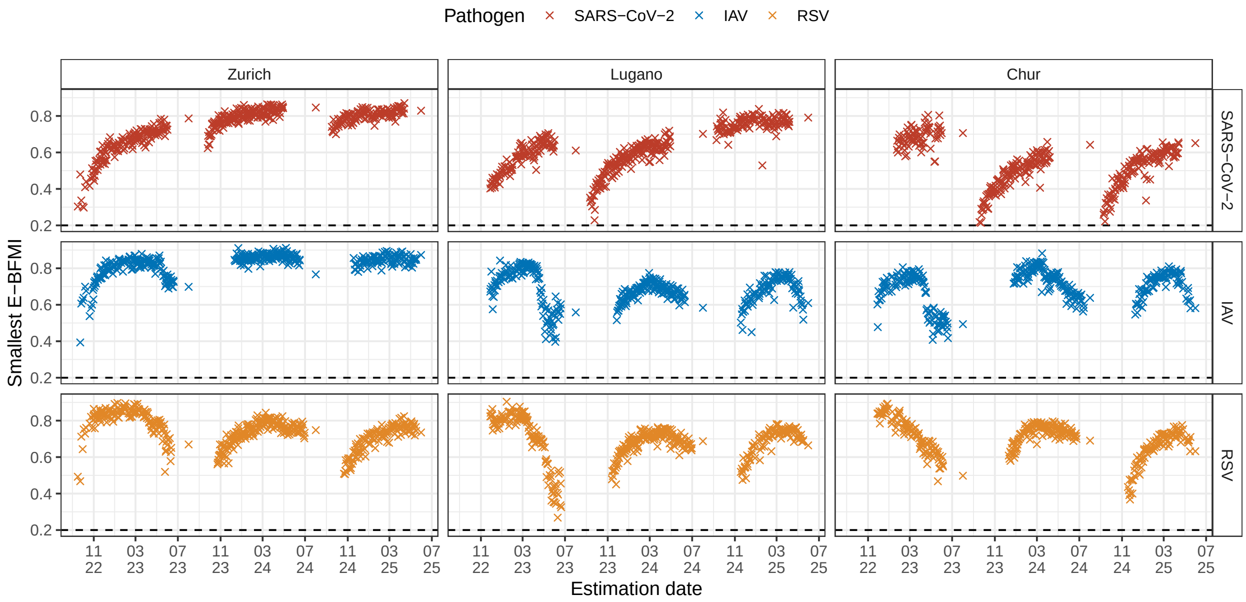

**Fig S5. Smallest E-BFMI per fitted model.** Boxplots show the smallest E-BFMI out of 4 chains for all real-time model fits for SARS-CoV-2, IAV, and RSV, across catchments and seasons. E-BFMI values above 0.2 are considered sufficient.

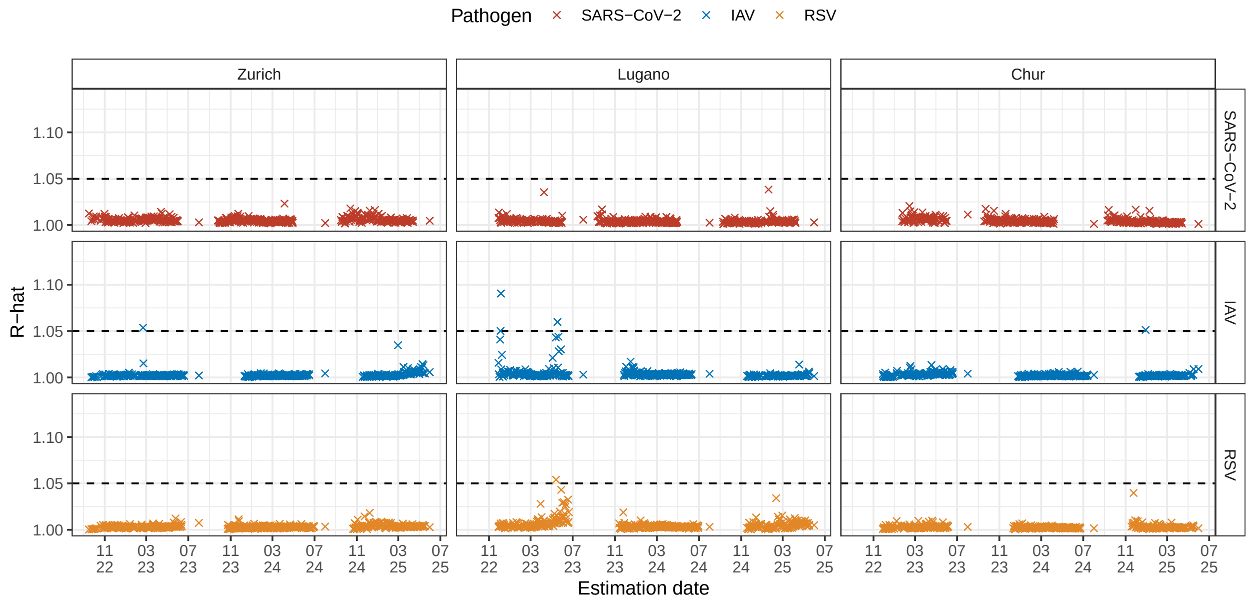

**Fig S6. Gelman-Rubin convergence diagnostic ($\hat{R}$) per fitted model.** Boxplots show the average $\hat{R}$ value of $R_t$ estimates for all real-time model fits for SARS-CoV-2, IAV, and RSV, across catchments and seasons Values of $\hat{R} < 1.05$ are considered sufficient.

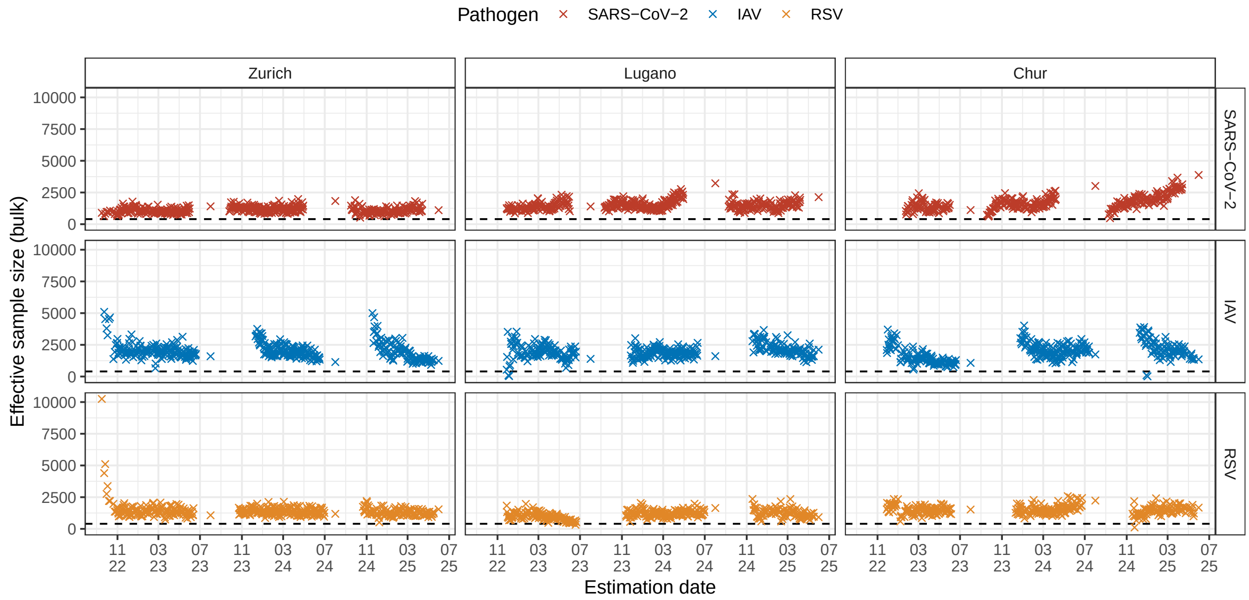

**Fig S7. Bulk effective sample size per fitted model.** Boxplots show the average bulk effective sample size of $R_t$ estimates for all real-time model fits for SARS-CoV-2, IAV, and RSV, across catchments and seasons. An effective sample size above 100 per Markov chain is considered sufficient.

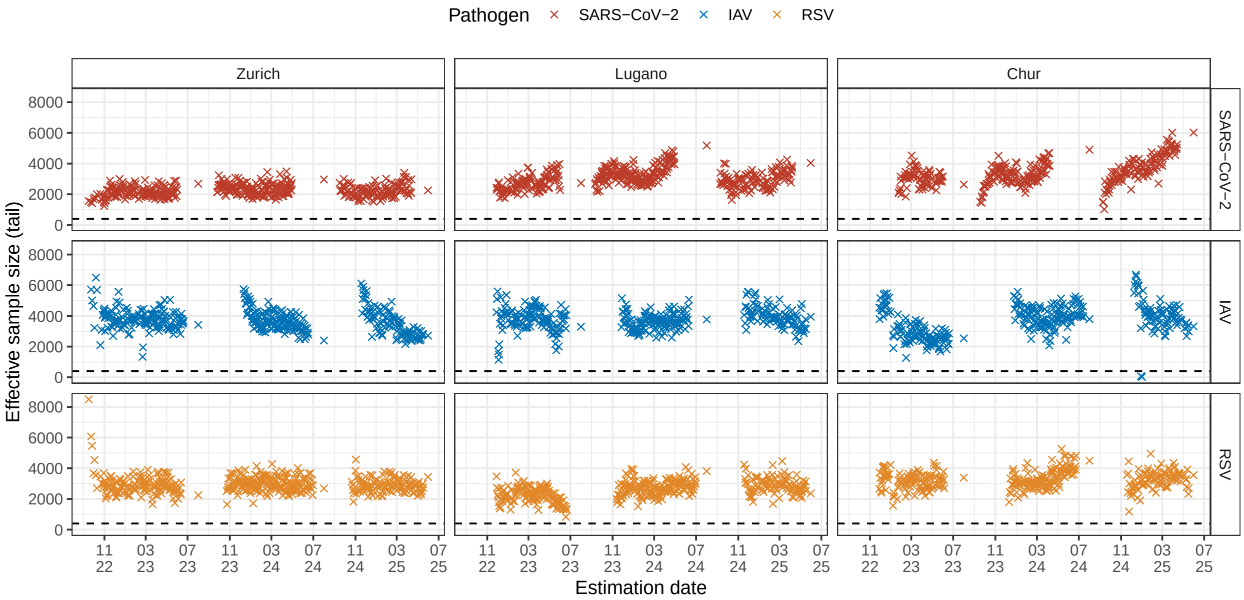

**Fig S8. Tail effective sample size per fitted model.** Boxplots show the average tail effective sample size of $R_t$ estimates for all real-time model fits for SARS-CoV-2, IAV, and RSV, across catchments and seasons. An effective sample size above 100 per Markov chain is considered sufficient.

### E Sensitivity analyses

#### E.1 Flow data

Figures S9 and S10 show a comparison of models fitted with and without daily flow data to measurements of SARS-CoV-2 concentrations at the WWTPs of Zurich and Lugano. When no daily flow data was used, we assumed a constant flow equal to the median value. As can be seen, the estimated wave for SARS-CoV-2 in Zurich had a lower peak and a smaller $R_t$ when no flow data was used, because the higher flow rates during the winter season were not taken into account (Figure S9). In contrast, the estimated wave in Lugano had a higher peak and a higher $R_t$ when no flow data was used, as Lugano had lower flow rates during the winter season (Figure S10).

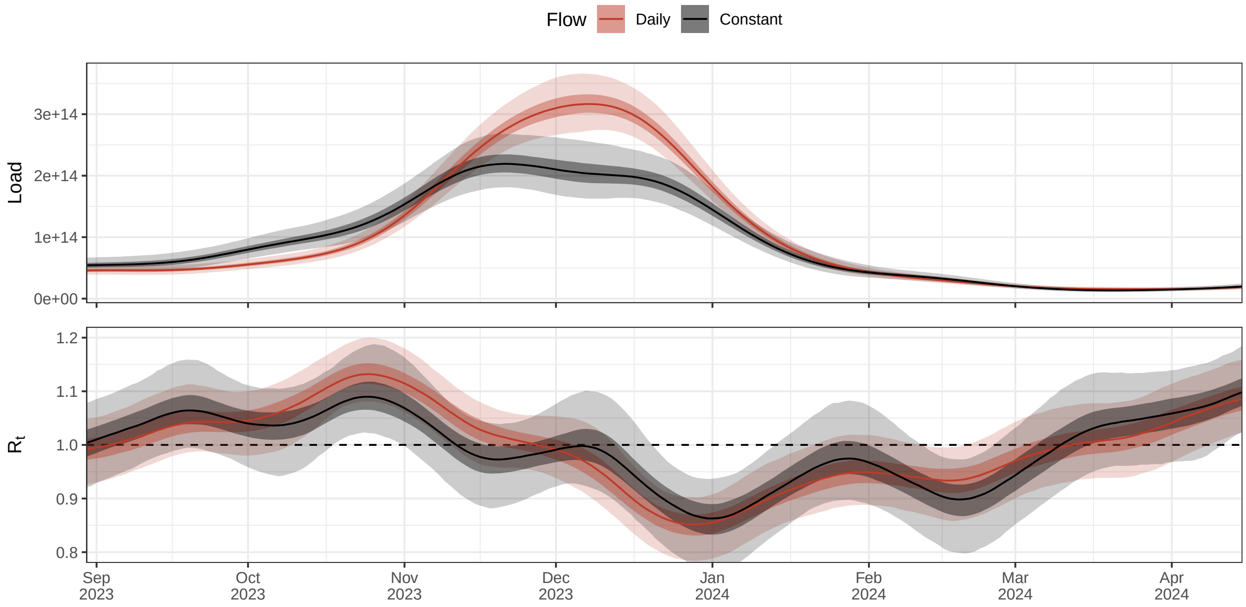

**Fig S9. Comparison of SARS-CoV-2 wave in Zurich estimated with and without flow data.** Shown is the estimated total daily load in wastewater (top) and the effective reproduction number (bottom) for SARS-CoV-2 during the winter season 2023/24 in Zurich, estimated i) using daily flow data (red) and ii) assuming a constant flow (black). Lines indicate the posterior median and dark and light bands the 50% and 95% credible intervals of the marginal posterior distribution, respectively.

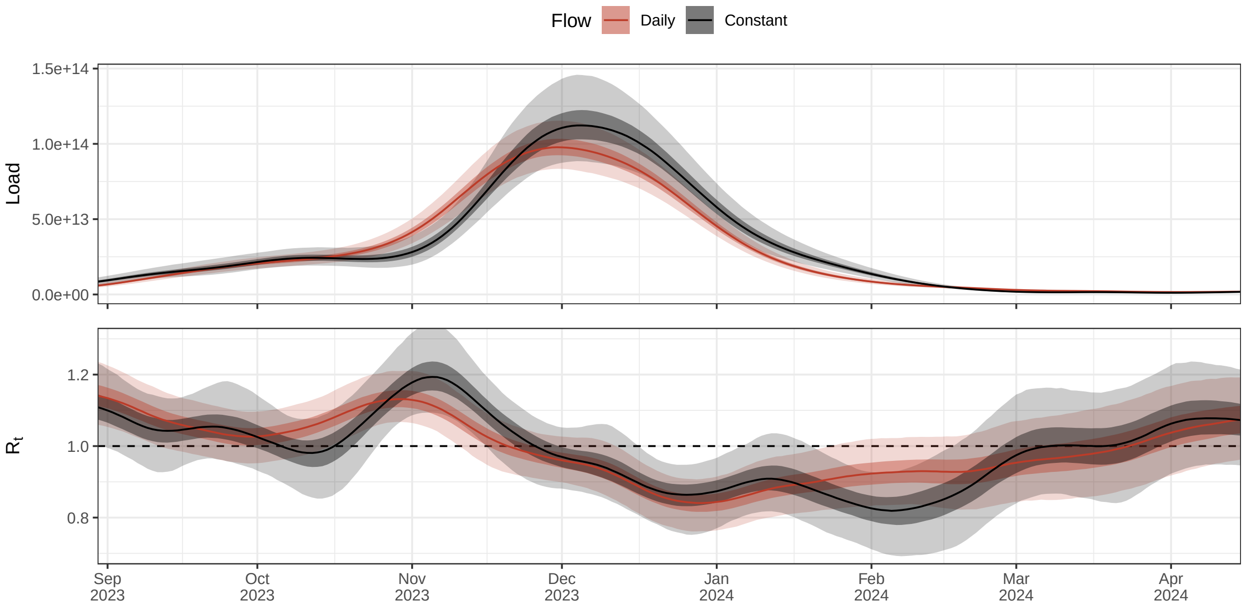

**Fig S10. Comparison of SARS-CoV-2 wave in Lugano estimated with and without flow data.** Shown is the estimated total daily load in wastewater (top) and the effective reproduction number (bottom) for SARS-CoV-2 during the winter season 2023/24 in Zurich, estimated i) using daily flow data (red) and ii) assuming a constant flow (black). Lines indicate the posterior median and dark and light bands the 50% and 95% credible intervals of the marginal posterior distribution, respectively.

#### E.2 Outlier detection

Figures S11 and S12 show a comparison of effective reproduction number estimates obtained when either including outliers, as in the main text, or removing them before model fitting. As can be seen, both for SARS-CoV-2 in Chur and for RSV in Zurich, the removal of outliers had no relevant effect on the estimated $R_t$ trajectory. This suggests that our modeling of occasional, strong spikes in concentrations can successfully identify and isolate individual outliers, reducing their effect on the estimated transmission dynamics to near zero.

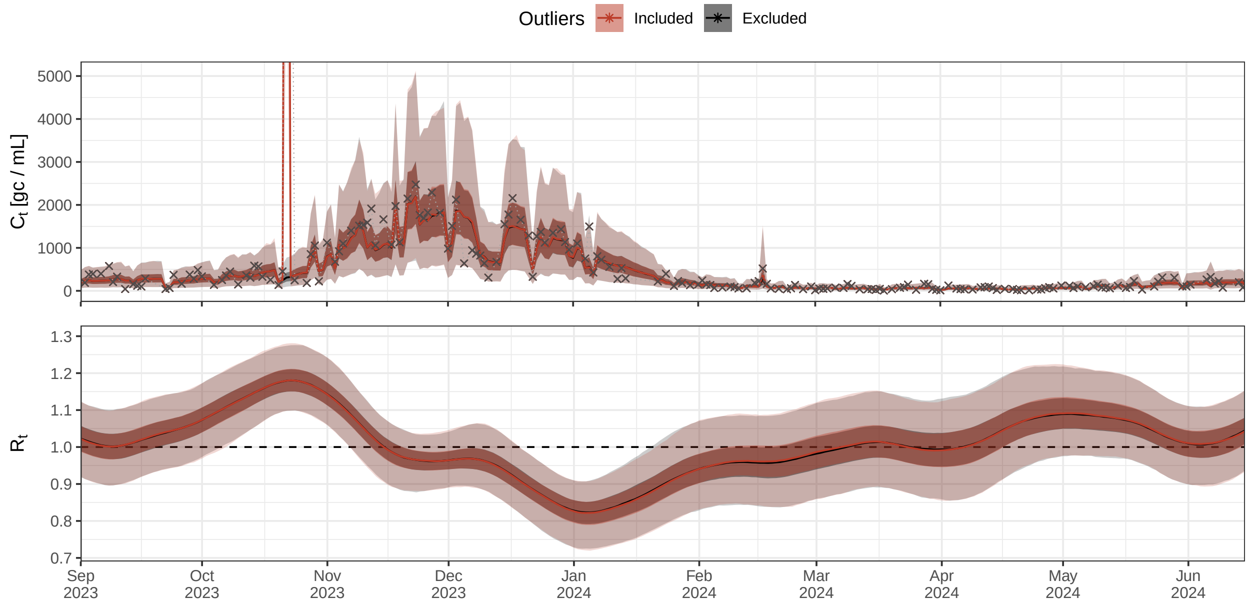

**Fig S11. Robustness of estimated transmission dynamics of SARS-CoV-2 in Chur to outlier measurements.** Shown is the model fit of the measured concentration in wastewater (top) and the estimated effective reproduction number (bottom) for SARS-CoV-2 during the winter season 2023/24 in Chur, estimated i) with outliers included, as in the main text (orange) and ii) with outliers removed (black). Lines indicate the posterior median and dark and light bands the 50% and 95% credible intervals of the marginal posterior distribution, respectively.

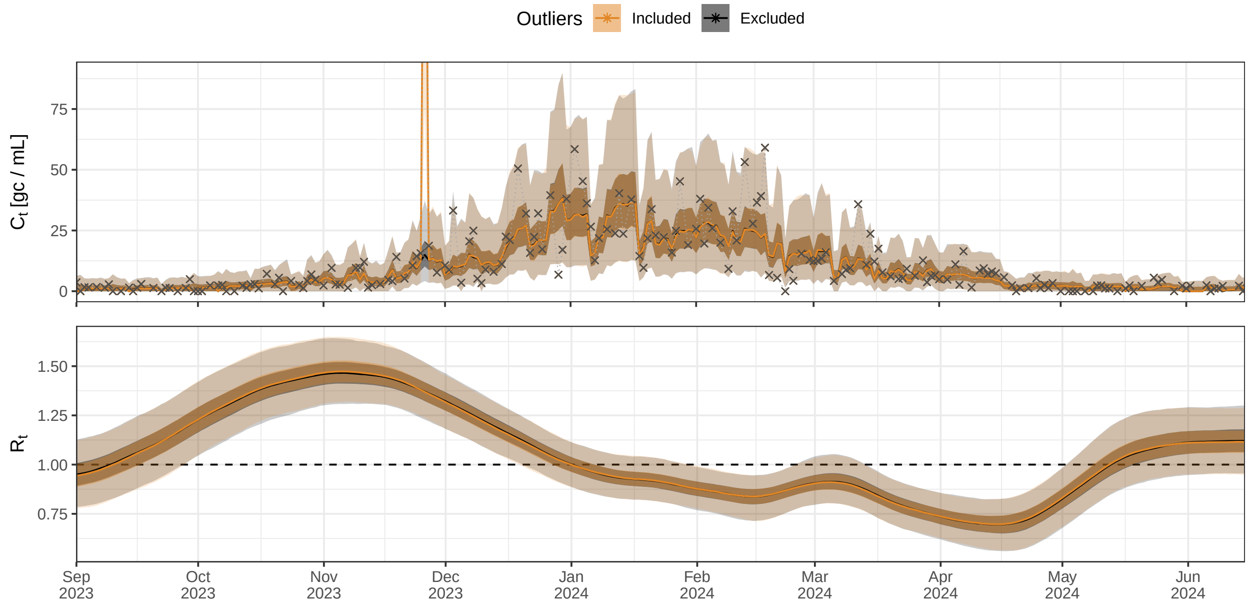

**Fig S12. Robustness of estimated transmission dynamics of RSV in Zurich to outlier measurements.** Shown is the model fit of the measured concentration in wastewater (top) and the estimated effective reproduction number (bottom) for RSV during the winter season 2023/24 in Zurich, estimated i) with outliers included, as in the main text (orange) and ii) with outliers removed (black). Lines indicate the posterior median and dark and light bands the 50% and 95% credible intervals of the marginal posterior distribution, respectively.

#### E.3 Shedding load

In our main analysis, we calibrated the average shedding load per infection using data from Sentinel and participatory surveillance in Switzerland (Supplement C.3). For comparison, Figures S13 to S15 show $R_t$ estimates based on a simpler calibration, where we assumed that the load detected at the start of each season (average of first three consecutive positive samples) corresponds to the total load shed by 0.01% of the catchment population. For most catchments and targets, the resulting $R_t$ estimates were practically identical to the main analysis. Notable exceptions were estimates for SARS-CoV-2 at the catchments of Lugano and Chur, which were more uncertain than in the main analysis, particularly during the winter seasons 2022/23 and 2024/25. We also conducted a systematic sensitivity analysis of the assumed total detectable load per infection using the example of SARS-CoV-2 in Zurich during the winter season 2023/24 (Figure S16). Here we find that estimated transmission parameters such as the effective reproduction number are in principle invariant to the assumed load per infection, which acts as a constant scaling factor for the number of infections in the population. We therefore obtain practically identical estimates in most settings. However, as our model also accounts for stochasticity in the infection process, estimates of $R_t$ become more uncertain when the infection incidence is very low. In the example shown in Figure S16, we find a notable difference in uncertainty when the estimated peak incidence is 20 instead of 2000, which corresponds to a regime with few infections that is considerably influenced by chance events. Overall, this means that misspecification of the load per infection is only relevant if the true incidence is very low (in this case, an underestimation of the load per infection could lead to an underestimation of $R_t$ uncertainty) or if the true incidence is high but the load per infection is massively overestimated (in this case, the estimated incidence would be too low, leading to overly uncertain $R_t$ estimates). Outside these cases, the assumed load per infection has little effect on $R_t$ estimates.

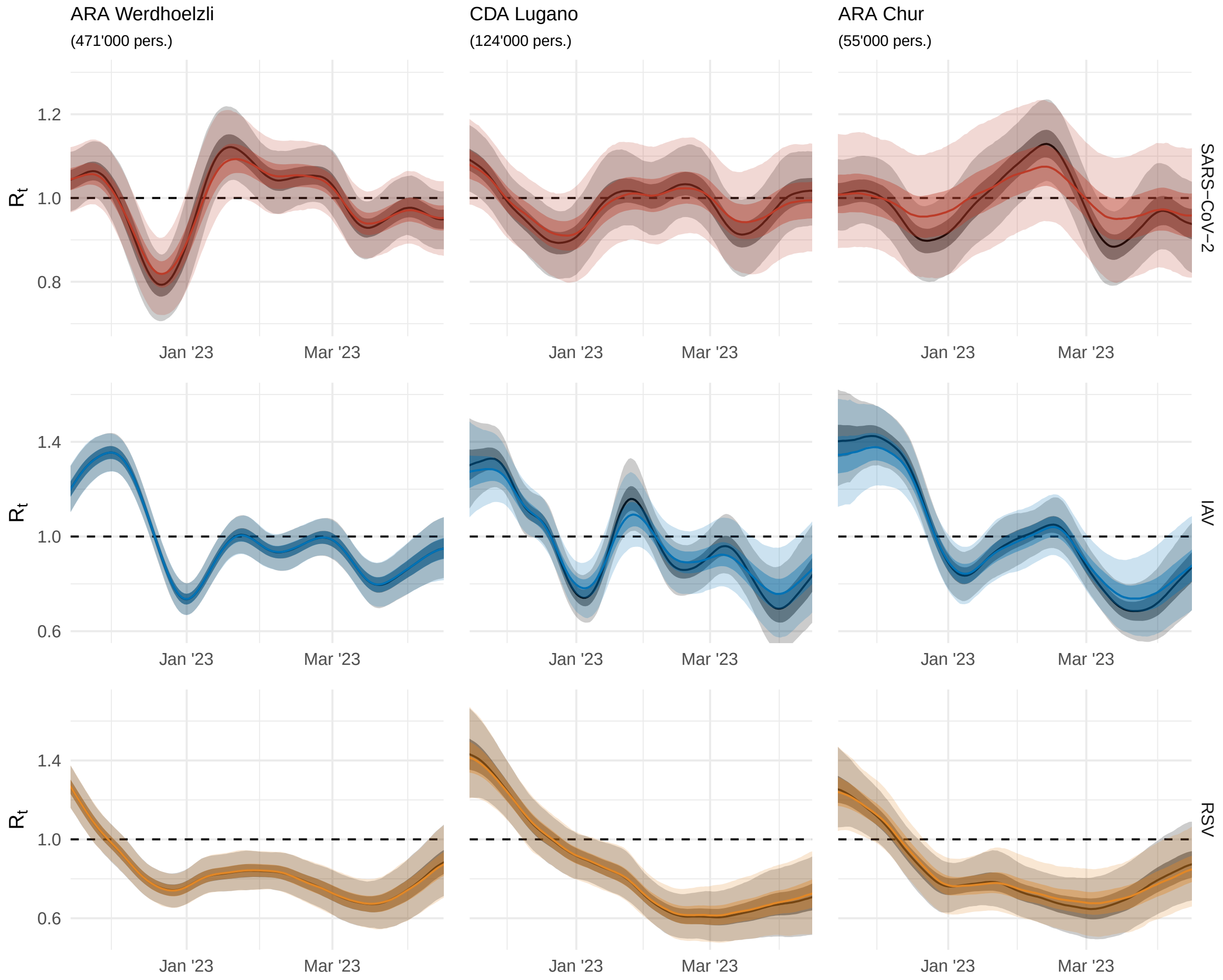

**Fig S13. $R_t$ estimates based on an alternative calibration of the shedding load per infection during the winter season 2022/23.** Estimated reproduction numbers of SARS-CoV-2, IAV, and RSV for the wastewater catchments of Zurich, Lugano, and Chur, Switzerland, during the winter season 2022/23 obtained when calibrating the total detectable shedding load per infection at the start of the season to 0.01% of the catchment population. For comparison, the corresponding $R_t$ estimates from the main analysis are shown in grey. Lines indicate the posterior median and dark and light bands the 50% and 95% credible intervals of the marginal posterior distribution, respectively.

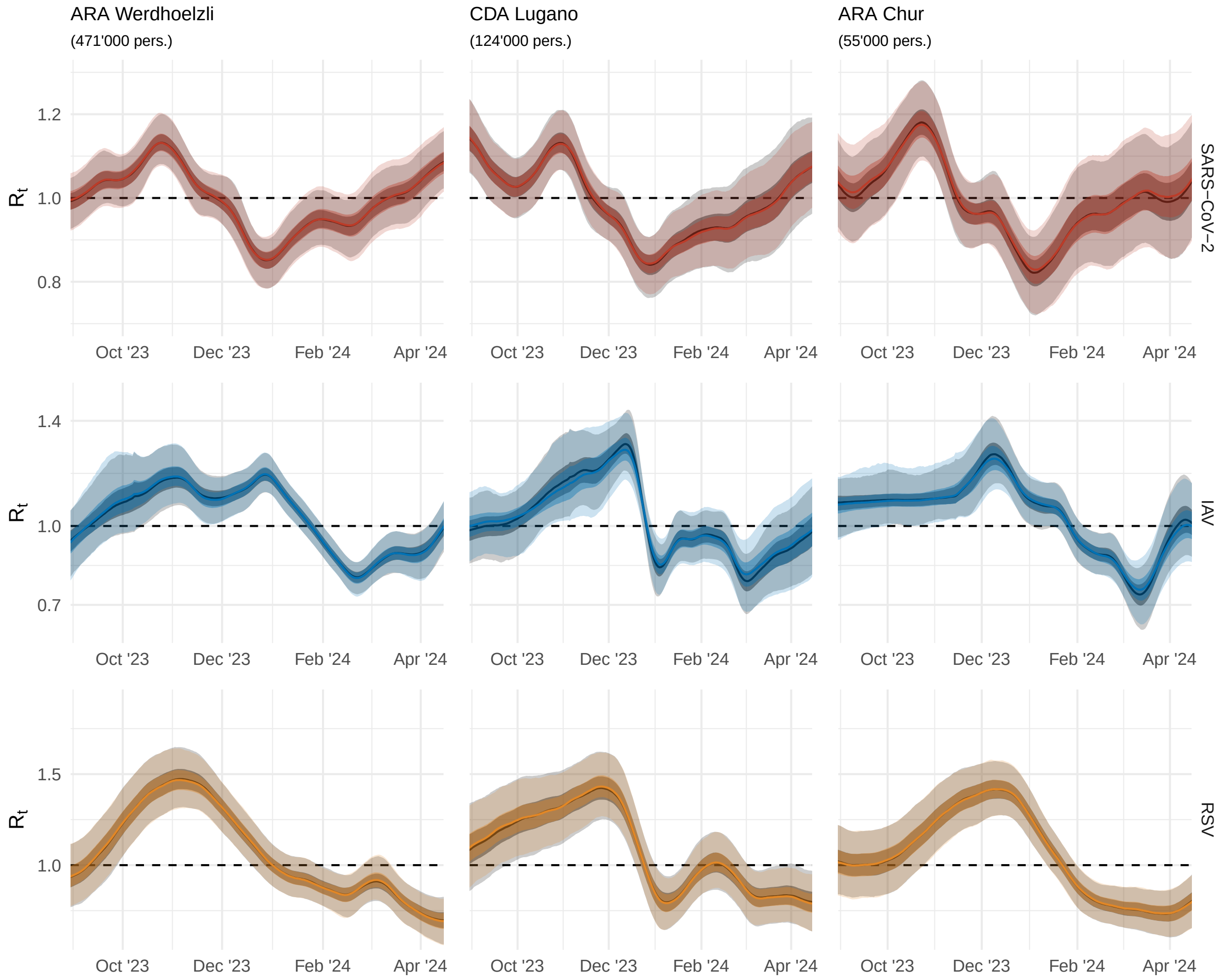

**Fig S14. $R_t$ estimates based on an alternative calibration of the shedding load per infection during the winter season 2023/24.** Estimated reproduction numbers of SARS-CoV-2, IAV, and RSV for the wastewater catchments of Zurich, Lugano, and Chur, Switzerland, during the winter season 2023/24 obtained when calibrating the total detectable shedding load per infection at the start of the season to 0.01% of the catchment population. For comparison, the corresponding $R_t$ estimates from the main analysis are shown in grey. Lines indicate the posterior median and dark and light bands the 50% and 95% credible intervals of the marginal posterior distribution, respectively.

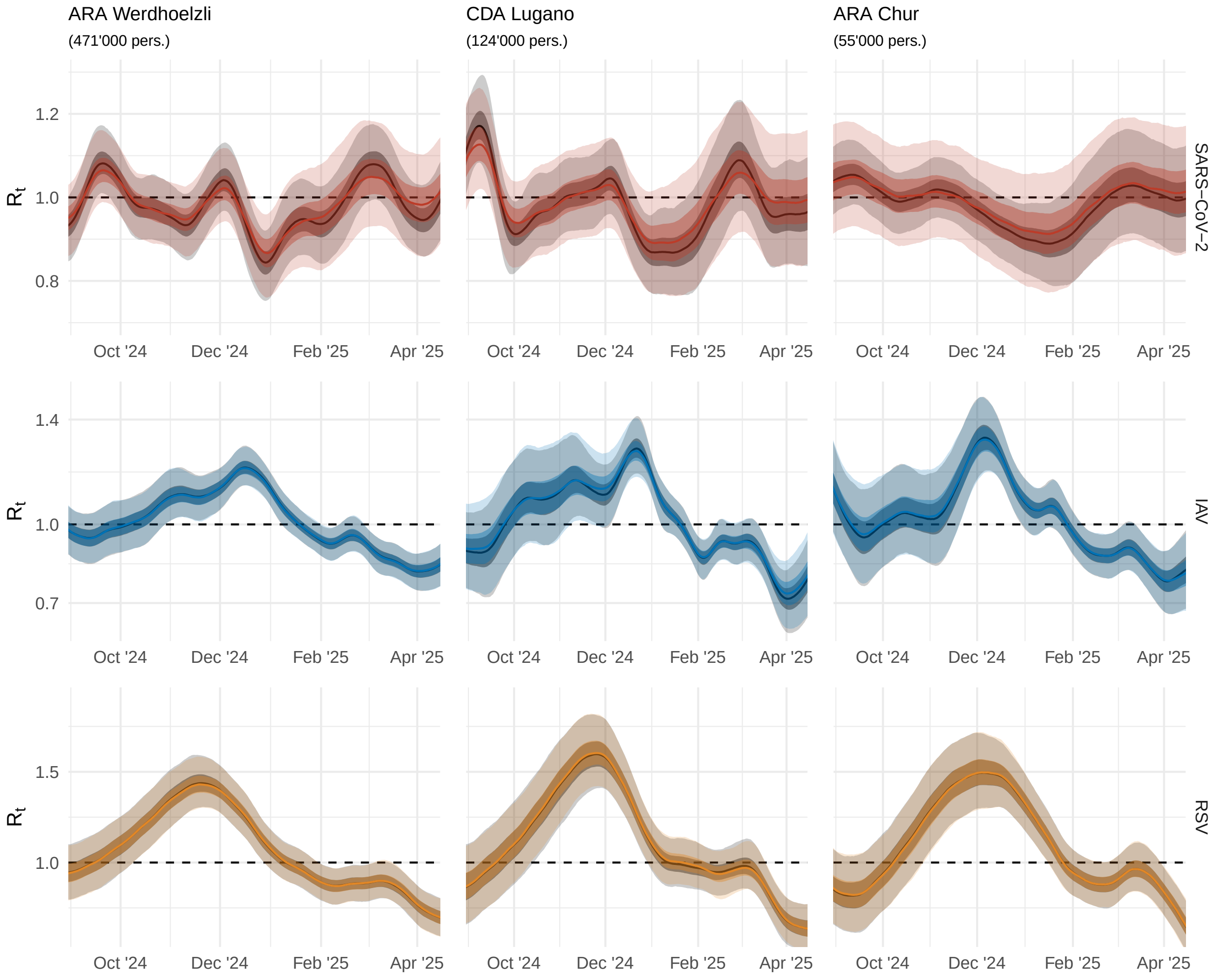

**Fig S15. $R_t$ estimates based on an alternative calibration of the shedding load per infection during the winter season 2024/25.** Estimated reproduction numbers of SARS-CoV-2, IAV, and RSV for the wastewater catchments of Zurich, Lugano, and Chur, Switzerland, during the winter season 2024/25 obtained when calibrating the total detectable shedding load per infection at the start of the season to 0.01% of the catchment population. For comparison, the corresponding $R_t$ estimates from the main analysis are shown in grey. Lines indicate the posterior median and dark and light bands the 50% and 95% credible intervals of the marginal posterior distribution, respectively.

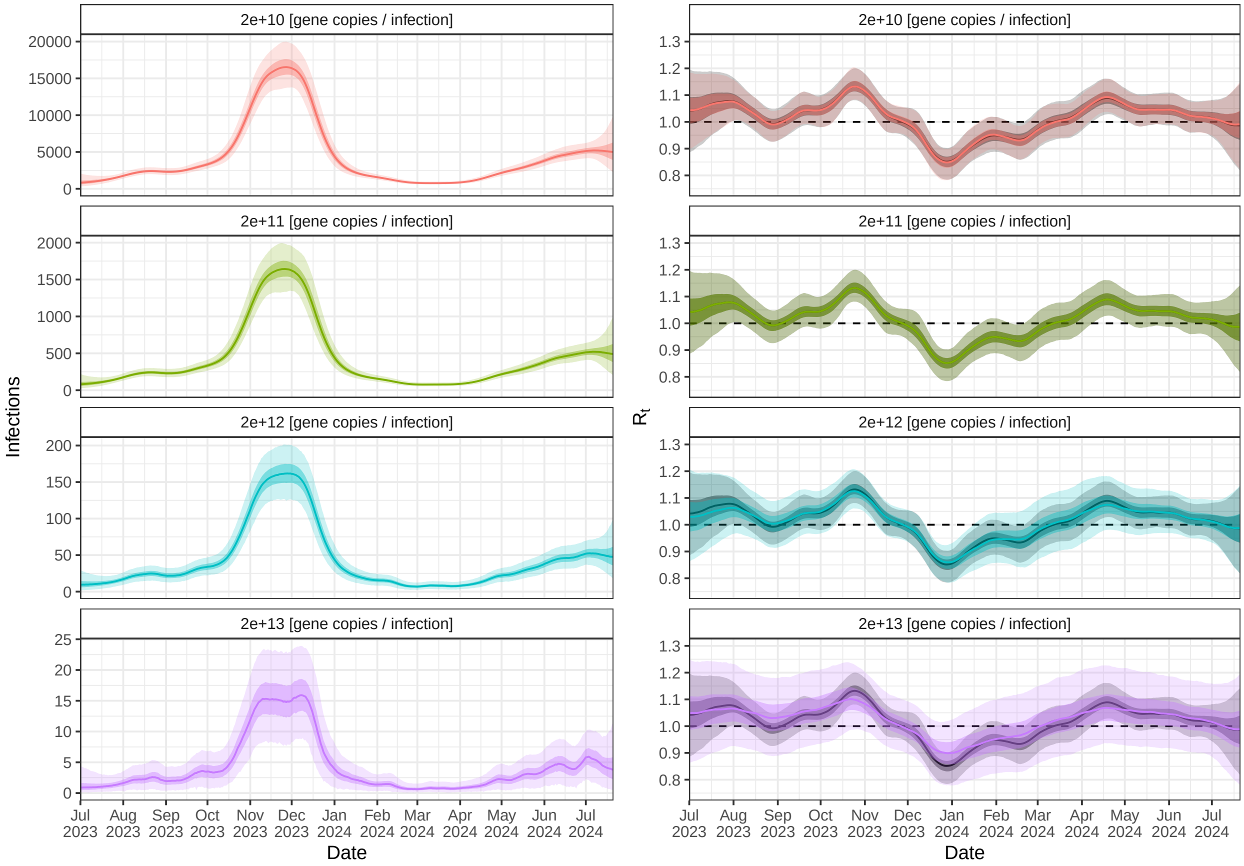

**Fig S16. Sensitivity of estimated transmission dynamics to the assumed shedding load per infection.** Shown are estimated infection numbers and $R_t$ of SARS-CoV-2 in Zurich during the winter season 2023/24, under different assumptions about the total detectable load shed per infection. Higher loads per infection correspond to lower infection numbers, and vice versa. In the main analysis, a load per infection of $2 \times 10^{11}$ gene copies per infection was assumed. For comparison, the corresponding $R_t$ estimate from the main analysis is shown in grey in all panels. Lines indicate the posterior median and dark and light bands the 50% and 95% credible intervals of the marginal posterior distribution, respectively.

Figure S17 furthermore shows a sensitivity analysis of the assumed individual-level variation in shedding loads for the example of RSV in Zurich during the winter season 2023/24. Here we find that $R_t$ estimates are practically identical for assumed coefficients of load variation of 0% (no variation), 100% (as in main text) or 200%. This result does not imply that individual-level variation in shedding has no effect on the noise of pathogen concentrations in wastewater. Rather, any unexplained variation in concentrations is captured by our model of measurement noise, leading to identical estimates of $R_t$. This also underlines the lack of identifiability of individual-level shedding variation from a single time series of concentration measurements with many other potential noise factors.

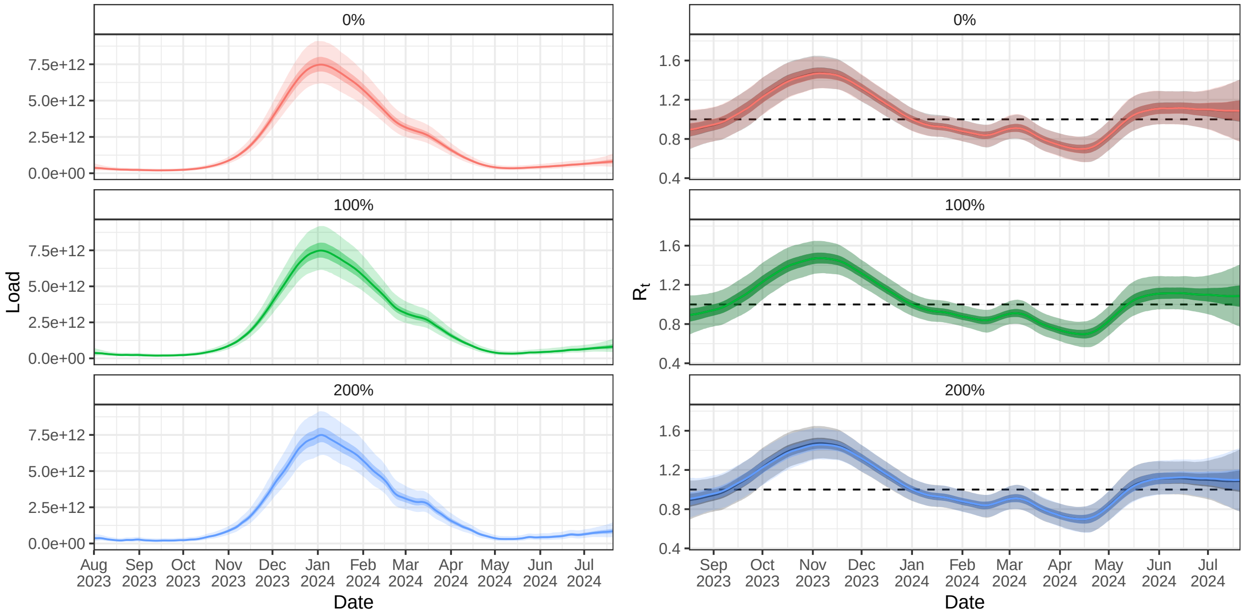

**Fig S17. Sensitivity of estimated transmission dynamics to the assumed individual-level variation in shedding loads.** Shown are estimated total daily loads in wastewater and $R_t$ of RSV in Zurich during the winter season 2023/24, under different assumptions about variation in total load shed by different individuals. In the main analysis, a coefficient of load variation of 100 % was assumed. For comparison, the corresponding $R_t$ estimate from the main analysis is shown in grey in all panels. Lines indicate the posterior median and dark and light bands the 50% and 95% credible intervals of the marginal posterior distribution, respectively.

#### E.4 Subsampling

Figures S18 and S19 show the accuracy and uncertainty of estimated $R_t$ and the variation in trajectories when subsampling our data to different combinations of weekdays. We estimated $R_t$ for all possible weekday combinations of each frequency (3 days per week, 1 day per week, and 1 day per 2 weeks). For a frequency of 3 days per week, $R_t$ estimates deviated only slightly from the baseline (5 days per week) and had comparable uncertainty, with practically no dependence on the specific weekdays sampled. At lower frequencies, median $R_t$ estimates deviated more from the baseline and credible intervals became increasingly wide. We also observed a dependence on the sampled weekdays, especially at the start of the IAV and RSV seasons. During this time period, a high proportion of non-detects means that, depending on the subsampled weekday, no positive measurement may be observed for several weeks. $R_t$ estimates during this time period will then reflect our prior assumption of constant transmission.

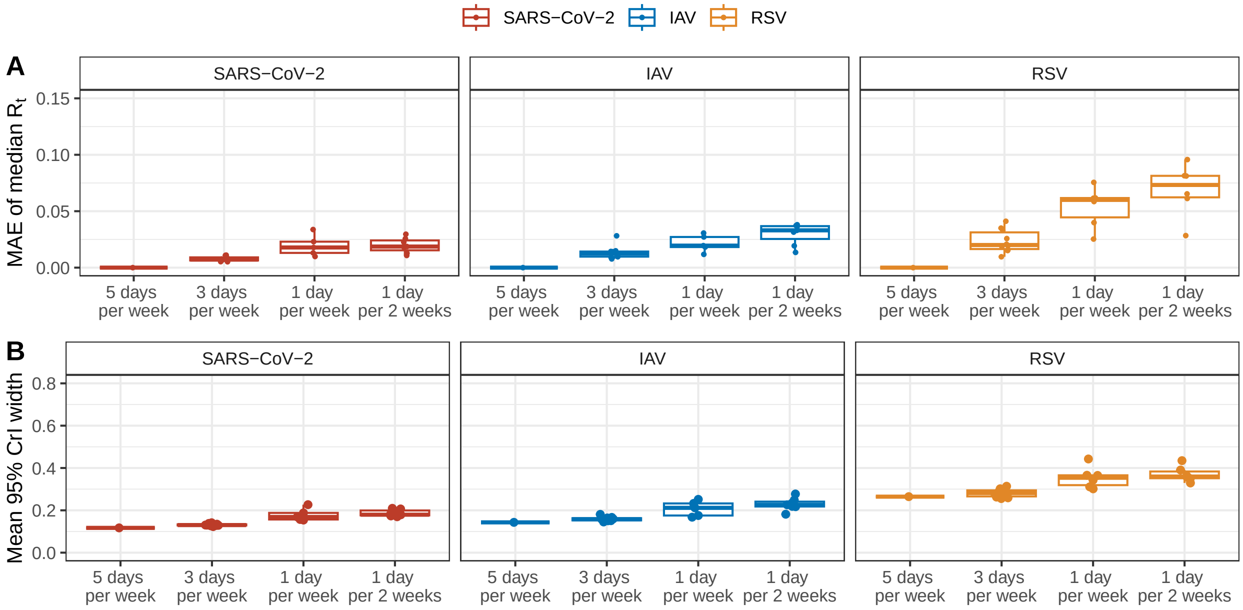

**Fig S18. Accuracy and uncertainty of $R_t$ estimates under sparse measurement.** (A) Mean absolute error (MAE) of median $R_t$ estimates from subsampled measurements compared to the baseline using all measurements (5 days per week), for all possible weekday combinations, respectively. (B) Mean width of the 95% credible intervals for $R_t$ estimates from subsampled measurements, for all possible weekday combinations, respectively. Estimates used data for the catchment of Zurich until May 31, 2024, subsampled to a frequency of 3 days per week, 1 day per week, and 1 day per 2 weeks.

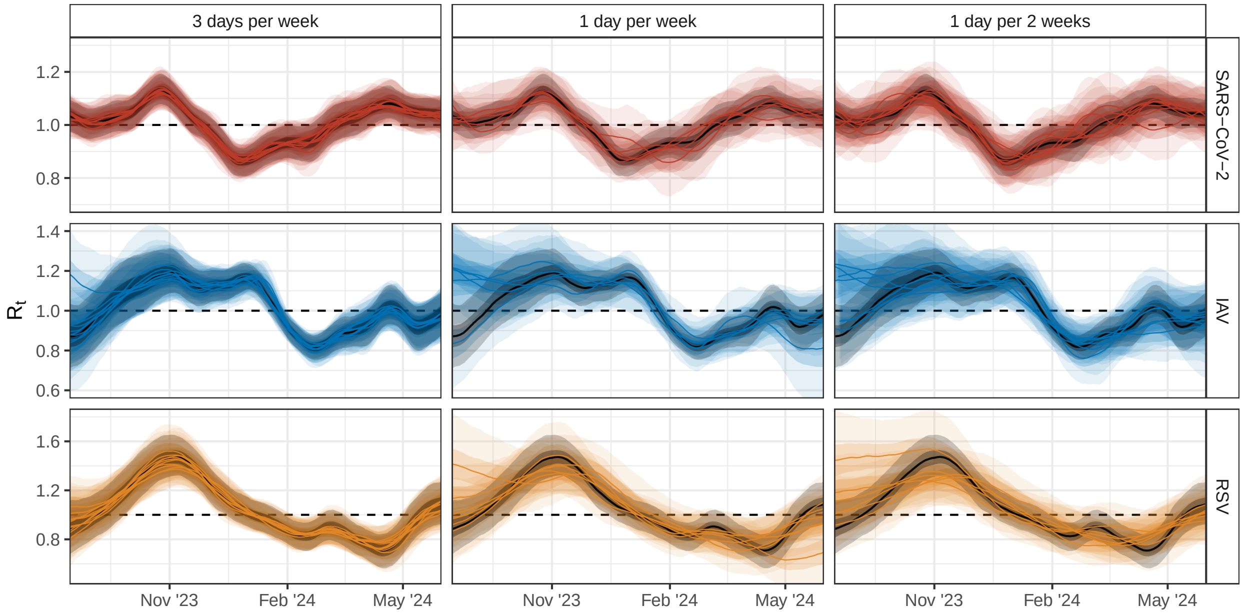

**Fig S19. Sensitivity of $R_t$ estimates to sampled weekdays under sparse measurement.** Retrospective $R_t$ estimates on May 31, 2024 at the catchment of Zurich, based on samples on 3 days per week, 1 day per week, and 1 day per 2 weeks, overlayed for all possible weekday combinations, respectively. The baseline using all available measurements (5 days per week) is shown in grey. Lines indicate the posterior median and dark and light bands the 50% and 95% credible intervals of the marginal posterior distribution, respectively.

#### F Additional results

In the main text, we showed the estimated reproduction number $R_t$ for the catchments of Zurich, Lugano, and Chur during the winter season 2023/24. $R_t$ estimates for these catchments during the winter seasons 2022/23 and 2024/25 are shown in Figures S20 and S21. The real-time performance of $R_t$ estimates and short-term concentration forecasts for these seasons is assessed in Figures S22 and S23. Moreover, Figures S24 to S26 show the real-time mean absolute error (MAE) of median $R_t$ estimates for the winter seasons 2022/23, 2023/24 and 2024/25, respectively. Estimates of the epidemic growth rate $r_t$ for all three winter seasons are shown in Figures S27 to S29. Finally, $R_t$ estimates for all other sampled catchments during the seasons 2022/23, 2023/24, and 2024/25 are shown in Figures S30 to S37.

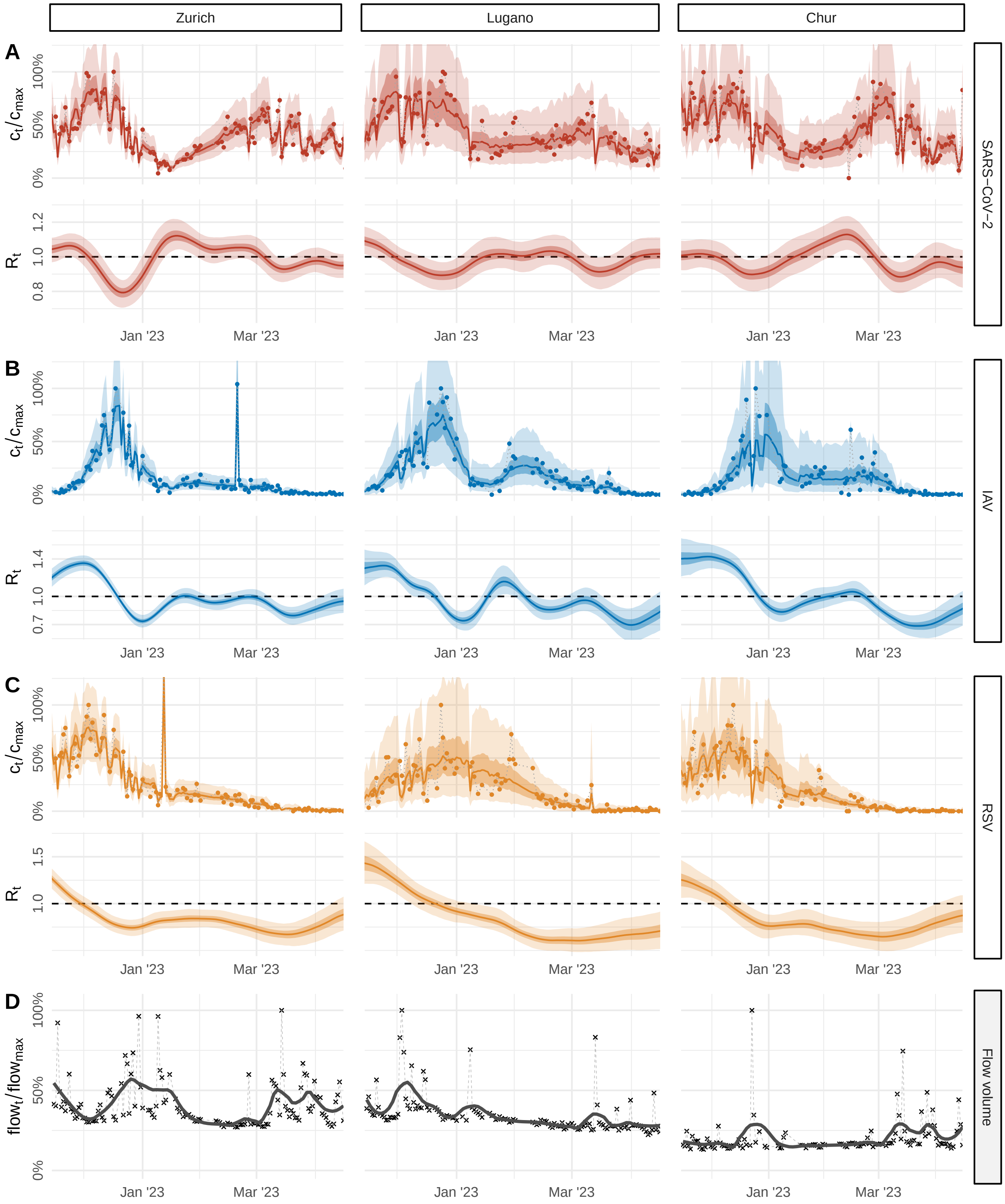

**Fig S20. Viral transmission dynamics in Switzerland quantified from wastewater at treatment plants in Zurich, Chur, and Lugano, 2022/23 winter season.** (A-C) Relative pathogen concentrations (top) and estimates of the effective reproduction number $R_t$ (bottom) for SARS-CoV-2, influenza A virus (IAV), and respiratory syncytial virus (RSV) based on longitudinal samples from a large (Zurich, 471 000 persons), medium-size (Lugano, 124 000 persons), and small (Chur, 55 000 persons) wastewater catchment in Switzerland during the winter season 2022/23. Shown are measured pathogen concentrations (dots), and the posterior median (lines) and 50% and 95% credible intervals (dark and light bands) of predicted concentrations and estimated $R_t$, respectively. (D) Relative daily flow volumes at the sampled wastewater treatment plant of each catchment. Lines show a smooth trend estimated using LOESS with a 4-week window.

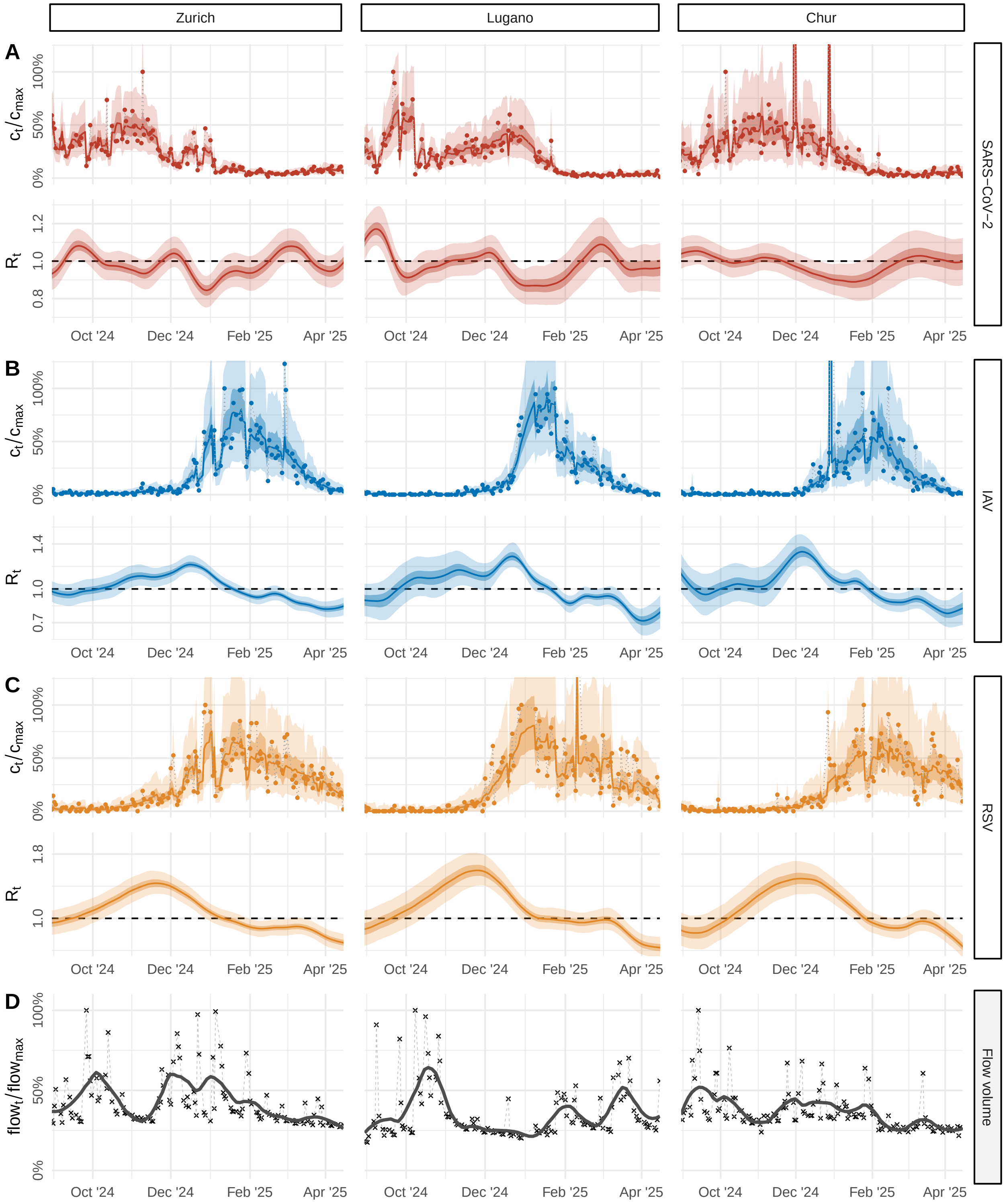

**Fig S21. Viral transmission dynamics in Switzerland quantified from wastewater at treatment plants in Zurich, Chur, and Lugano, 2024/25 winter season.** (A-C) Relative pathogen concentrations (top) and estimates of the effective reproduction number $R_t$ (bottom) for SARS-CoV-2, influenza A virus (IAV), and respiratory syncytial virus (RSV) based on longitudinal samples from a large (Zurich, 471 000 persons), medium-size (Lugano, 124 000 persons), and small (Chur, 55 000 persons) wastewater catchment in Switzerland during the winter season 2024/25. Shown are measured pathogen concentrations (dots), and the posterior median (lines) and 50% and 95% credible intervals (dark and light bands) of predicted concentrations and estimated $R_t$, respectively. (D) Relative daily flow volumes at the sampled wastewater treatment plant of each catchment. Lines show a smooth trend estimated using LOESS with a 4-week window.

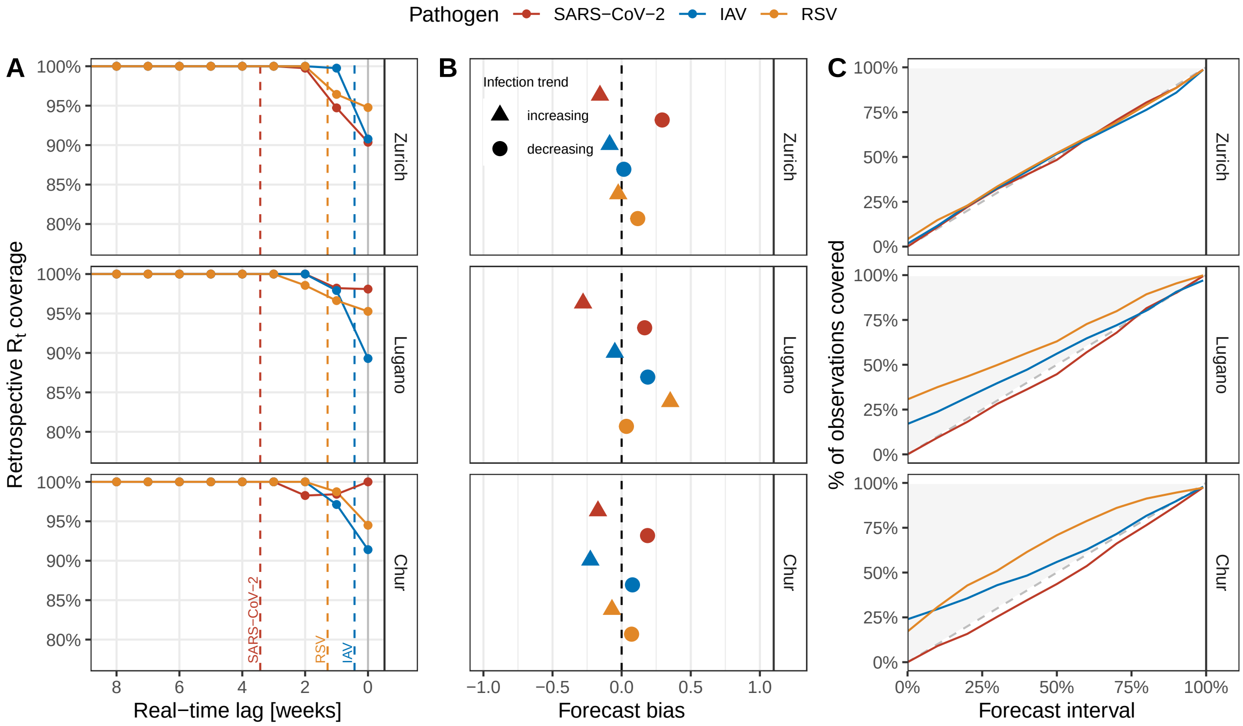

**Fig S22. Real-time performance of wastewater-based reproduction number estimates and short-term concentration forecasts during the winter season 2022/23.** Shown is the performance of real-time $R_t$ estimates and 1–14 day ahead forecasts of concentration measurements for the wastewater catchments of Zurich, Lugano, and Chur, Switzerland, during the 2022/23 seasonal wave of SARS-CoV-2 (red), IAV (blue), and RSV (orange). (A) Consistency of real-time $R_t$ (estimated on each day with new data) with retrospective $R_t$ (estimated at the end of the seasonal wave). For each real-time $R_t$ estimate, we computed the retrospective coverage, i. e. the percentage of days on which the 95% credible interval (CrI) contained the retrospective median $R_t$. Dots show the coverage stratified by lags of 0–8 weeks from the date of estimation. Vertical dashed lines show the 90% quantile of the shedding load distribution of each pathogen. (B) Bias of concentration measurement forecasts, stratified by increasing and decreasing infection trends. Scores range between -1 (maximal underprediction) and 1 (maximal overprediction); 0 indicates no bias. (C) Calibration of concentration measurement forecasts. The diagonal line indicates a perfectly calibrated forecast, where all intervals cover exactly their nominal share of observations.

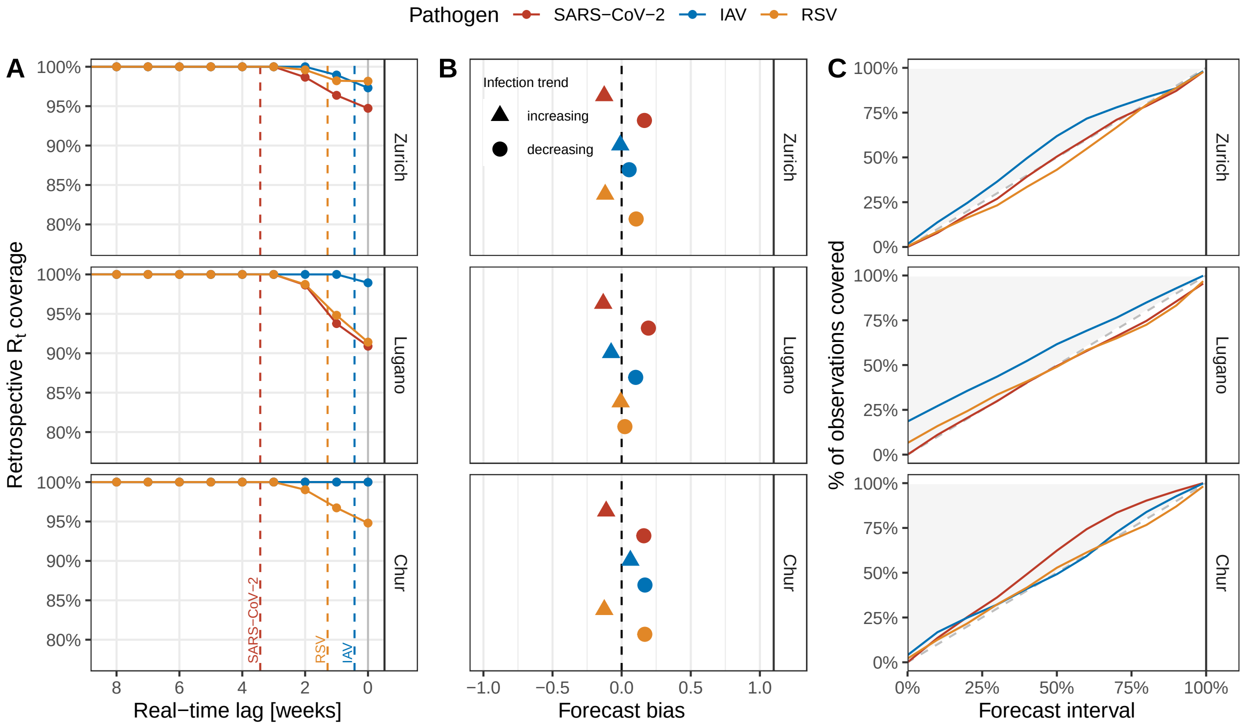

**Fig S23. Real-time performance of wastewater-based reproduction number estimates and short-term concentration forecasts during the winter season 2024/25.** Shown is the performance of real-time $R_t$ estimates and 1–14 day ahead forecasts of concentration measurements for the wastewater catchments of Zurich, Lugano, and Chur, Switzerland, during the 2024/25 seasonal wave of SARS-CoV-2 (red), IAV (blue), and RSV (orange). (A) Consistency of real-time $R_t$ (estimated on each day with new data) with retrospective $R_t$ (estimated at the end of the seasonal wave). For each real-time $R_t$ estimate, we computed the retrospective coverage, i. e. the percentage of days on which the 95% credible interval (CrI) contained the retrospective median $R_t$. Dots show the coverage stratified by lags of 0–8 weeks from the date of estimation. Vertical dashed lines show the 90% quantile of the shedding load distribution of each pathogen. Note that for Chur, coverage was 100% at all lags for both SARS-CoV-2 and IAV. (B) Bias of concentration measurement forecasts, stratified by increasing and decreasing infection trends. Scores range between -1 (maximal underprediction) and 1 (maximal overprediction); 0 indicates no bias. (C) Calibration of concentration measurement forecasts. The diagonal line indicates a perfectly calibrated forecast, where all intervals cover exactly their nominal share of observations.

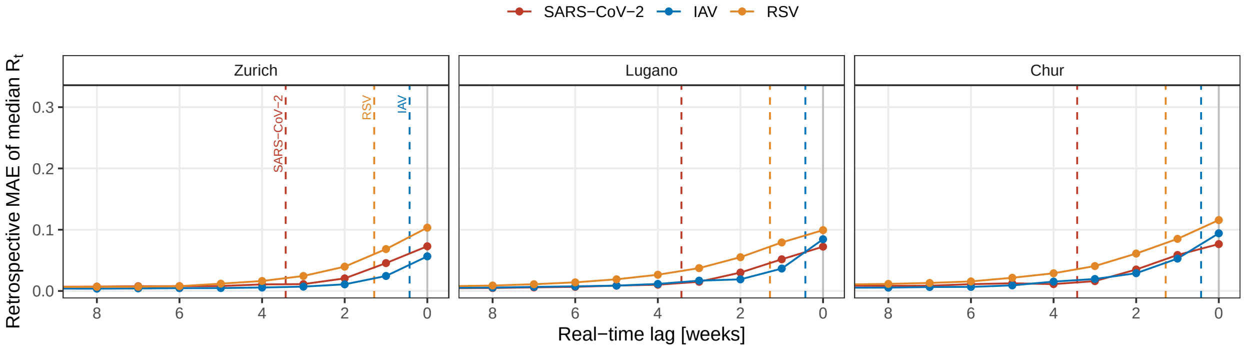

**Fig S24. Real-time accuracy of wastewater-based reproduction number estimates during the winter season 2022/23.** Dots show the mean absolute error (MAE) of real-time median $R_t$ (estimated on each day with new data) compared to the retrospective median $R_t$ (estimated at the end of the seasonal wave) for the wastewater catchments of Zurich, Lugano, and Chur, Switzerland, during the 2022/23 seasonal wave of SARS-CoV-2 (red), IAV (blue), and RSV (orange).

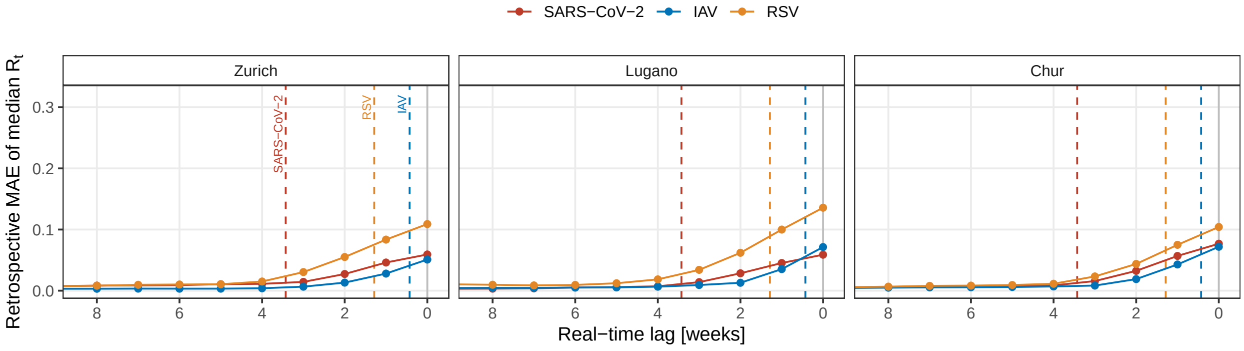

**Fig S25. Real-time accuracy of wastewater-based reproduction number estimates during the winter season 2023/24.** Dots show the mean absolute error (MAE) of real-time median $R_t$ (estimated on each day with new data) compared to the retrospective median $R_t$ (estimated at the end of the seasonal wave) for the wastewater catchments of Zurich, Lugano, and Chur, Switzerland, during the 2023/24 seasonal wave of SARS-CoV-2 (red), IAV (blue), and RSV (orange).

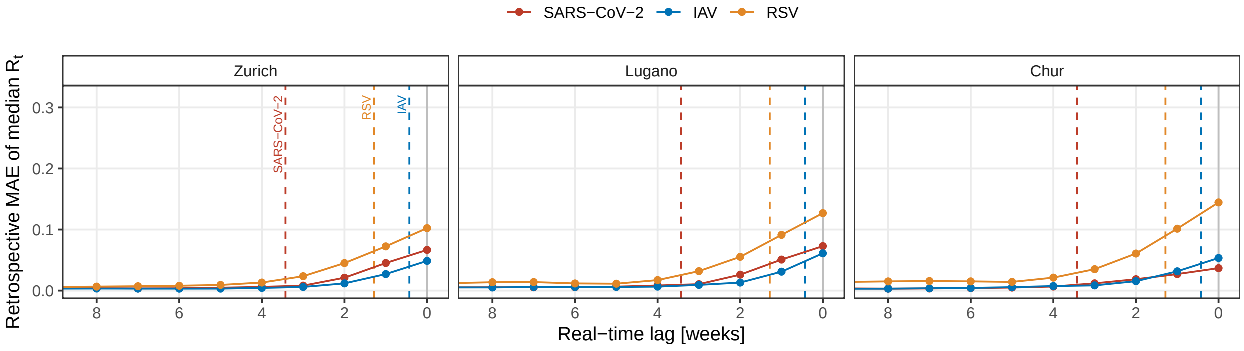

**Fig S26. Real-time accuracy of wastewater-based reproduction number estimates during the winter season 2024/25.** Dots show the mean absolute error (MAE) of real-time median $R_t$ (estimated on each day with new data) compared to the retrospective median $R_t$ (estimated at the end of the seasonal wave) for the wastewater catchments of Zurich, Lugano, and Chur, Switzerland, during the 2024/25 seasonal wave of SARS-CoV-2 (red), IAV (blue), and RSV (orange).

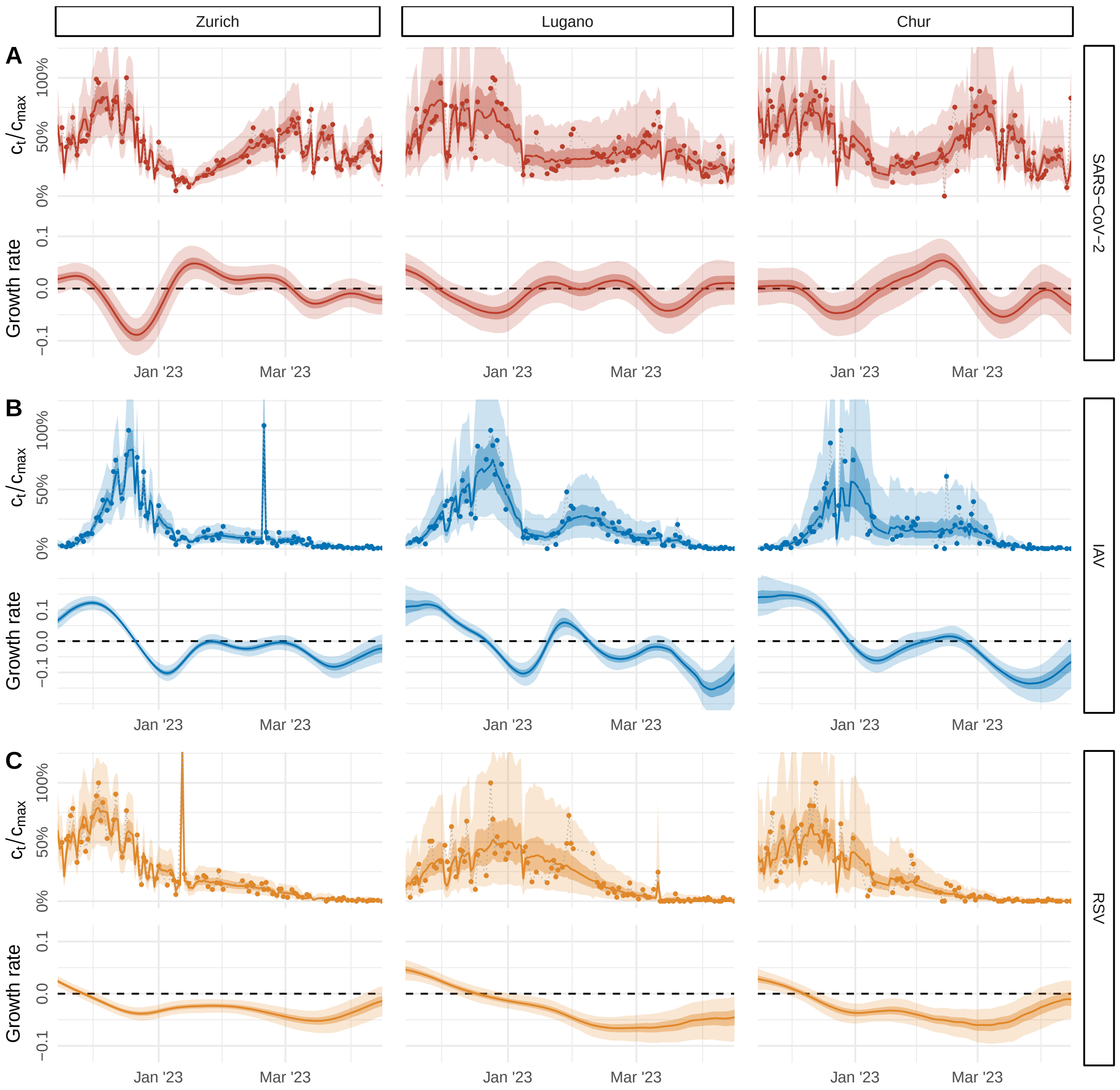

**Fig S27. Estimated epidemic growth rates during the winter season 2022/23.** (A-C) Relative pathogen concentrations (top) and estimates of the epidemic growth rate $r_t$ (bottom) for SARS-CoV-2, influenza A virus (IAV), and respiratory syncytial virus (RSV) based on longitudinal samples from a large (Zurich, 471 000 persons), medium-size (Lugano, 124 000 persons), and small (Chur, 55 000 persons) wastewater catchment in Switzerland during the winter season 2022/23. Shown are measured pathogen concentrations (dots), and the posterior median (lines) and 50% and 95% credible intervals (dark and light bands) of predicted concentrations and estimated $r_t$, respectively. (D) Relative daily flow volumes at the sampled wastewater treatment plant of each catchment. Lines show a smooth trend estimated using LOESS with a 4-week window.

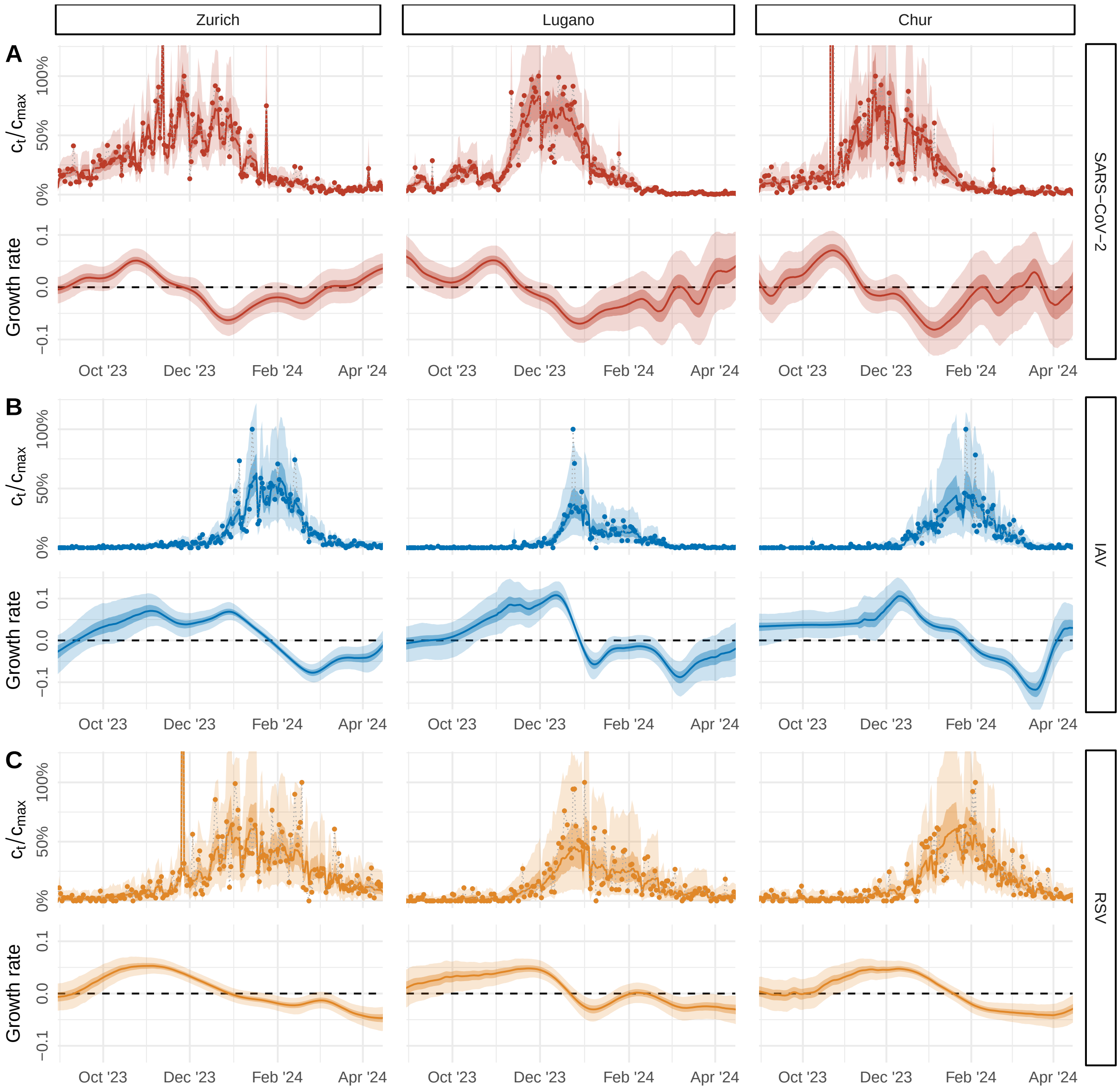

**Fig S28. Estimated epidemic growth rates during the winter season 2023/24.** (A-C) Relative pathogen concentrations (top) and estimates of the epidemic growth rate $r_t$ (bottom) for SARS-CoV-2, influenza A virus (IAV), and respiratory syncytial virus (RSV) based on longitudinal samples from a large (Zurich, 471 000 persons), medium-size (Lugano, 124 000 persons), and small (Chur, 55 000 persons) wastewater catchment in Switzerland during the winter season 2023/24. Shown are measured pathogen concentrations (dots), and the posterior median (lines) and 50% and 95% credible intervals (dark and light bands) of predicted concentrations and estimated $r_t$, respectively. (D) Relative daily flow volumes at the sampled wastewater treatment plant of each catchment. Lines show a smooth trend estimated using LOESS with a 4-week window.

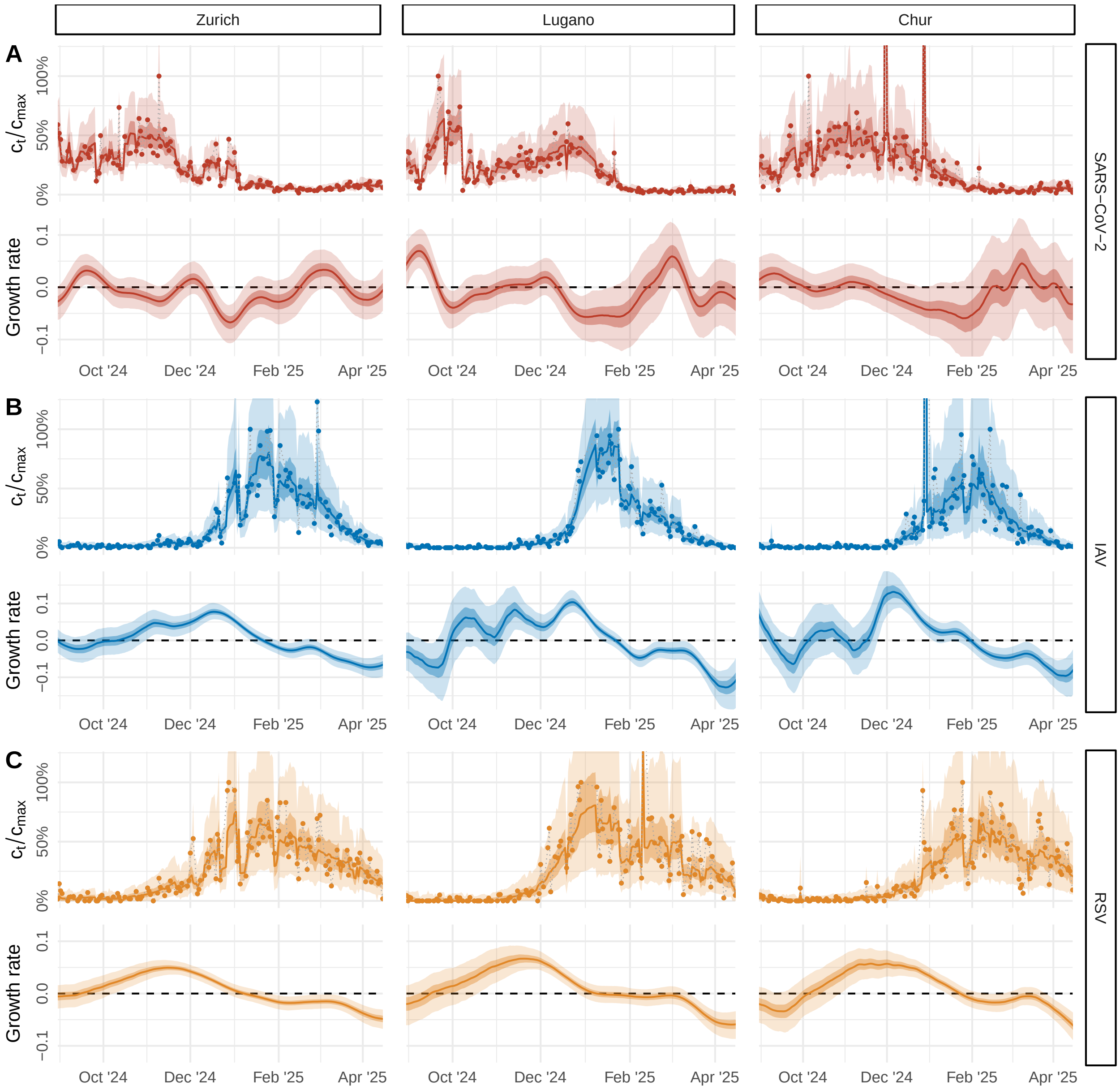

**Fig S29. Estimated epidemic growth rates during the winter season 2024/25.** (A-C) Relative pathogen concentrations (top) and estimates of the epidemic growth rate $r_t$ (bottom) for SARS-CoV-2, influenza A virus (IAV), and respiratory syncytial virus (RSV) based on longitudinal samples from a large (Zurich, 471 000 persons), medium-size (Lugano, 124 000 persons), and small (Chur, 55 000 persons) wastewater catchment in Switzerland during the winter season 2024/25. Shown are measured pathogen concentrations (dots), and the posterior median (lines) and 50% and 95% credible intervals (dark and light bands) of predicted concentrations and estimated $r_t$, respectively. (D) Relative daily flow volumes at the sampled wastewater treatment plant of each catchment. Lines show a smooth trend estimated using LOESS with a 4-week window.

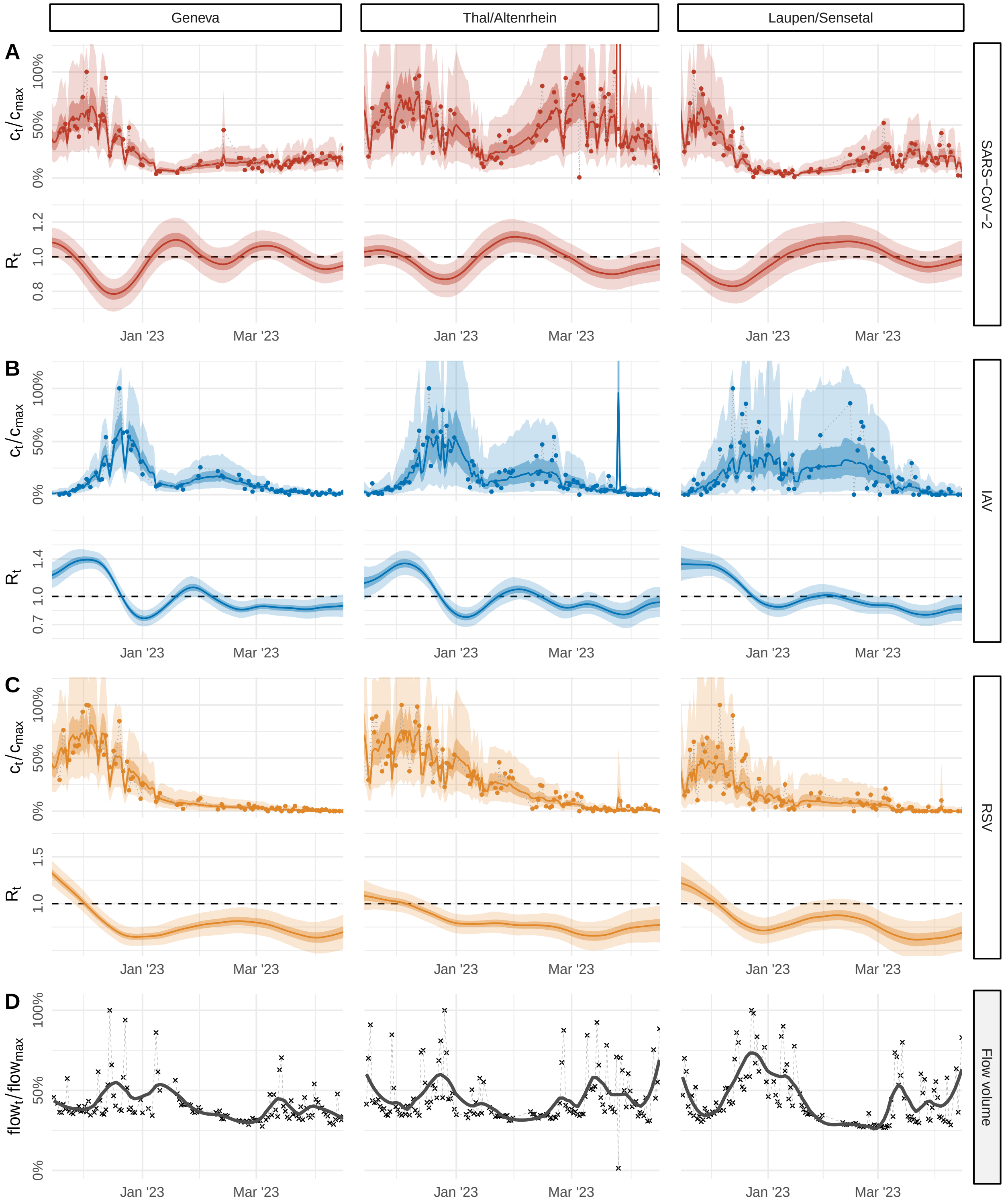

**Fig S30. $R_t$ estimates for additional catchments: Geneva, Thal, and Laupen, winter season 2022/23.** (A-C) Relative pathogen concentrations (top) and estimates of the effective reproduction number $R_t$ (bottom) for SARS-CoV-2, influenza A virus (IAV), and respiratory syncytial virus (RSV) based on longitudinal samples from the wastewater catchments of Geneva (454 000 persons), Thal / Altenrhein region (64 000 persons), and Laupen / Sensetal region (62 000 persons) during the winter season 2022/23. Shown are measured pathogen concentrations (dots), and the posterior median (lines) and 50% and 95% credible intervals (dark and light bands) of predicted concentrations and estimated $R_t$, respectively. (D) Relative daily flow volumes at the sampled wastewater treatment plant of each catchment. Lines show a smooth trend estimated using LOESS with a 4-week window.

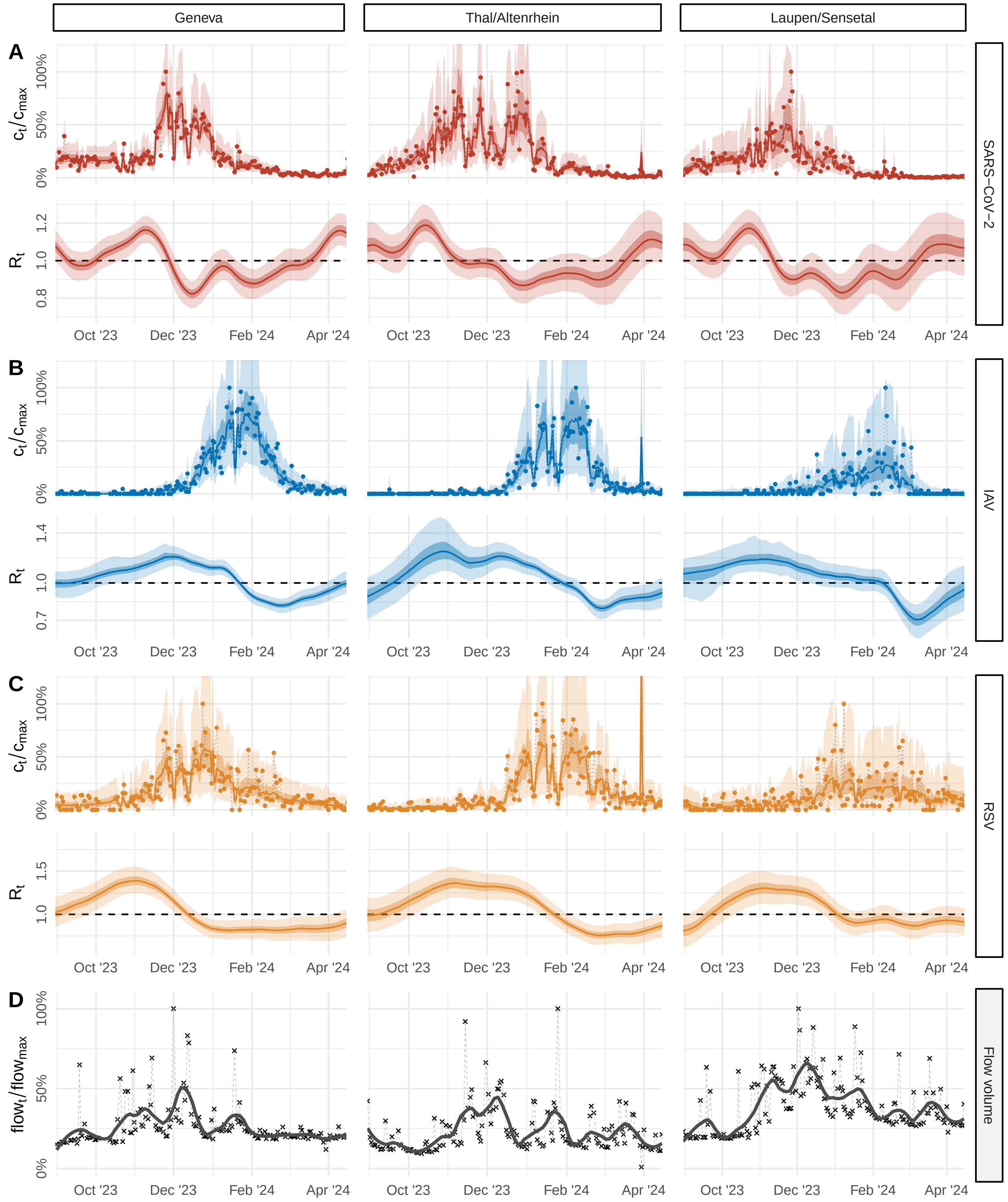

**Fig S31. $R_t$ estimates for additional catchments: Geneva, Thal, and Laupen, winter season 2023/24.** (A-C) Relative pathogen concentrations (top) and estimates of the effective reproduction number $R_t$ (bottom) for SARS-CoV-2, influenza A virus (IAV), and respiratory syncytial virus (RSV) based on longitudinal samples from the wastewater catchments of Geneva (454 000 persons), Thal / Altenrhein region (64 000 persons), and Laupen / Sensetal region (62 000 persons) during the winter season 2023/24. Shown are measured pathogen concentrations (dots), and the posterior median (lines) and 50% and 95% credible intervals (dark and light bands) of predicted concentrations and estimated $R_t$, respectively. (D) Relative daily flow volumes at the sampled wastewater treatment plant of each catchment. Lines show a smooth trend estimated using LOESS with a 4-week window.

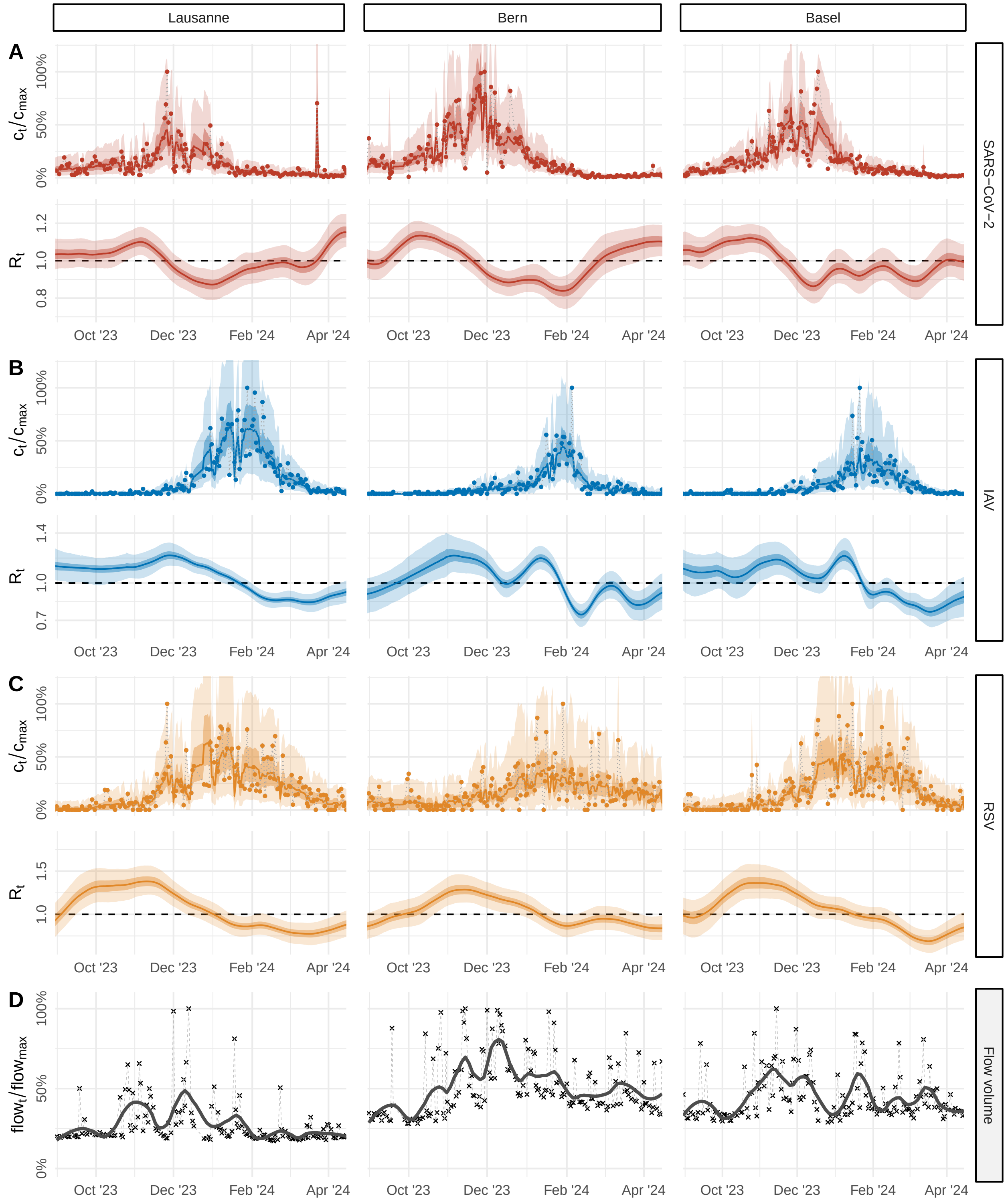

**Fig S32. $R_t$ estimates for additional catchments: Lausanne, Bern, and Basel, winter season 2023/24.** (A-C) Relative pathogen concentrations (top) and estimates of the effective reproduction number $R_t$ (bottom) for SARS-CoV-2, influenza A virus (IAV), and respiratory syncytial virus (RSV) based on longitudinal samples from the wastewater catchments of Lausanne (240 000 persons), Bern (225 000 persons), and Basel (268 000 persons) during the winter season 2023/24. Shown are measured pathogen concentrations (dots), and the posterior median (lines) and 50% and 95% credible intervals (dark and light bands) of predicted concentrations and estimated $R_t$, respectively. (D) Relative daily flow volumes at the sampled wastewater treatment plant of each catchment. Lines show a smooth trend estimated using LOESS with a 4-week window.

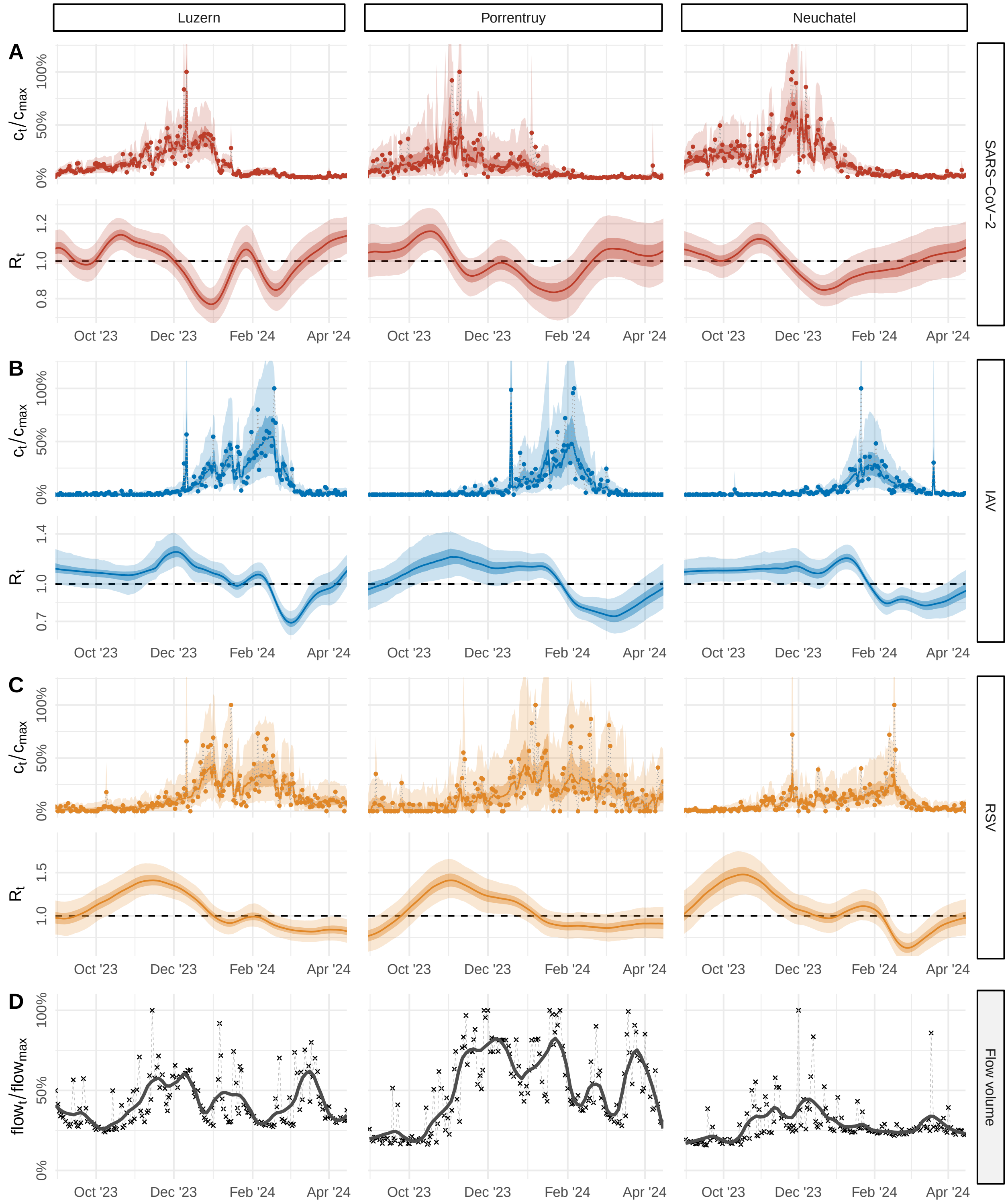

**Fig S33.** **$R_t$ estimates for additional catchments: Luzern, Porrentruy, and Neuchatel, winter season 2023/24.** (A-C) Relative pathogen concentrations (top) and estimates of the effective reproduction number $R_t$ (bottom) for SARS-CoV-2, influenza A virus (IAV), and respiratory syncytial virus (RSV) based on longitudinal samples from the wastewater catchments of Luzern (179 000 persons), Porrentruy (17 000 persons), and Neuchatel (41 000 persons) during the winter season 2023/24. Shown are measured pathogen concentrations (dots), and the posterior median (lines) and 50% and 95% credible intervals (dark and light bands) of predicted concentrations and estimated $R_t$, respectively. (D) Relative daily flow volumes at the sampled wastewater treatment plant of each catchment. Lines show a smooth trend estimated using LOESS with a 4-week window.

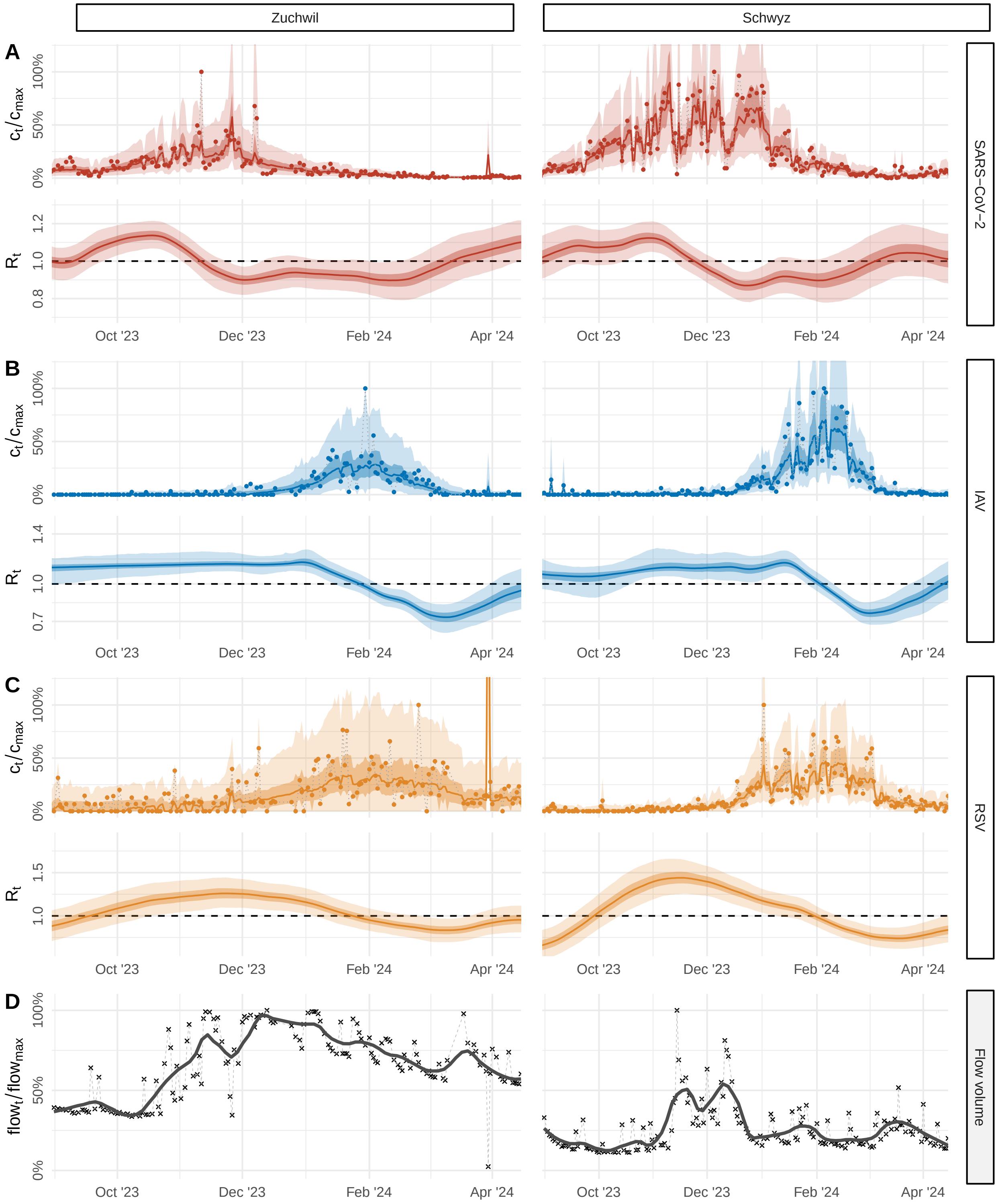

**Fig S34. $R_t$ estimates for additional catchments: Zuchwil and Schwyz, winter season 2023/24.** (A-C) Relative pathogen concentrations (top) and estimates of the effective reproduction number $R_t$ (bottom) for SARS-CoV-2, influenza A virus (IAV), and respiratory syncytial virus (RSV) based on longitudinal samples from the wastewater catchments of Zuchwil (96 000 persons) and Schwyz (31 000 persons) during the winter season 2023/24. Shown are measured pathogen concentrations (dots), and the posterior median (lines) and 50% and 95% credible intervals (dark and light bands) of predicted concentrations and estimated $R_t$, respectively. (D) Relative daily flow volumes at the sampled wastewater treatment plant of each catchment. Lines show a smooth trend estimated using LOESS with a 4-week window.

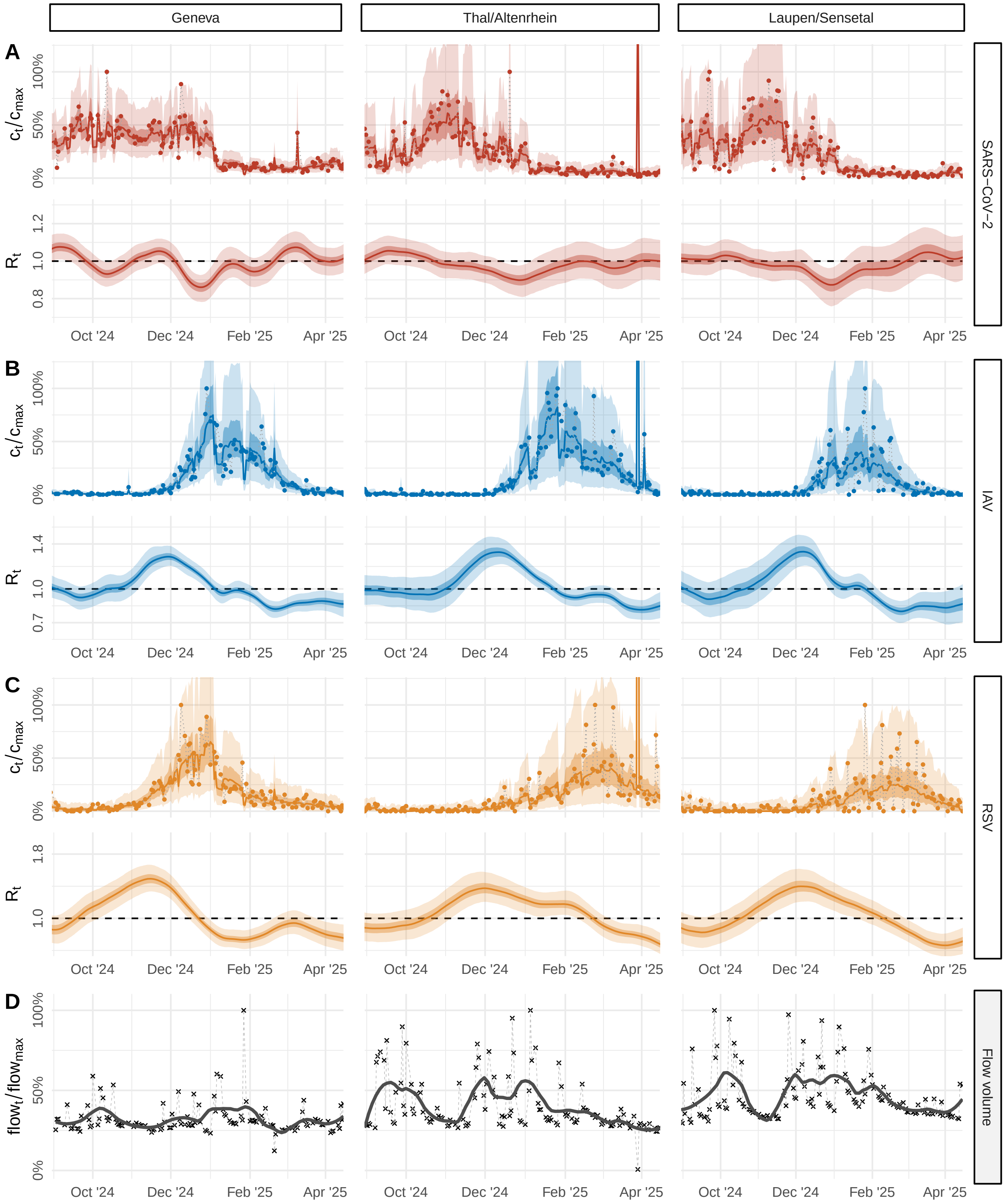

**Fig S35. $R_t$ estimates for additional catchments: Geneva, Thal, and Laupen, winter season 2024/25.** (A-C) Relative pathogen concentrations (top) and estimates of the effective reproduction number $R_t$ (bottom) for SARS-CoV-2, influenza A virus (IAV), and respiratory syncytial virus (RSV) based on longitudinal samples from the wastewater catchments of Geneva (454 000 persons), Thal / Altenrhein region (64 000 persons), and Laupen / Sensetal region (62 000 persons) during the winter season 2024/25. Shown are measured pathogen concentrations (dots), and the posterior median (lines) and 50% and 95% credible intervals (dark and light bands) of predicted concentrations and estimated $R_t$, respectively. (D) Relative daily flow volumes at the sampled wastewater treatment plant of each catchment. Lines show a smooth trend estimated using LOESS with a 4-week window.

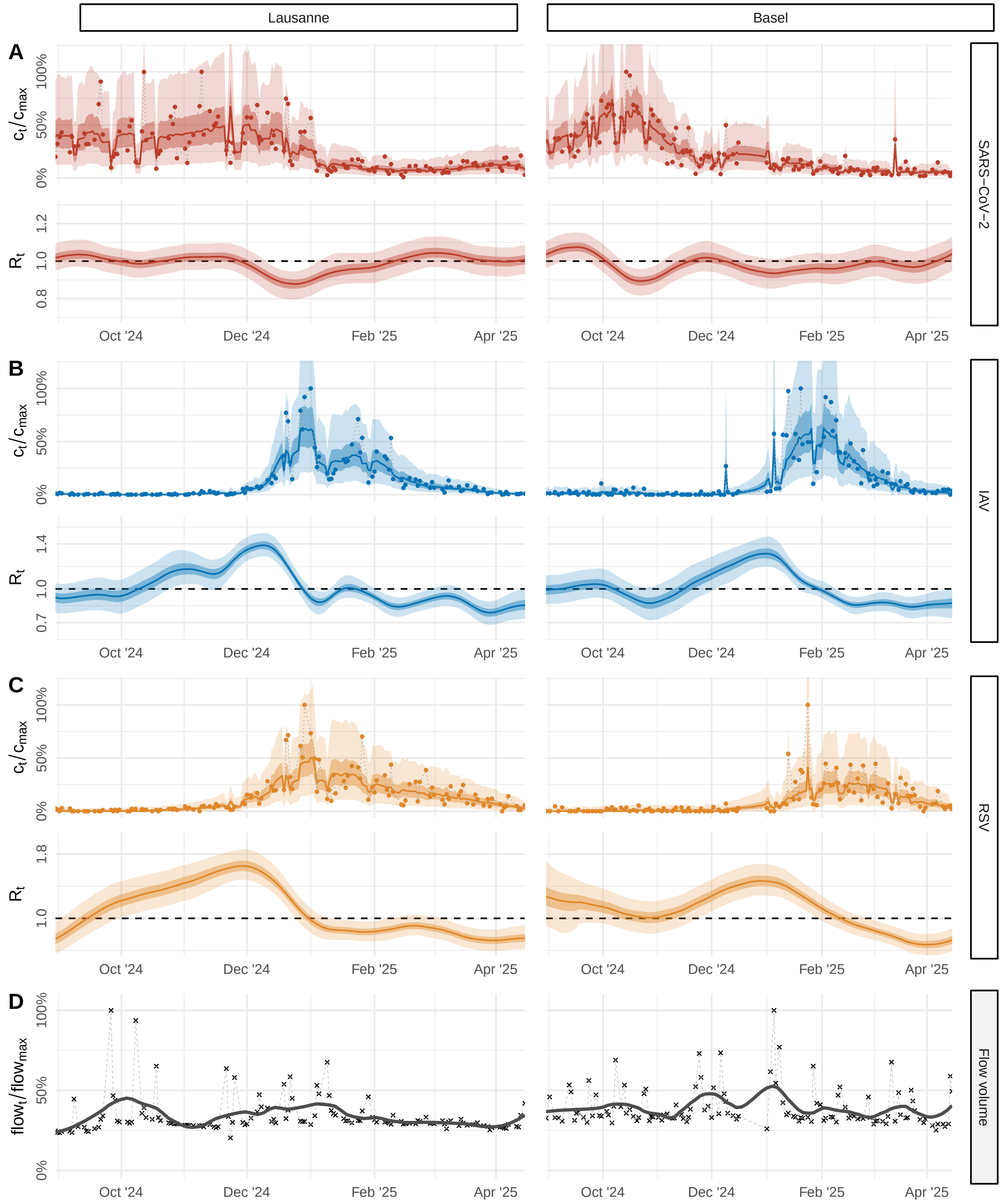

**Fig S36.** $R_t$ **estimates for additional catchments: Lausanneand Basel, winter season 2024/25.** (A-C) Relative pathogen concentrations (top) and estimates of the effective reproduction number $R_t$ (bottom) for SARS-CoV-2, influenza A virus (IAV), and respiratory syncytial virus (RSV) based on longitudinal samples from the wastewater catchments of Lausanne (240 000 persons) and Basel (268 000 persons) during the winter season 2024/25. Shown are measured pathogen concentrations (dots), and the posterior median (lines) and 50% and 95% credible intervals (dark and light bands) of predicted concentrations and estimated $R_t$, respectively. (D) Relative daily flow volumes at the sampled wastewater treatment plant of each catchment. Lines show a smooth trend estimated using LOESS with a 4-week window.

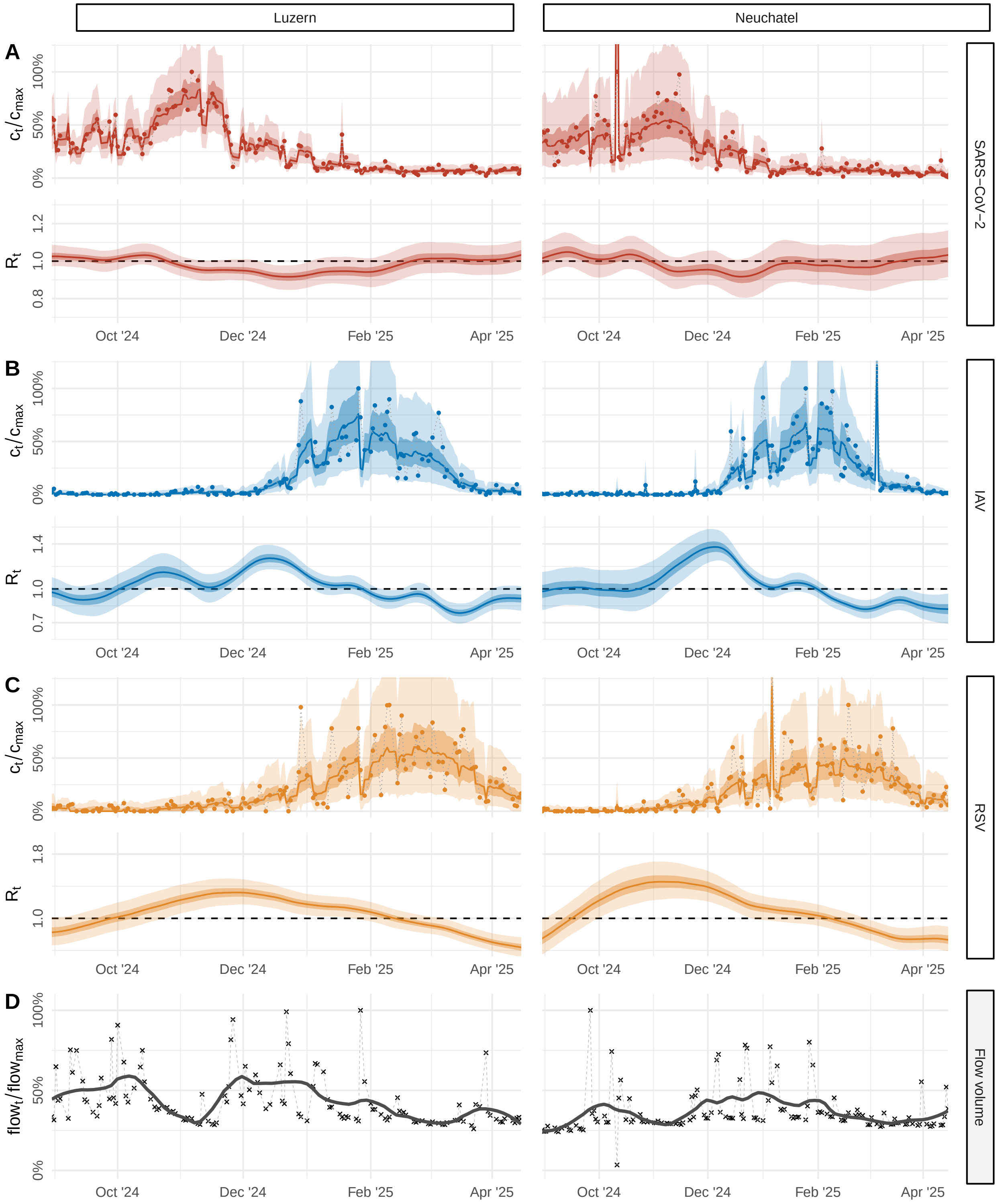

**Fig S37. $R_t$ estimates for additional catchments: Luzern and Neuchatel, winter season 2024/25.** (A-C) Relative pathogen concentrations (top) and estimates of the effective reproduction number $R_t$ (bottom) for SARS-CoV-2, influenza A virus (IAV), and respiratory syncytial virus (RSV) based on longitudinal samples from the wastewater catchments of Luzern (179 000 persons) and Neuchatel (41 000 persons) during the winter season 2024/25. Shown are measured pathogen concentrations (dots), and the posterior median (lines) and 50% and 95% credible intervals (dark and light bands) of predicted concentrations and estimated $R_t$, respectively. (D) Relative daily flow volumes at the sampled wastewater treatment plant of each catchment. Lines show a smooth trend estimated using LOESS with a 4-week window.

**Movie S1. Real-time $R_t$ estimation and concentration forecasting for SARS-CoV-2 in Zurich, winter season 2023/24.** Top panel shows concentration forecasts and observed concentrations (stars), normalized to median flow. Bottom panel shows real-time estimates and 14-day-ahead projections of $R_t$, along with retrospective $R_t$ (in gray). Lines indicate posterior medians; shaded bands represent 50% (dark) and 95% (light) credible intervals.

**Movie S2. Real-time $R_t$ estimation and concentration forecasting for Influenza A in Zurich, winter season 2023/24.** Top panel shows concentration forecasts and observed concentrations (stars), normalized to median flow. Bottom panel shows real-time estimates and 14-day-ahead projections of $R_t$, along with retrospective $R_t$ (in gray). Lines indicate posterior medians; shaded bands represent 50% (dark) and 95% (light) credible intervals.

**Movie S3. Real-time $R_t$ estimation and concentration forecasting for RSV in Zurich, winter season 2023/24.** Top panel shows concentration forecasts and observed concentrations (stars), normalized to median flow. Bottom panel shows real-time estimates and 14-day-ahead projections of $R_t$, along with retrospective $R_t$ (in gray). Lines indicate posterior medians; shaded bands represent 50% (dark) and 95% (light) credible intervals.

**22.** Teh YW, Elesedy B, He B, Hutchinson M, Zaidi S, Bhoopchand A, et al. Efficient Bayesian Inference of Instantaneous Reproduction Numbers at Fine Spatial Scales, with an Application to

Mapping and Nowcasting the Covid-19 Epidemic in British Local Authorities. Journal of the Royal Statistical Society Series A: Statistics in Society. 2022;185(Supplement_1):S65–S85.
