## Supplementary figures and images for "Robust real-time estimation of pathogen transmission dynamics from wastewater"

### Movie S1

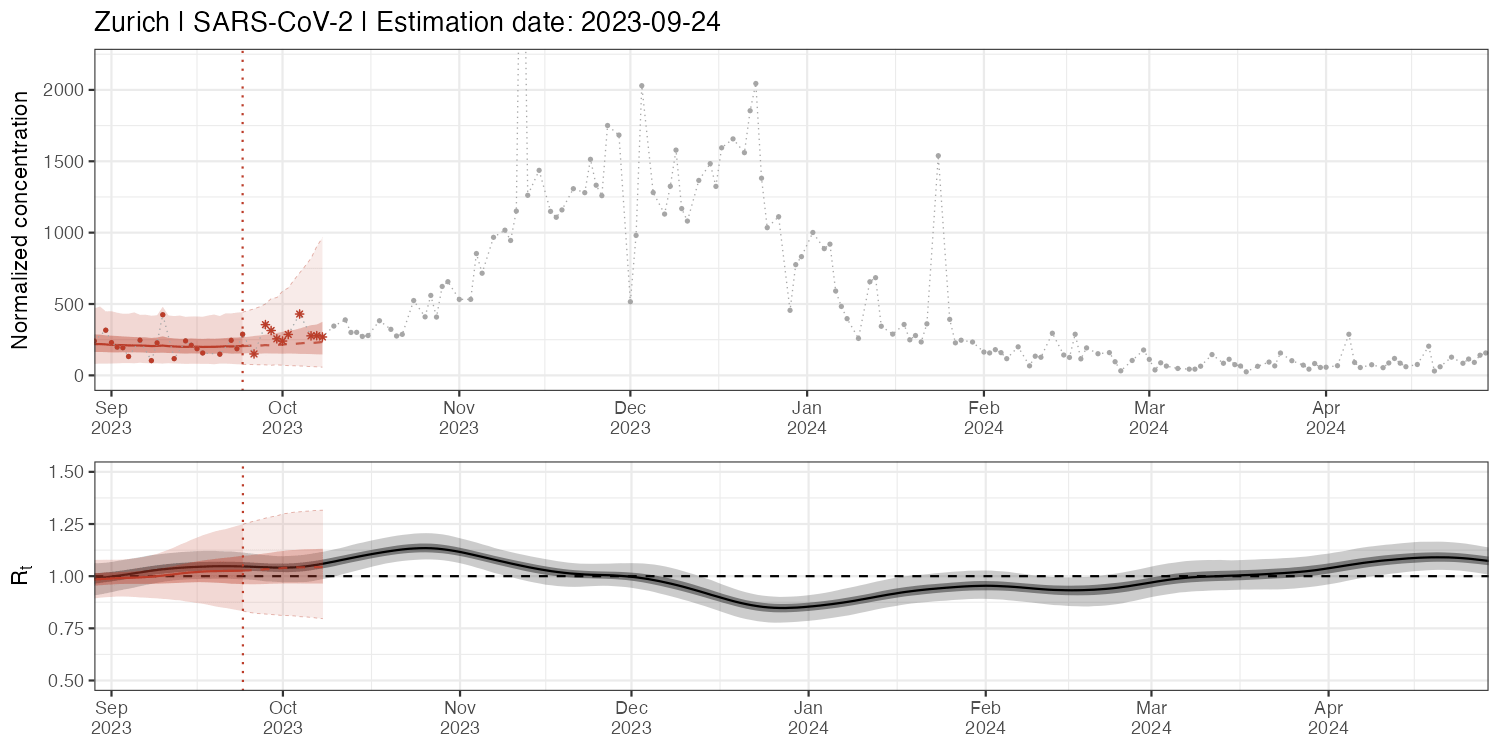

### Movie S2

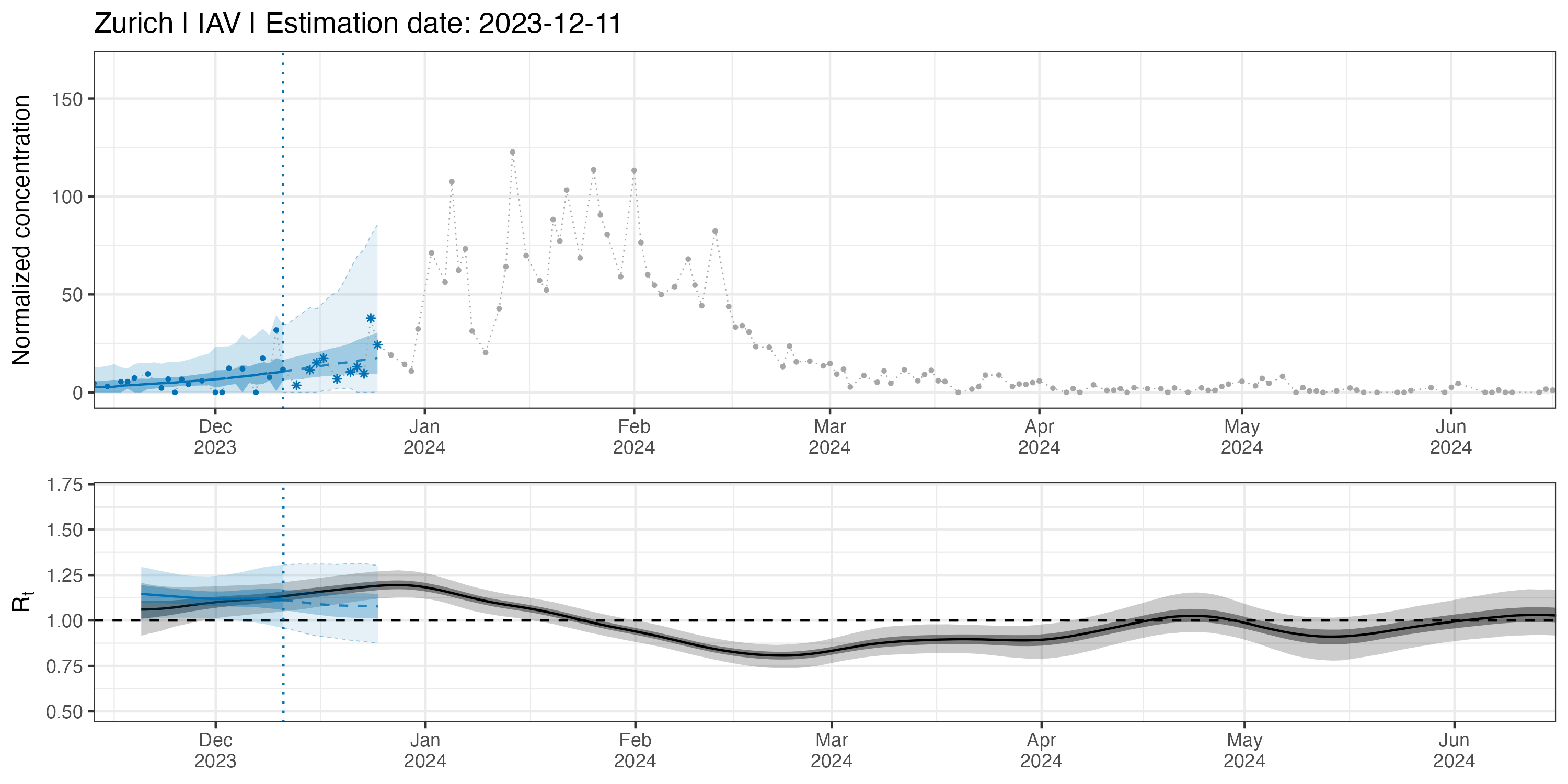

### Movie S3

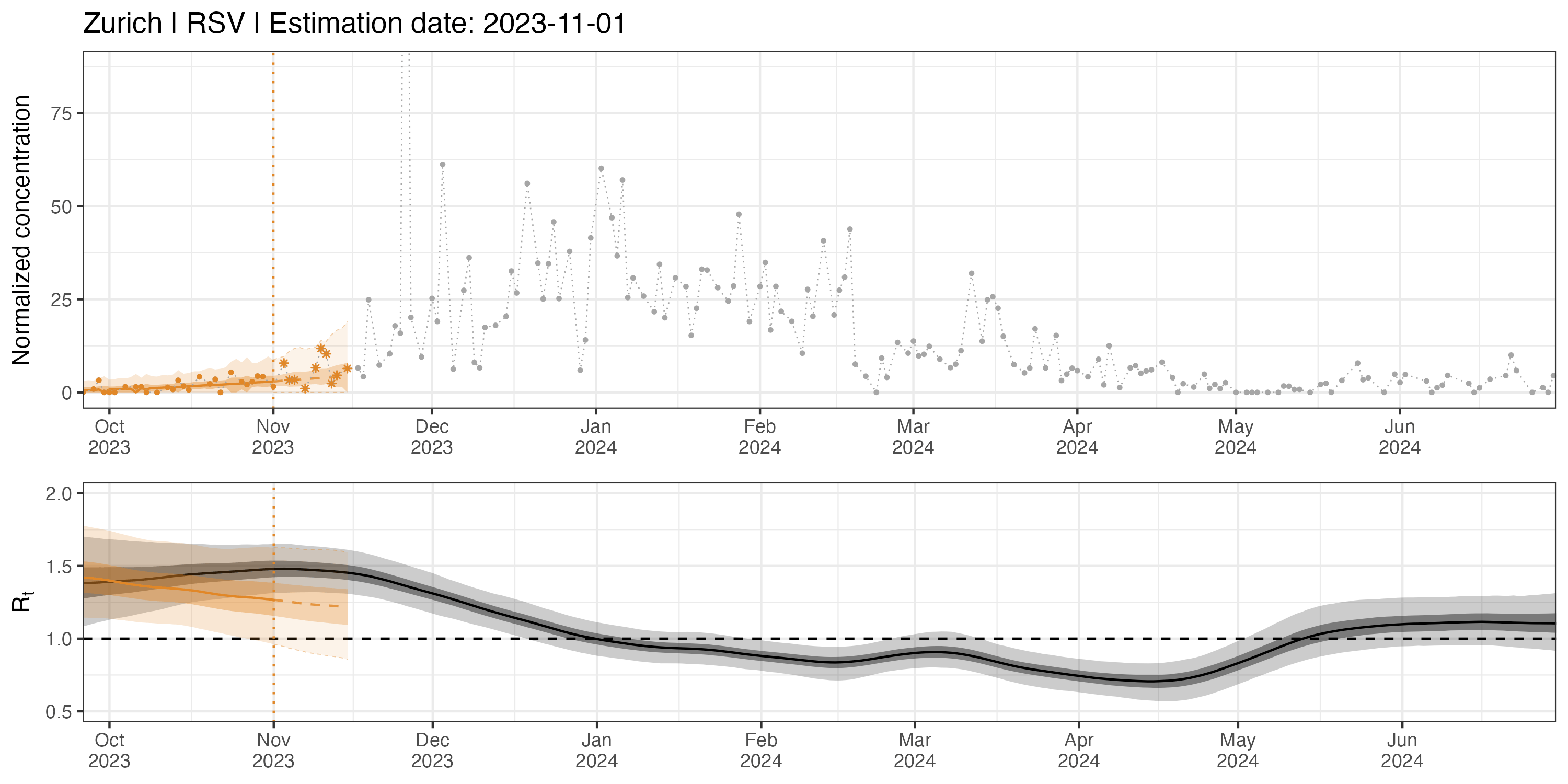
